# Behavioral dynamics of COVID-19: estimating under-reporting, multiple waves, and adherence fatigue across 92 nations

**DOI:** 10.1101/2020.06.24.20139451

**Authors:** Hazhir Rahmandad, Tse Yang Lim, John Sterman

**Affiliations:** Massachusetts Institute of Technology, Cambridge, MA

## Abstract

Effective responses to the COVID-19 pandemic require integrating behavioral factors such as risk-driven contact reduction, improved treatment, and adherence fatigue with asymptomatic transmission, disease acuity, and hospital capacity. We build one such model and estimate it for all 92 nations with reliable testing data. Cumulative cases and deaths through 22 December 2020 are estimated to be 7.03 and 1.44 times official reports, yielding an infection fatality rate (IFR) of 0.51% which has been declining over time. Absent adherence fatigue cumulative cases would have been 47% lower. Scenarios through June 2021 show that modest improvement in responsiveness could reduce cases and deaths ≈14%, more than the impact of vaccinating half of the population by that date. Variations in responsiveness to risk explain two orders of magnitude difference in per-capita deaths despite reproduction numbers fluctuating ∼ 1 across nations. A public online simulator facilitates scenario analysis over the coming months.

## Introduction

Effective responses to the COVID-19 pandemic require an understanding of its global magnitude and the drivers of variations in outbreaks across nations. Yet more than ten months after WHO declared a global pandemic, the true number of cases and infection fatality rate remain uncertain, and the experience of different nations varies widely. As of late December 2020, countries have reported cumulative cases ranging between 7.03 and 7200 per 100,000, and case fatality rates between 0.05% and 9.0%. Asymptomatic infection (Gudbjartsson, Helgason et al. 2020), variation in testing rates across countries (Roser, Ritchie et al. 2020), and false negatives (Fang, Zhang et al. 2020, Li, Yao et al. 2020, Wang, Xu et al. 2020) complicate assessment of the true magnitude of the pandemic from official data. The inference problem also requires disentangling other explanatory mechanisms: (i) differences in population density and social networks create variations in the effective reproduction number, R_E_; (ii) risk perceptions, behavior change, adherence fatigue, and policy responses alter transmission rates endogenously; (iii) testing is prioritized based on symptoms and risk factors, so detection depends on both testing rates and current prevalence (Onder, Rezza et al. 2020); (iv) limited hospital capacity is allocated based on case severity, affecting fatality rates; (v) age, socio-economic status, comorbidities, differential adherence to non-pharmaceutical interventions (NPIs) such as social distancing and masking, and improvements in treatment affect transmission risk and case severity (Guan, Ni et al. 2020, O’Driscoll, Dos Santos et al. 2020, Wu and McGoogan 2020); (vi) weather may play a role in transmission (Xu, Rahmandad et al. 2020); and (vii) all these vary across nations.

Prior studies have shed light on important parts of the puzzle by estimating basic epidemiological parameters and IFR in well-controlled settings (Russell, Hellewell et al. 2020), assessing the asymptomatic fraction and prevalence in specific populations (Hao, Cheng et al. 2020, Li, Pei et al. 2020, Mizumoto, Kagaya et al. 2020, Salje, Tran Kiem et al. 2020, Sutton, Fuchs et al. 2020), estimating the role of undocumented infections (Ghaffarzadegan and Rahmandad 2020, Li, Pei et al. 2020, Moghadas, Fitzpatrick et al. 2020), illuminating the effects of various NPIs on expected future cases (Chinazzi, Davis et al. 2020, Flaxman, Mishra et al. 2020, Hsiang, Allen et al. 2020, Kissler, Tedijanto et al. 2020, Ruktanonchai, Floyd et al. 2020, Walker, Whittaker et al. 2020, Wu, Leung et al. 2020), and contrasting risks and healthcare demands across countries and populations (Britton, Ball et al. 2020, Gatto, Bertuzzo et al. 2020, Moghadas, Shoukat et al. 2020, Struben 2020, Walker, Whittaker et al. 2020).

Endogenous changes in behaviors and policies in response to outbreaks could also drive large variations in outcomes across nations. However, classical epidemic models, such as the Susceptible-Exposed-Infectious-Recovered (SEIR) model, do not include endogenous behavior and policy responses. The classical models project exponential outbreaks that are ultimately limited by depletion of the susceptible pool or by the exogenous introduction of interventions that lower the effective reproduction rate (R_E_). Prior work in system dynamics shows that endogenous behavioral responses to infectious diseases significantly alter the dynamics. Sterman (2001, Chapter 9.4) describes a wide range of behavioral feedbacks in the context of the HIV/AIDS epidemic, and shows that endogenous responses to risk can shift the dynamics from a single outbreak to an endemic condition with periods of reduced incidence followed by large outbreaks. Further, Rahmandad and Sterman (2008) showed that the impact of endogenous behavior change in the SEIR model is large compared to variations caused by different values of epidemic parameters determining the basic reproduction rate, R_0_, and by different structures for the contact network among individuals.

The history of the COVID-19 pandemic to date strongly suggests a role for behavioral feedbacks. Many nations have experienced multiple waves of COVID-19 as outbreaks induced behavior change and government policies that reduced transmission, only to see second or even third waves as the resulting reductions in incidence and deaths eroded NPI adherence by individuals and relaxation of government policies. To fit the data, many COVID-19 models capture changes in transmission rates as a function of time; (2) using data on policy adoption timing in specific nations; or (3) by incorporating policy switches that turn on in response to risks (Flaxman, Mishra et al. 2020, Kissler, Tedijanto et al. 2020, Li, Pei et al. 2020, Walker, Whittaker et al. 2020). These approaches improve the fit of models to past data but perform poorly in counter-factual experiments and long-term projections where historical data are uninformative and extrapolation of time-based functions unreliable. A smaller group of studies have taken a more endogenous view of responses where ongoing (Ghaffarzadegan and Rahmandad 2020, Struben 2020) or expected risks (Acemoglu, Chernozhukov et al. 2020, Farboodi, Jarosch et al. 2020) condition contact rates and transmission. Prior work, however, has not used these risk-driven response functions to explain the large cross-national variations in outcomes (Struben (2020) provides an exception).

Overall, we lack a global view of the pandemic that is simultaneously consistent with these more focused findings, explains the orders-of-magnitude variation in official per-capita case and death rates across countries, and offers reliable projections consistent with multiple waves of mortality and incidence (Lopez and Rodo 2020).

### Model

We use a multi-country modified SEIR model to simultaneously estimate SARS-CoV-2 transmission across 92 countries (all nations that report data sufficient to enable estimation). For each country, the model tracks the population from susceptible through pre-symptomatic, infected, and recovered or deceased states, with explicit stocks for those whose cases are detected or undetected, and hospitalized or not (Figure *1*A). Infection moves people from the *Susceptible* population (S) into the *Pre-Symptomatic Infected* stock. After an *Incubation Period*, these pre-symptomatic individuals flow into the *Infected Pre-Detection* stock. After a further *Onset to Detection Delay*, this group splits among multiple pathways. First, those testing positive for COVID-19 flow into either *Detected COVID+ Not Hospitalized* or *Detected COVID+ Hospitalized*. Infected individuals who do not receive a positive test result, whether for lack of testing or a false negative test result, transition into either *Undetected COVID+* or *Undetected COVID+ Hospitalized*. We assume demand for testing and hospitalization are driven by symptoms, so all asymptomatic individuals will be in the *Undetected COVID+* stock.

**Figure 1.**
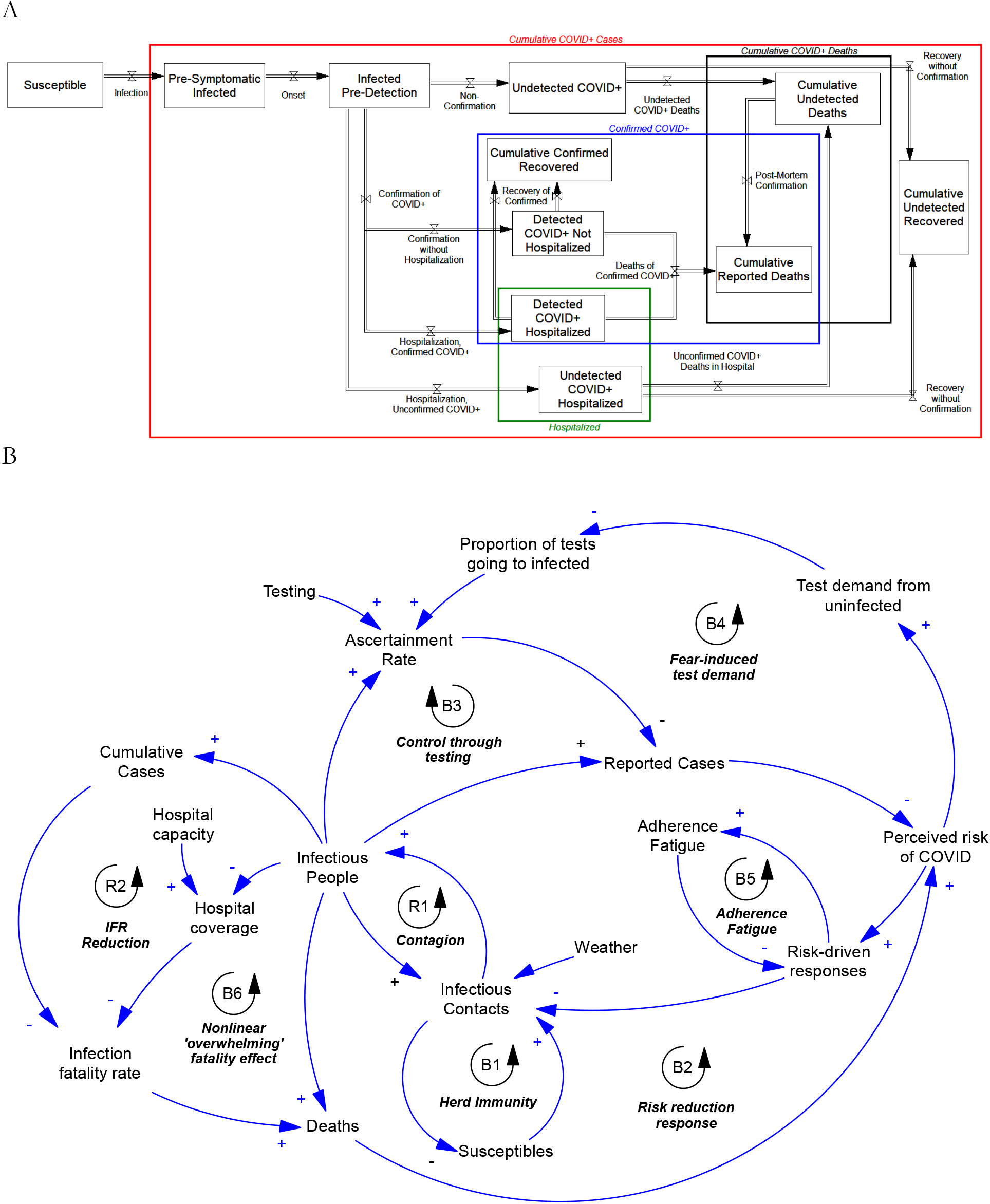
Model overview. A) Expanded SEIR model for each country highlighting key stocks and flows. B) Major feedbacks captured in the model.

In addition to its multi-country scope, the model includes three novel features (Figure *1*B). First, tests are allocated to individuals based on symptom severity relative to available testing capacity. Individuals with more severe symptoms, including those with COVID-19 and those without but presenting with similar symptoms (e.g., influenza) or risk factors (e.g., frontline healthcare workers), get priority for testing. We model symptom severity with a zero-inflated Poisson distribution, where zero severity indicates asymptomatic infection. In this framework, originally developed in the context of testing in project models (Rahmandad and Hu 2010), each increment of symptom severity increases the chance of receiving a COVID test, become hospitalized, or die. The formulation provides a consistent method to capture the significant heterogeneity in symptom severity, from life-threatening to asymptomatic cases, and its impact on testing and hospitalization, without the need to disaggregate the population by different severity levels. Such disaggregation would make parameter estimation prohibitively time consuming (S1 and Figure S3 provide details). Besides its computational benefits, the formulation keeps the number of free (estimated) parameters to a minimum, specifically two: the fraction of asymptomatic cases and the mean severity of symptomatic COVID+ individuals. Severity-prioritized test allocation determines the ascertainment rate of COVID-positive cases as a function of prevalence and the current testing rate. It also results in different average COVID severity in the tested vs. untested populations. We also account for false negatives from tests (Fang, Zhang et al. 2020, Wang, Xu et al. 2020).

Second, hospital capacity is allocated between COVID-positive cases and demand from non-COVID patients. The COVID infection fatality rate therefore depends endogenously on the adequacy of hospital capacity relative to the burden of severe cases, along with the age distribution of the population (Guan, Ni et al. 2020, O’Driscoll, Dos Santos et al. 2020, Wu and McGoogan 2020). Furthermore, we account for reductions in IFR that may result from improved treatments, deaths of high-risk populations such as the elderly and those with comorbidities in the first wave of the pandemic, heterogeneous adherence to NPIs as higher-risk subpopulations perceive greater risk than those lacking risk factors, shifting incidence toward younger, lower-risk people, and other factors.

Third, the hazard rate of transmission responds to the perceived risk of COVID. Perceived risk reduces transmission through adoption of NPIs, from social distancing and masking to lockdowns. Perceived risk is based on subjective perceptions of the hazard of death, which respond with a lag to both official data (reported in the media) and actual deaths (gleaned from personal experience and word of mouth). Rising mortality eventually increases perceived risk, driving the hazard rate of transmission down, while declining cases and deaths can lead to the erosion of perceived risk, potentially leading to rebound outbreaks. We also account for adherence fatigue, modeled as a reduction in the impact of perceived risk on behaviors that reduce contacts. We use the recent (100-day) reduction in contacts compared to pre-pandemic levels as the driver of adherence fatigue, and estimate a country-level parameter quantifying resulting reductions in the population’s responsiveness to increases in perceived risk.

### Estimation

Model parameters are specified based on prior literature and formal estimation. Parameters specified from the literature include the incubation period (mean *μ* = 5 days (Guan, Ni et al. 2020, Linton, Kobayashi et al. 2020)), onset-to-detection delay (*μ* = 5 days (Linton, Kobayashi et al. 2020)), post-onset illness duration (*μ* = 15 days (Guan, Ni et al. 2020)), and the sensitivity of RT-PCR-based testing (70% (Fang, Zhang et al. 2020, Wang, Xu et al. 2020)). Sensitivity analysis is presented in S7.

We estimate the remaining parameters using a panel of data covering all nations with at least 1000 confirmed COVID-19 cases by 22 December 2020 and sufficient testing data to enable parameter estimation, a total of 92 nations spanning 4.92 billion people. These include all disease hotspots to date, with two notable exceptions, China and Brazil, for which reliable testing data are not available. The panel includes reported daily testing rates, reported COVID cases and deaths, and all-cause mortality (where available), along with population, population density, age distribution, hospital capacity, and daily meteorological data.

The model is nonlinear and complex, and any estimation framework is unlikely to have clean analytical solutions or provable bounds on errors and biases. Therefore, in designing our estimation procedure we apply three guideposts: (1) being conservative by incorporating uncertainties; (2) avoiding over-fitting; and (3) enhancing the generalizability and robustness of estimates and projections. To do so we use a negative binomial likelihood function, which accommodates overdispersion and autocorrelation; we keep the number of estimated parameters to the minimum feasible, typically one or two for each mechanism; we utilize a hierarchical Bayesian framework (Gelman and Hill 2006) to couple parameter estimates across different countries, reducing the risk of over-fitting the data; and we use existing knowledge characterizing parameters and their expected similarity across countries to inform the priors for the magnitude of that coupling across countries. For example, the asymptomatic fraction of cases and other parameters representing biological processes should have low cross-national variance, whereas parameters specifying risk perceptions and responses are expected to vary more widely (see S7 for sensitivity analysis on priors). Compared to more common choices in similar estimation settings (e.g. Gaussian likelihood functions), these choices tend to widen the credible regions for our estimates and reduce the quality of the fit between model and data by having fewer parameters and imposing coupling among them. In return, the results may better capture uncertainties, are more informative about the underlying processes, and provide more reliable projections. For example, better fits to individual nations could be obtained by estimating every parameter separately for each nation, or including more free parameters, but doing so would dramatically increase the number of parameters to be estimated, would falsely assume that the experience of every nation is completely independent of all others, and could yield estimates that differ significantly across countries even for parameters, such as the asymptomatic fraction of cases, that should vary little across nations on an age-adjusted basis.

The estimation method identifies both the most likely value and the credible regions for the unknown parameters, given the data on reported cases and deaths (and for a subset of countries, excess total mortality). To avoid overfitting, we do not use any time-varying parameters in the estimation. The maximum likelihood method described above and in the supplement yields the estimated parameters. To quantify the uncertainties in parameters and projections we use a Markov Chain Monte Carlo method designed for high dimensional parameter spaces (Vrugt, Ter Braak et al. 2009); see S2 for details.

### Building confidence in the model

Estimating parameters for a complex model and assessing its ability to capture important real-world processes are both critical and challenging. Before discussing results we present four sets of analyses addressing these challenges, and later report extensive sensitivity analyses to quantify various uncertainties. First, we compare model outputs to data. Figure *2*A compares actual and simulated reported daily new cases and deaths for 18 larger countries using data through 22 December 2020. S5 and S6 report the full sample. Over the full set of nations and full sample, Mean Absolute Errors Normalized by the mean of the actual data (MAEN) are 5.6% and 5.0% for cumulative infections and deaths, respectively, and less than 20% for 69 (75.0%) and 64 (69.6%) of the 92 nations, despite wide variation in the size and dynamics of national outbreaks, from those nearly quenched (New Zealand, Thailand), to those still growing (e.g. Latvia, Hungary) to those exhibiting multiple waves (e.g. Iran, Israel, USA). R^2^ exceeds 0.9 for 54% of 368 country-specific time series and exceeds 0.5 for 86%. Aggregation of within-nation heterogeneities reduces the quality of fit in a few countries. For example, we do not explicitly model outbreaks concentrated among subpopulations such as migrant workers (important in e.g., Qatar, Singapore) or nursing homes (important in, e.g., Belgium, France). The coupling of parameters across countries also limits the fit for outliers. Moreover, because the model does not include any time-varying parameters (e.g., Thanksgiving gatherings in the USA, school and university schedules, and other calendrical events that condition contacts), the model is expected to miss some important events. Nevertheless, 80% and 85% of official infection and death rates fall within the 95% uncertainty intervals from the beginning of the pandemic through the end of 2020.

**Figure 2.**
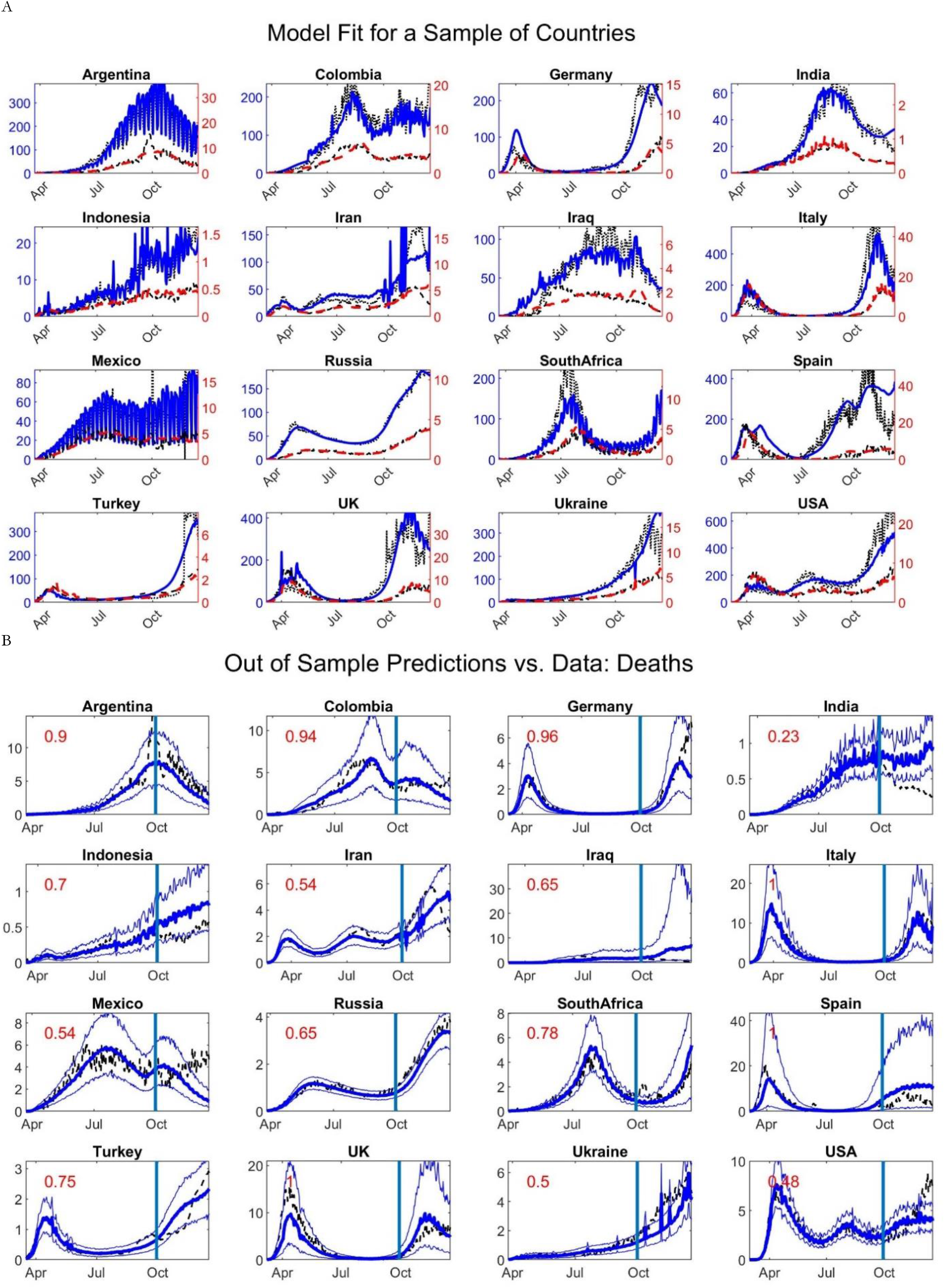
Model projections vs. data. A) Simulated (thick lines, Solid and Dashed for cases and deaths) vs. data (thin, dotted lines) for reported cases (top lines; right axis, thousands/day) and deaths (bottom lines; left axis; deaths/day) in countries with more than 45 million population and 300,000 confirmed cases by 22 December 2020 and testing data until at least September 2020. B) Out of sample death predictions for the same countries comparing data (dashed line) against estimates (thick solid line; 95% CIs are thin solid lines) using data until 30 September 2020 (denoted by vertical line). Numbers on each graph show the fraction of actual data during the prediction period inside the 95% confidence interval. High frequency noise (e.g. reported cases in Mexico) is due to weekly cycles in testing data.

Second, we assess out-of-sample accuracy. Figure *2*B shows out-of-sample prediction performance for reported deaths after fitting the model to data through 30 September 2020, then projecting outcomes through 22 December 2020. Across all predictions, 70% and 72% of observations for infections and deaths, respectively, fall within the 95% prediction intervals over the last 82-day period. Out-of-sample projections are limited by the fact that by 30 September 2020 the majority of countries had not experienced second waves or significant adherence fatigue, limiting the ability to estimate the magnitude of these behavioral feedbacks with the truncated sample. Despite these challenges, the model correctly predicts the existence of second waves in the majority of countries, and in many cases correctly predicts the timing and magnitude as well (see S6 for details and full results).

Third, we validated the estimation framework using synthetic data generated by simulating the model with known parameters and adding auto-correlated noise in infection rates and IFR. Our estimation procedure accurately identifies the vast majority of parameters in the synthetic dataset. The absolute error between the estimated and true values was significantly smaller than the estimated uncertainty (median error 21% of the 95% credible interval (CI), with 77% of the absolute errors less than half the 95% CI (see S3 for details).

Finally, we compare model estimates of actual cumulative cases against available national-level estimates from serological surveys. Few national seroprevalence studies include reliably representative samples. Nevertheless, using the SeroTracker project (Arora, Joseph et al. 2020) we identified nine country-level meta-estimates for actual prevalence. Figure *3* compares those meta-analytic estimates against official data from testing and model estimates. Seroprevalence and official counts vary by an order of magnitude or more. Model estimates are close for seven of the nine seroprevalence estimates and higher than surveys for Spain and Luxembourg. Note that seroprevalence data were not used in model specification or parameter estimation.

**Figure 3.**
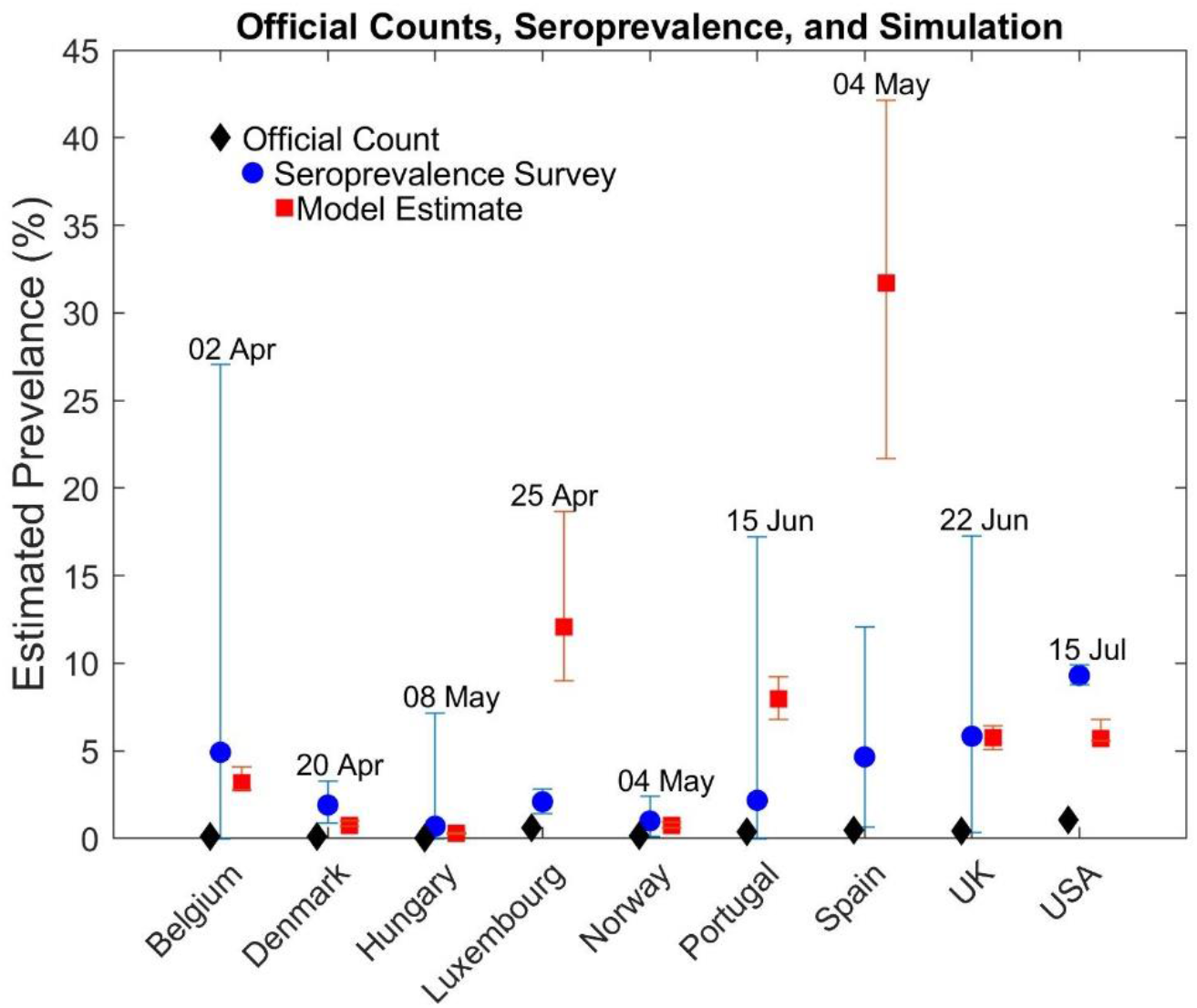
Comparison of cumulative percentage of cases based on official data (black diamond, left), seroprevalence estimates (blue circles with 95% CIs, middle), and model estimates (red squares with 95% CIs, right) for 9 countries at various dates.

## Results

### Quantifying under-reporting

We find COVID-19 prevalence and deaths are widely under-reported. Across the 92 nations, the estimated ratio of actual to reported cumulative cases (through 22 December 2020) is 7.03, corresponding to 465 million undetected cases (95% CI 478-513 million). Underreporting varies substantially across nations (10^th^-90^th^ percentile range 3.2-18; Figure *4*A).

Underreporting is due in part to the large fraction of asymptomatic infections, estimated at 50%, consistent with other estimates (Oran and Topol 2020). The mean inter-quartile range, MIQR, of the credible intervals in the national estimates is 1.3%. However, the estimated asymptomatic fraction varies little across nations (standard deviation, *σ* = 0.9%) and therefore cannot explain the large cross-national variation in the ratio of estimated to reported cases (Figure 4A).

**Figure 4.**
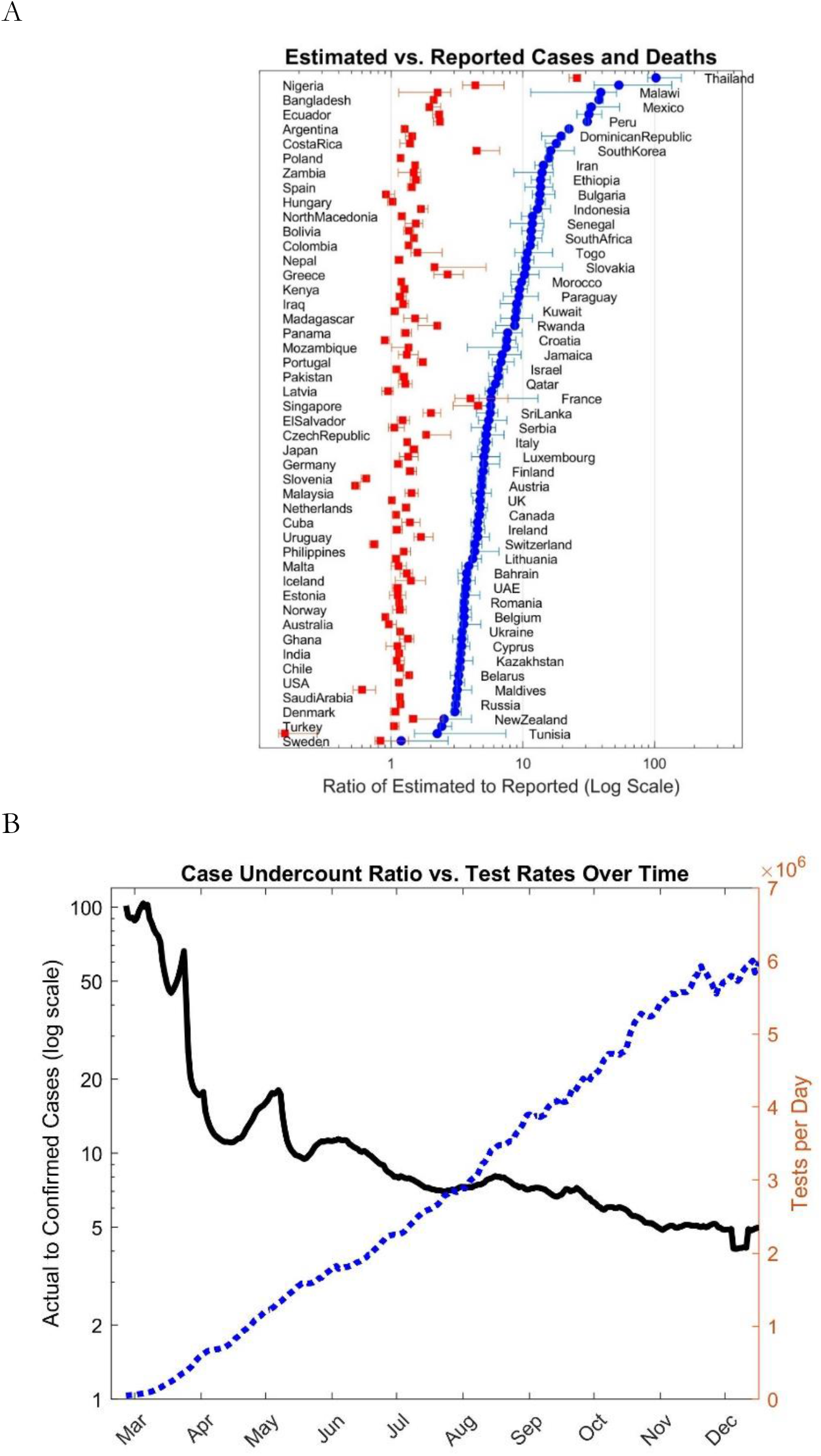
Underestimation of cases and deaths. A: Ratio of (estimated) actual to reported cases (blue circles) and deaths (red squares) by country; log scale as of 22 December 2020. B: Ratio of (estimated) actual to reported cases (solid line; left logarithmic axis) and total tests per day (dotted line; right axis) over time for the full sample (all 92 nations).

The extent of underestimation depends primarily on testing capacity and how it is utilized. If every person could be tested regularly the estimated ratio of actual to reported cases would be approximately 1.43, given assumed test sensitivity of ≈70% (Fang, Zhang et al. 2020, Wang, Xu et al. 2020). Testing capacity is limited, however. When testing capacity is small relative to the need, individuals presenting with COVID and COVID-like symptoms are prioritized, along with at-risk groups such as health care providers. Consequently, a larger proportion of those tested will be positive, but many infected individuals will go undetected, increasing the degree of underestimation, as seen in e.g. Mexico. Conversely, when testing capacity is high relative to the population, more of those infected will be identified, as seen in e.g. New Zealand. Over the full sample, increased testing has been continuously reducing the undercount ratio, though with decreasing returns (Figure 4B).

COVID-19 deaths are also underreported (Figure 4A). We estimate 2.07 (2.04-2.32) million deaths by 22 December 2020 across the 92 countries, 1.44 times larger than reported. Results are consistent with some country-specific estimates (Weinberger, Chen et al. 2020, Woolf, Chapman et al. 2020). Underreporting is significantly less for deaths than cases because deaths are concentrated among severe cases who are more likely to have been tested, and post-mortem testing corrects some of the undercount.

### Trends and Fluctuations in Cases and Mortality

National IFR estimates are reported in Figure 5A. IFR across the 92 nations through 22 December 2020 is 0.51% (CI: 0.47%-0.53%), with wide cross-national variation, from 0.04% (CI: 0.03%-0.04%; Qatar) to 1.99% (CI: 1.44%-2.21%; France), a range similar to estimates across counties in the USA (Basu 2020). These variations arise in the model from differences in age distribution (O’Driscoll, Dos Santos et al. 2020, Wu and McGoogan 2020) and the adequacy of health care. Consistent with prior estimates (Walker, Whittaker et al. 2020), we find hospitalization can reduce the age- and severity-adjusted risk of death to 46% of the rate without treatment, with large cross-national variation likely due to variations in treatment quality across nations (*σ*=22%; MIQR: 7%). Simulated IFR varies endogenously over time, exhibiting fluctuations around an overall declining trend (Figure 5B). The fluctuations are due to variations in the adequacy of treatment capacity, with IFR rising when surging caseloads overtake hospital capacity. The peak in global IFR in spring 2020 arose as cases overwhelmed hospital capacity in several nations, including many European countries with older populations. Since then, many—but not all—countries show notable reductions in IFR, due to factors including (i) improvements in treatments and greater availability of ventilators and PPE; (ii) heterogeneity in cases as some in the highest-risk populations were lost in the early waves; and (iii) heterogeneity in risk perceptions and responses as older, high-risk individuals adhere more strongly to NPIs compared those who perceive less personal risk, resulting in a decline in the average age of new confirmed cases in many nations. Available data do not enable us to identify the contribution of these different processes, but their combined effect is estimated to reduce IFR by 32.5% (*σ*=23.7%; MIQR=10.3%) for every doubling in cumulative cases. Despite the overall decline in IFR, hospital capacity shortages caused by renewed waves of infection increase IFR above the trend (e.g., India in June and July).

**Figure 5.**
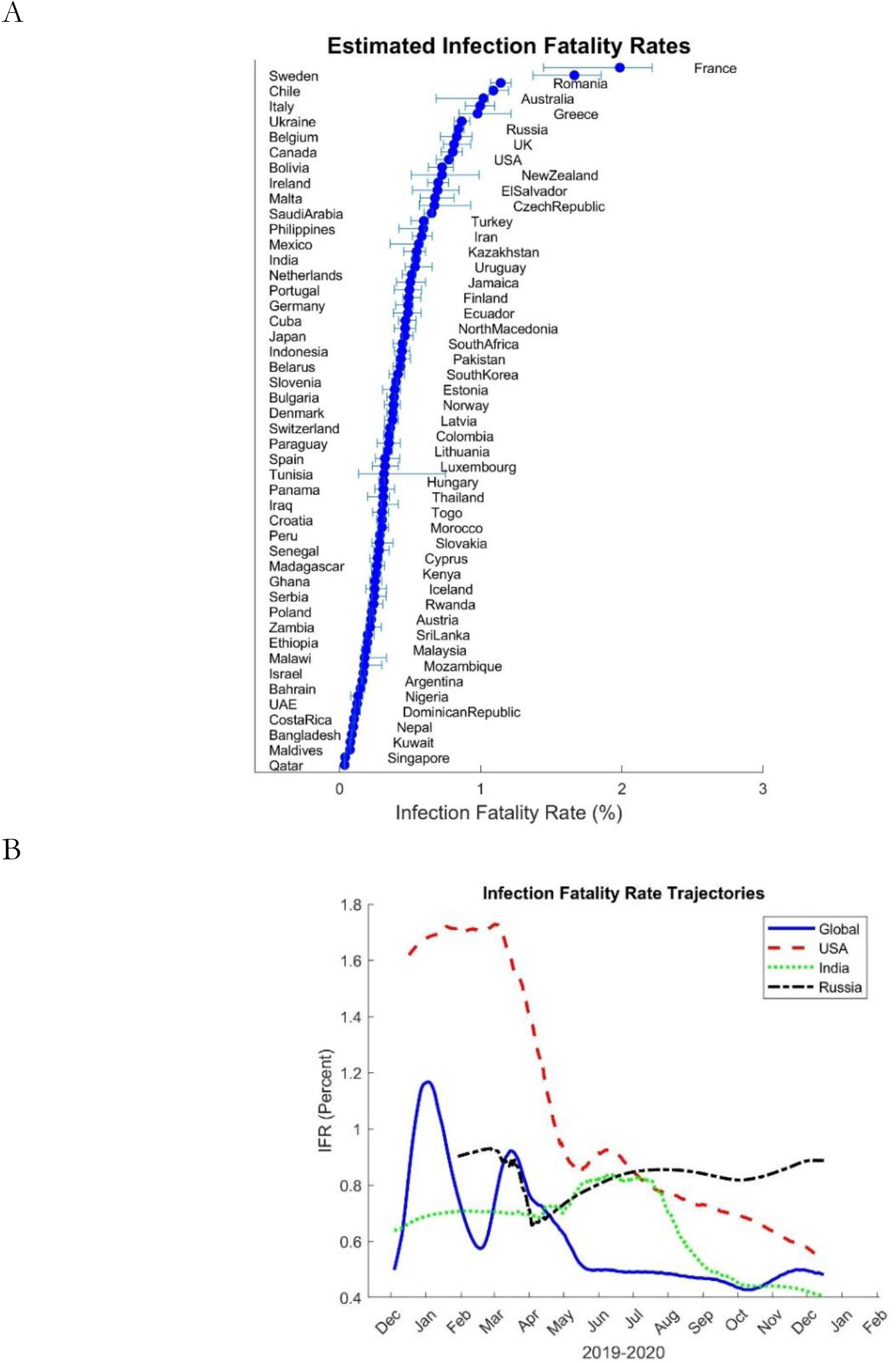
Fatality rates. A) Average estimated infection fatality rates (%) across countries as of 22 December 2020. B) IFR trajectories over time for the full sample and selected nations.

We also find significant heterogeneity across countries in the initial effective reproduction number, R_E_ (median 2.73, IQR 2.16-3.66), reflecting differences in population density, social networks, and cultural practices (Figure S9 provides details). We find a composite index of weather conditions by nation (Xu, Rahmandad et al. (2020) strongly affects transmission, generally increasing the hazard rate of transmission with the onset of winter.

Importantly, the effective reproduction rate, R_E_, changes over time as people and policymakers respond to perceived risk (Pan, Liu et al. 2020). We find behavioral and policy responses to the perceived risk of death reduce transmission and R_E_ with a mean lag of 38 days, though with substantial cross-national variation (*σ*=53.4; MIQR: 13.3). These responses are relaxed as perceived risk falls, though more slowly (mean lag 245 days; *σ*=188; MIQR: 64). Importantly, we also find that extended periods of contact reduction, and the personal and economic hardship it causes, lead to adherence fatigue—a reduction in the impact of perceived risk on behaviors that reduce contacts, with an estimated average elasticity of 1.24 (*σ*=1.07; MIQR: 0.19). A counter-factual simulation (Figure *6*, dotted line) shows that, absent adherence fatigue, total cases and deaths through 22 December 2020 would have been lower by 47% and 45%, respectively, corresponding to 265 (252-278) million cases and 1.18 (1.14-1.22) million deaths.

**Figure 6.**
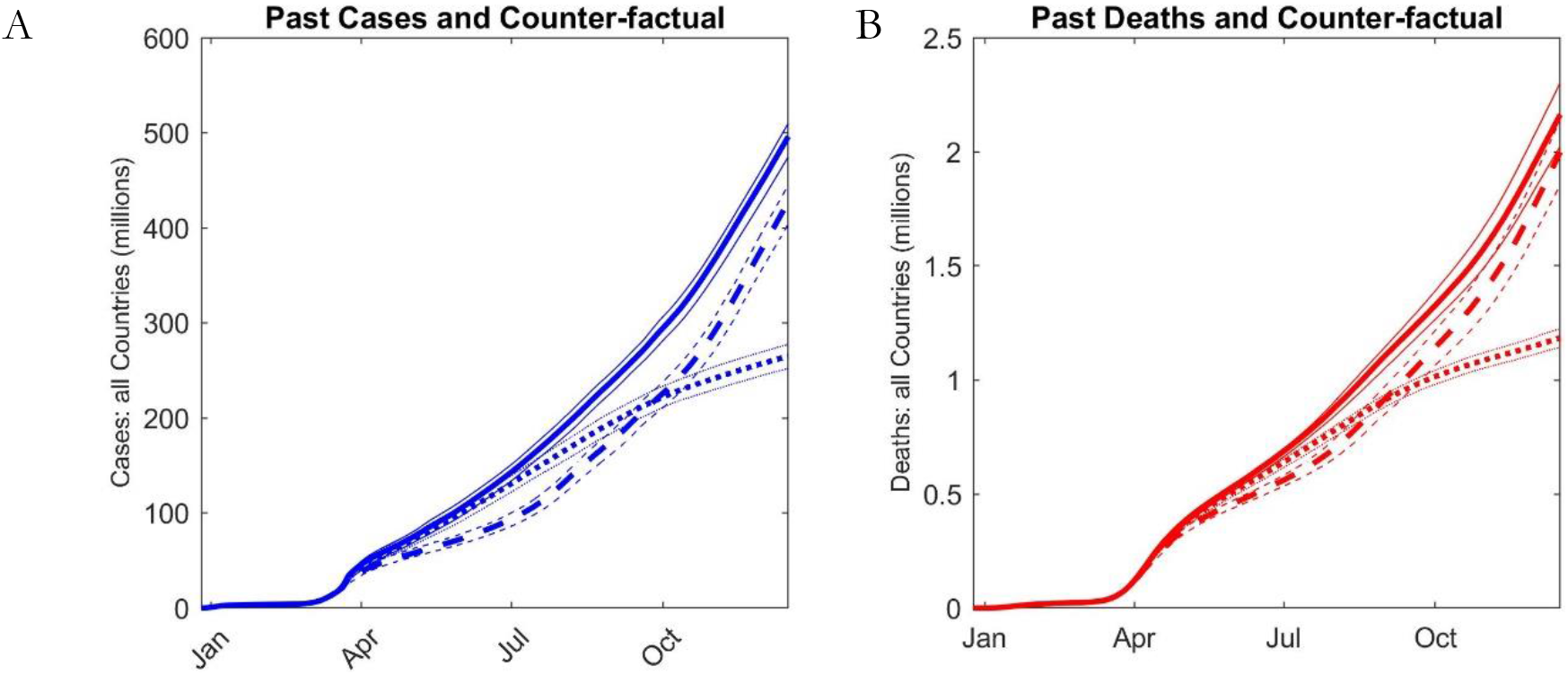
Counter-factual Experiments. Most likely estimate (solid lines) of cumulative cases (A) and deaths (B) compared with counterfactuals for scenario i, in which testing rises to 0.1% of population per day starting 11 March 2020 (dashed line); and scenario ii with no adherence fatigue (dotted line). Thin lines show 95% CIs.

The model endogenously captures the multiple waves observed across many countries through 22 December 2020 despite their different magnitudes and timing (Figure *2*A). These rebounds are due to lags in the perception of and responses to the risk of COVID-19. Initial reductions in transmission eventually lower deaths, leading to lower adherence to NPIs as perceived risk gradually falls, setting the stage for renewed outbreaks. Rebounds could also be triggered by changes in weather conditions, particularly the onset of colder weather. Rebound outbreaks are larger in nations where adherence fatigue is greater. Rebound waves are also larger where reductions in IFR are larger, because the decline in mortality erodes adherence to NPIs.

### Testing shapes early trajectories

Testing, treatment, risk perceptions, individual behavior, and government policy all change through several important feedbacks. We find that those receiving a positive test notably reduce their infectious contacts (Mean: 17.3% of original contact rate; *σ*=12.4%; MIQR: 9.9%). These reductions are especially important because symptomatic individuals, who are more likely to get tested, are estimated to be more infectious than asymptomatic ones (Mean asymptomatic infectiousness vs. symptomatic: 29.3%; *σ*=0.26%; MIQR: 1.3%; consistent with (Li, Pei et al. 2020)).

Testing also regulates the reported rate of deaths, which drives perceived risk. More reported deaths increase perceived risk, triggering behavioral and policy responses that reduce transmission. Plentiful early testing results in high detection rates, greater perceived risk, and stronger responses, slowing transmission. Conversely, insufficient early testing increases underestimation, limiting perceived risk and allowing transmission to further outpace testing. Testing thus reduces future cases, allowing a larger fraction of severe cases to be hospitalized, saving lives and reducing IFR. The exponential nature of contagion means even small early differences can lead to notable differences in epidemic size (Pei, Kandula et al. 2020), IFR, and total deaths.

Figure *6* (dashed line) shows the impact of enhanced early testing by comparing the estimated results to a counterfactual in which all countries test 0.1% of their population per day, a rate currently exceeded over the full sample (See Figure *3*B). We assume enhanced testing begins when WHO declared COVID-19 a pandemic (11 March 2020). We find enhanced testing would have reduced total cases from 496 million to 427 (CI: 404-445) million, with a reduction in deaths from 2.16 to 2.00 million (CI: 1.85-2.14).

### Projections with endogenous responses to risk and vaccination

We explore four scenarios projecting the pandemic through 30 June 2021 (Figure *7*): (I) *Baseline*: assumes country-specific testing rates continue as of 22 December 2020 with country-specific estimated parameters. (II) *Responsive*: countries become 20% more responsive to risk (details in supplement S5). Scenario II represents a modest increase in responsiveness given levels of perceived risk: on average, contact rates fall by 7% (σ=17%) January-June 2021, comparable to the impact of enhanced mask use (Chu, Akl et al. 2020). (III) Vaccination: Assumes every country will be fully vaccinated by the end of 2021 using a constant vaccination rate. This timeline is optimistic for developing countries; furthermore, the vaccine is assumed to be perfectly protective, to face no resistance, and to be administered with priority to high-risk individuals. (IV) Enhanced responsiveness and vaccination, combining Scenarios II and III.

**Figure 7.**
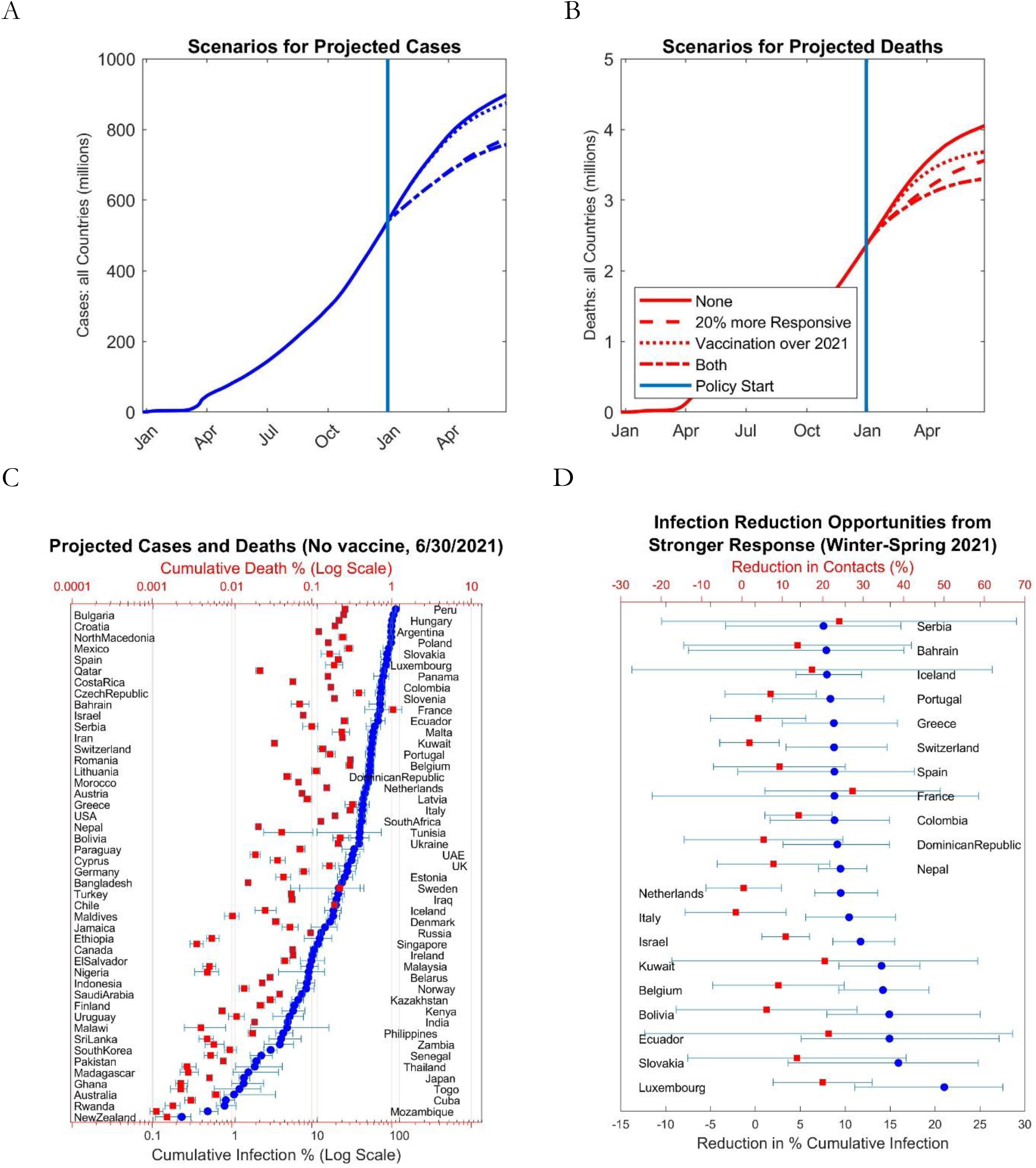
Projections across four scenarios. Estimated true cumulative cases (A) and deaths (B) across 92 countries until June 30 2021 under scenarios I (No vaccine, same responsiveness; solid line), II (20% more responsive; dashed), III (vaccinating population over 2021; dotted), and IV (responsiveness and vaccination, II+III; dash-dot). C) Median (95% CI) estimated true cumulative cases (blue circels; bottom axis) and deaths (red squares; top axis) as % of population, projected by the end of June 2021 under Scenario I (Logarithmic Scale). D) Median (95% CI) reductions in cumulative cases in Scenario II (20% more responsive) compared with Scenario I (in % of population) for top 20 countries (blue circles; bottom scale); red squares (top scale) represent average change in contacts compared to Scenario I for January- June 2021 period.

Figure *7* contrasts the results. Scenario I yields 900 million cumulative cases by 30 June 2021. Most cases are concentrated in a few countries, with USA (115 million median cumulative cases; 95% CI 108-122 million), Mexico (96 million; 82-105), India (60 million; 57-65), and Iran (40 million; 35-44) suffering the largest burdens. Figure *7*A & B show estimated cumulative actual cases and deaths by 30 June 2021 (Supplement S5 provides Scenario I projections over time). The responsive scenario (II) reduces cases (Figure *7*A) and deaths (Figure *7*B) by 14% (CI: 12%-16%) and 12% (CI: 7%-17%). The largest reductions arise in countries with large populations and that are estimated to have a large potential for new waves (Figure *7*D), including India (12.3 million reduction in cases), USA (9.51 million), Mexico (7.48 million), and Italy (6.3 million) among others. The vaccination scenario brings down cases and deaths by 2.7% and 9.1%, to 875 million (CI: 851-898) and 3.68 million (CI: 3.45-3.90) respectively, less than the impact of greater responsiveness to risk. Combining the two provides additional benefit but with decreasing returns for cases (16%; CI: 13%-18%). The relatively small impact of vaccination through mid 2021 is due to two factors. First, by mid 2021 we assume approximately half the population is immunized (but with a vaccine assumed to be 100% protective). Second, behavioral feedbacks undermine the impact of immunization: to the extent immunization reduces deaths and perceived risk, adherence to NPIs erodes, leading to more new cases.

These scenarios should not be interpreted as predictions: changes in testing, vaccination, individual behavior, and government responses to risk not accounted for in the model are possible, perhaps likely, in the coming months. The general finding is the significant sensitivity of outcomes to behavioral and policy responses to risk. Stronger responses to perceived risk would significantly reduce future cases. Lax responses and greater adherence fatigue would lead to larger rebound outbreaks. Moreover, responsiveness comes at a low cost in terms of cumulative reductions in contacts, a proxy for the reductions in travel, dining, shopping, and other activities that lead to unemployment and harm businesses. We next explore this important finding in more depth.

### A global dilemma: similar behaviors, different outcomes

Across scenarios, a few nations are projected to experience significant growth in incidence before vaccines become widely available, but most are able to stabilize their epidemics through NPIs, albeit with occasional new outbreak waves. Endogenous risk perceptions create an important balancing (negative) feedback that leads most countries to converge to an effective reproduction number R_E_ ≈ 1: R_E_ > 1 leads to rapid growth in cases and deaths, increasing perceived risk and renewed use of NPIs that bring R_E_ down; R_E_ < 1 lowers cases and deaths, leading to the erosion of perceived risk and reduced adherence to NPIs that then lead to more cases, raising R_E_ back toward one.

Critically, however, R_E_ fluctuates around 1 at very different quasi-steady state infection and fatality rates across nations. Recall that R_E_ ≈ 1 means that, on average, infected individuals infect one new case before they are removed from the infectious pool by recovery or death. That balance can be achieved at a high or low level of prevalence. Those nations with high responsiveness to risk settle at R_E_ ≈ 1 with low prevalence and death rates, while those with lower responsiveness to perceived risk achieve that balance only when cases and deaths rise high enough to drive R_E_ down toward 1. The large cross-national variation in the estimated behavioral responses to risk lead to death rates more than two orders of magnitude higher in nations with weak responsiveness and greater adherence fatigue compared to those with sustained strong responses (*Figure 8*). Over the 6 months preceding 22 December 2020, average estimated R_E_ values have approximately converged to ≈1 (mean=1.19, σ=0.21), indicating comparable levels of contact reduction across nations. In contrast, deaths per million over the same period show a 10-90th percentile range of 0.033 to 3.7.

**Figure 8.**
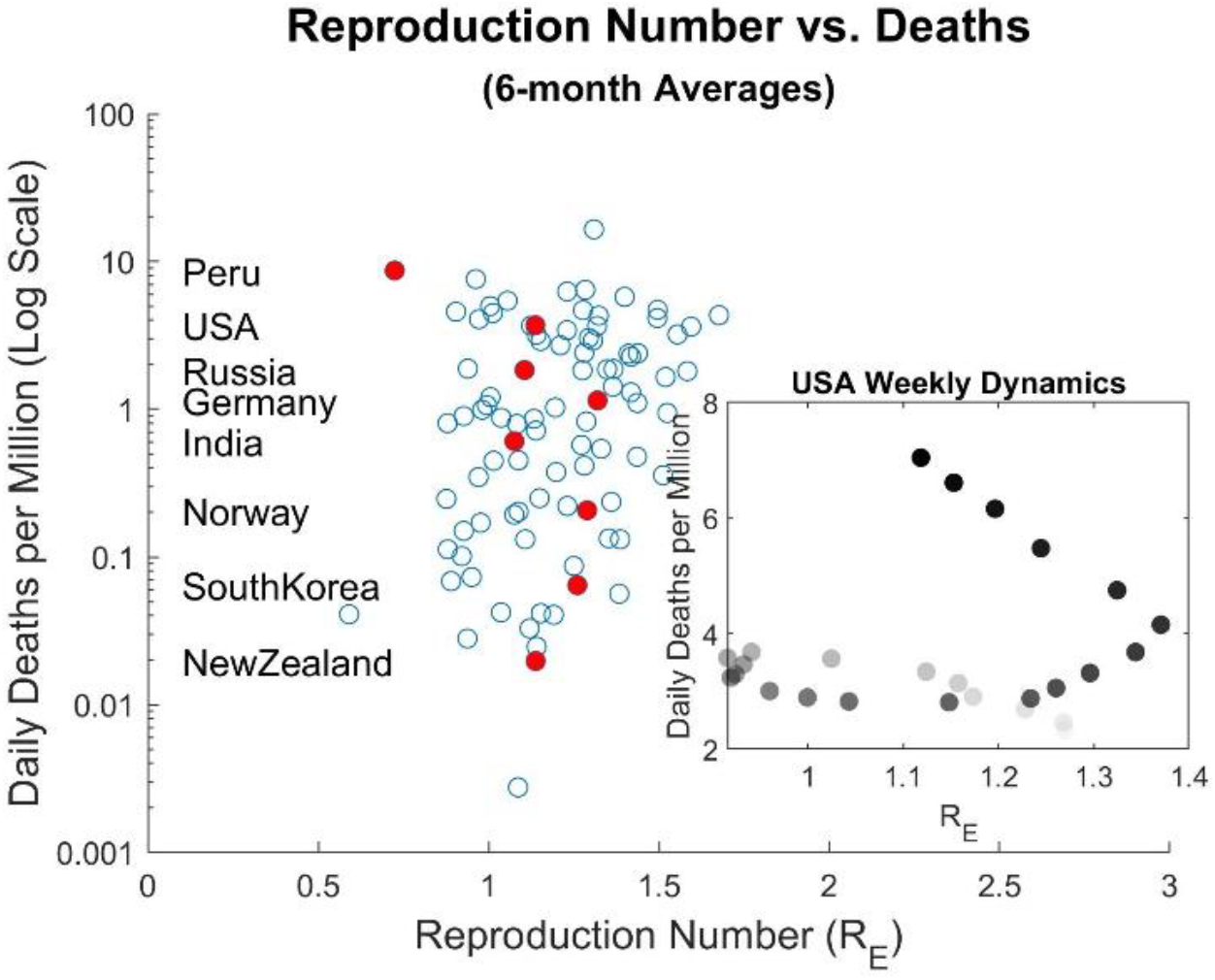
Effective reproduction number RE vs. estimated true daily deaths per million in each nation, averaged over the 6 months ending 22 December 2020, with a few countries highlighted. Inset: Oscillation in death rate vs. RE in the USA caused by lags in the perception of and behavioral responses to the risk of death, and lags between behavioral responses that reduce transmission and subsequent deaths. Data are weekly averages over the six-month period ending 22 December 2020 (darker circles are the more recent weeks).

### Summary of main results

We find prevalence and mortality are substantially under-reported by official data: across the 92 nations for which data are available, estimated cumulative COVID cases are approximately 7 times greater than official reports, with under-reporting across nations spanning three orders of magnitude. The magnitude of under-reporting has declining over time as testing has increased. Nevertheless, and despite the large magnitude of under-reporting, estimated cumulative cases through the end of 2020 constitute small fractions of the populations, so herd immunity remains distant in nearly all nations (see S5).

We find deaths are 1.44 times larger than official reports. The overall infection fatality rate to date is ≈0.51%, consistent with growing evidence (Russell, Hellewell et al. 2020, Verity, Okell et al. 2020). We also find substantial cross-national variation in IFR. The variation arises from differences in population age structure, and in the burden of severe cases relative to hospital capacity, highlighting the importance of limiting case growth. We also find that IFR has, on average, declined substantially since the onset of the pandemic. Despite the overall declining trend in IFR, mortality surges when renewed outbreaks overwhelm treatment capacity.

We estimate that approximately half of infections are asymptomatic, consistent with estimates from smaller samples (Gudbjartsson, Helgason et al. 2020, Lavezzo, Franchin et al. 2020, Mizumoto, Kagaya et al. 2020), with asymptomatic individuals estimated to be about one-third as infective as symptomatic patients.

The wide variation across nations arises endogenously and without major differences in biological parameters. First, inadequate early testing in some countries has led to greater underestimation of prevalence, and thus later and weaker responses, causing faster epidemic growth, further outstripping testing and treatment capacity in a self-reinforcing (positive) feedback. Second, the growth rate, timing, and size of outbreak waves depend strongly on the magnitude of behavioral and policy responses to perceived risk, and the lags in forming and eroding those perceptions, which we find vary notably across countries and determine the gain and phase lag in the negative feedbacks that lead to outbreak waves. These feedbacks amplify minor differences in testing and responsiveness to perceived risk, generating significant heterogeneity in cases and deaths despite convergence of R_E_ to ≈1 across nations. Projections through 30 June 2021 show that modest reductions in transmission through NPIs can lead to large reductions in cumulative cases and deaths in many countries, even absent effective vaccines. In contrast, the early impact of vaccines is modest, because risk-driven response leads to relaxation of NPIs and keeps cases and deaths high until a large fraction of the population is vaccinated.

Due to the rapidly evolving nature of the pandemic, behavior and policies, and vaccine availability and uptake, we have developed and will regularly update an online simulator enabling users to explore alternative vaccination and responsiveness scenarios over the coming months, freely available at https://exchange.iseesystems.com/public/mitsdl/covidglobal/.

### Robustness and boundary conditions

We conducted several analyses to assess the robustness of the results. (1) We ensured MCMC chains had converged (100% of Gelman-Rubin convergence statistics were below 1.2 and 97% under 1.1). (2) We varied the priors for cross-country parameter variances to 4 and 0.25 times the base values, resulting in <3% change across historical measures, though scenario results for a few countries are more sensitive (S7 provides details). (3) To see if any country disproportionately affects the results we repeated the analysis using three different samples, excluding the top 5 countries by (i) estimated cases, (ii) reported cases, and (iii) population. Only India and USA appear in all three sets. Average outcomes across the remaining countries changed less than 1% (see S7). (4) We assessed the sensitivity of results to the parameters estimated from prior research by calculating the elasticities of key outcomes to parametric assumptions. Across metrics most elasticities are less than approximately 0.5. The largest is for the impact of test sensitivity; greater sensitivity reduces under-estimation.

These robustness tests do not address a few limitations that temper the interpretation of the results. Some arise from limits on data availability across nations. For example, we are unable to include China and Brazil because they do not report adequate testing data, and other useful data, such as all-cause mortality, are not available for some nations. Some limitations arise from the computational burden of estimation: although we limit the number of parameters to be estimated to avoid overfitting, the analyses we report, including estimation of credible intervals via MCMC, take 2 weeks of continuous parallelized computation on a 48-core server.

Other limitations are due to inherent uncertainties. First, we do not represent within-nation heterogeneity that may affect the course of the epidemic. These variations likely matter especially in large, diverse nations (e.g., USA), including differences in transmission risk between rural and urban areas, differences in adherence to NPIs based on political views, and especially differences in the ability of individuals and households to limit transmission risk or receive treatment based on socio-economic status, race and ethnicity, and other factors affecting social justice (Britton, Ball et al. 2020, Laxminarayan, Wahl et al. 2020, Painter and Qiu 2020). Second, we model IFR as depending on age distribution and hospitalization, but do not explicitly model how well different nations are able to protect vulnerable subpopulations (the effect is aggregated into the overall impact of cumulative cases on IFR). Third, the model aggregates behavioral and government policy responses, and does not represent the effectiveness of specific NPIs. Fourth, due to the computational burden it would impose, we did not use filtering (e.g. Kalman or particle) or state-resetting methods to account for time-varying determinants of transmission (e.g. the impact of holidays). Fifth, we do not account for emergence of new variants of the virus that may alter transmissibility or IFR. Finally, without explicit travel networks our results may under-estimate the risk of re-introduction of the disease where it has been contained.

## Discussion

The study yields conceptual, methodological, and policy implications. Conceptually, the model we develop integrates the biological and social factors conditioning transmission captured in typical SEIR models with a range of behavioral factors, some of which are novel in epidemiological modeling. We include endogenous test allocation and hospitalization based on symptom severity relative to capacity; endogenous risk perceptions and responses, including adherence fatigue; and learning and other factors that cause the infection fatality rate to decline on average over time. While these mechanisms are intuitively plausible, empirical estimation of each feedback loop is indispensable for selecting which mechanisms to include in the model—and which to leave out; inferring the actual toll of COVID-19; explaining the multiple waves of infection; offering reliable long-term projections; and assessing the likely impact of policies. Quantification also enables us to explain orders of magnitude variations in outcomes across nations without resorting to exogenous time-varying parameters or ad-hoc nation-specific fixed effects, and with parameters characterizing COVID-19 that are consistent with prior research.

Two methodological contributions may inform future work. Our modeling framework captures heterogeneity in disease severity, which conditions test and treatment capacity allocation, without the need for explicit disaggregation into subpopulations or to the individual level. Our approach makes estimation computationally feasible and provides a consistent method to model the allocation of testing, hospital capacity, and the impact of acuity on mortality. Second, our hierarchical Bayesian estimation framework enables us to use the data from all nations to inform the estimated parameters for each. As expected, we find parameters capturing biological attributes of SARS-CoV-2 and COVID-19, such as the asymptomatic fraction of cases, have far less cross-national variation than parameters characterizing risk perceptions and behavioral responses to the threat.

A counter-intuitive finding provides important policy implications. After controlling the initial peak, most countries have settled into a quasi-steady state with the effective reproduction number R_E_ fluctuating around one, but with caseloads and death rates varying by two orders of magnitude. This result arises directly from the endogenous inclusion of behavioral feedbacks: lower mortality erodes adherence to NPIs, raising R_E_ and leading to rebound outbreaks, which then lead to renewed contact reductions that bring R_E_ back down. However, the estimated responsiveness to risk varies widely across nations. Those with strong responses bring R_E_ to 1 with few cases and deaths, while those with weak responses require much larger death rates to drive R_E_ toward one. Critically, vaccine introduction does not change this fundamental result: vaccines save lives, triggering the relaxation of NPIs. The initial reduction in death rates will therefore be slower than expected based purely on the protection offered by vaccines. Consequently, different nations pay widely different prices in lost lives. Although we do not carry out a detailed analysis of the economic costs of different NPIs, the behavioral and policy changes that reduce contacts enough to bring R_E_ to one are a rough measure of the self-isolation, distancing, and other actions that reduce economic activity and employment by cutting travel, dining, shopping, and other activities sustaining commerce and industry. Thus, by increasing responsiveness to risks, communities and nations can bring down death rates at little additional economic cost, a finding consistent with analysis of the 1918 influenza pandemic (Correia, Luck et al. 2020). Although contrary to the intuitions of many policy makers, the results suggest no strong tradeoff between saving lives and saving the economy. Stronger responsiveness to risk and adherence to NPIs offer an opportunity to save lives at low costs even as vaccines are approved and deployed.

## Data Availability

All data, codes, and simulation models are publicly available.

https://github.com/tseyanglim/CovidGlobal

## Supplementary Materials

### S1 MODEL STRUCTURE AND KEY FORMULATIONS

The model simulates the evolution of COVID-19 epidemic, risk perception and response, testing, hospitalization, and fatality at the level of a country, and couples all countries in the parameter estimation step. Here we explain key equations and structures in each sector, followed by complete listing of model equations and parameters in S8. Full model, data, and analysis code is available online at https://github.com/tseyanglim/CovidGlobal.

#### Population Groups and Transmission Dynamics

The model is a derivative of the well-known SEIR (Susceptible, Exposed, Infectious, Recovered) framework for simulating infection dynamics. Figure S1 provides an overview of key population groups and the population movements among them^1^.

**Figure S1.**
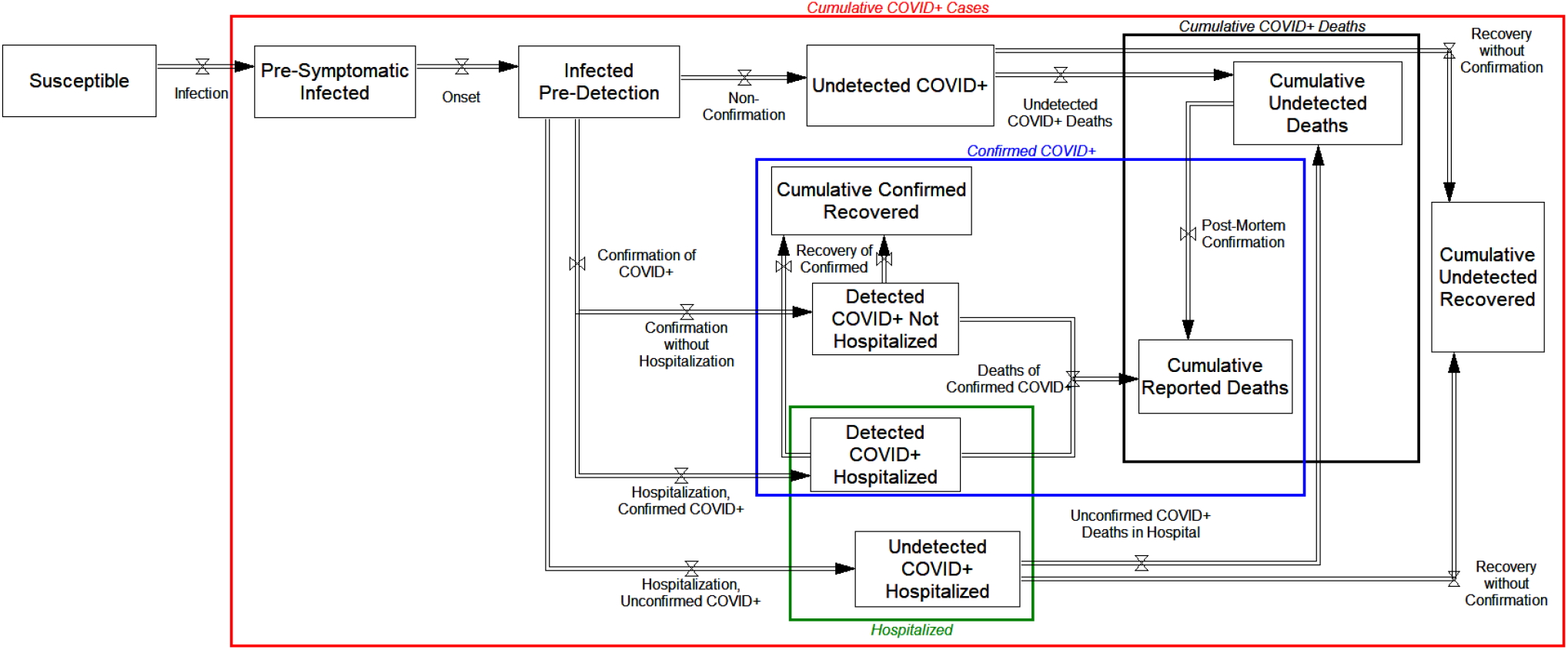
Key population stocks and flows. Rectangles represent stocks (state variables), while arrows and valves represent the flows between them (state transitions). Some in the *Susceptible* population (S) flow into the *Pre-Symptomatic Infected* stock (P) based on the *Infection Rate* (*r*_*SP*_). After an average *Incubation Period* (τ_P_), these pre-symptomatic infected flow into the *Infected Pre-Detection* (*I*_*P*_) stock. After a further average *Onset to Detection Delay* (*τ*_*T*_), this group splits among multiple pathways. First, if tested positive for COVID-19, they flow into either *Infectious Confirmed Not Hospitalized* (*I*_*C*_) or *Hospitalized Infectious Confirmed* (*I*_*CH*_). Anyone not tested positive, whether for lack of testing or erroneous test results, transitions into either *Hospitalized Infectious Unconfirmed* (*I*_*UH*_) or *Infectious Unconfirmed Post-Detection* (*I*_*U*_). We assume demand for testing and hospitalization are driven by symptoms, so all asymptomatic patients will be in the latter category.

From these Infectious categories, resolution flows (*r*_*…*_) take individuals to either Recovered (R_…_) or Dead (D_…_) states, with corresponding subscripts _*U, C, CH*_, and _*UH*_ for stocks and _*UU, UHCH*_ etc. for flows. Given the differences in severity and potential survival extension due to hospitalization, we distinguish between resolution delay for those in hospital (*Hospitalized Resolution Time; τ*_*H*_) and those not hospitalized (*Post-Detection Phase Resolution Time*; *τ*_*R*_). We use first order exponential delays for all lags, though sensitivity analyses showed very little impact of using higher order delays.

The *Infection Rate* (*r*_*SP*_) controls the flow from S to P and depends on *Infectious Contacts* (*C*_*I*_), fraction of total *Population* (*N*) that is susceptible, and *Weather Effect on Transmission* (W). The latter is a function of *R*_*W*_, the country-level projections for impact of weather on COVID-19 transmission risk year-round developed by Xu and colleagues (*1*) and a parameter, *Sensitivity to Weather* (*s*_*W*_), to be estimated:

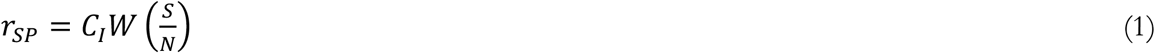

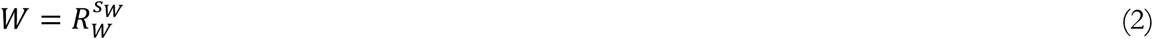

Infectious contacts depend on the *Reference Force of Infection* (β), various infectious sub-populations (and their relative transmission rates; *m*_*a*_ for asymptomatic and *m*_*T*_ for confirmed), and *Contacts Relative to Normal* (*F*_*C*_), which captures behavioral and policy responses as a fractional multiplier to baseline infectious contacts:

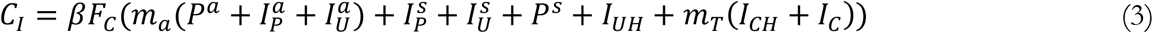

In this equation we separate various stocks (of I and P) into asymptomatic (a superscript) and symptomatic (s superscript). That distinction is treated analytically using a zero-inflated Poisson distribution that is discussed in the next section. In light of evidence on the short serial interval for COVID-19, likely below the incubation period (*2, 3*), we do not distinguish the infectivity of pre-symptomatic individuals from those post onset. Contagion dynamics start from *Patient Zero Arrival Time, T*_*0*_, another estimated parameter. The key mechanisms regulating the population flows among these stocks are discussed below, and a schematic of important relationships is provided in Figure S2.

Five parameters are estimated in the equations discussed above. One of them (s_W_) is global (i.e. assumed identical across countries; see the estimation section below for details on the distinction between global and country-specific parameters) and the remaining four are country-specific: *β, m*_*T*_, *m*_*a*_, *and T*_*0*_.

**Figure S2.**
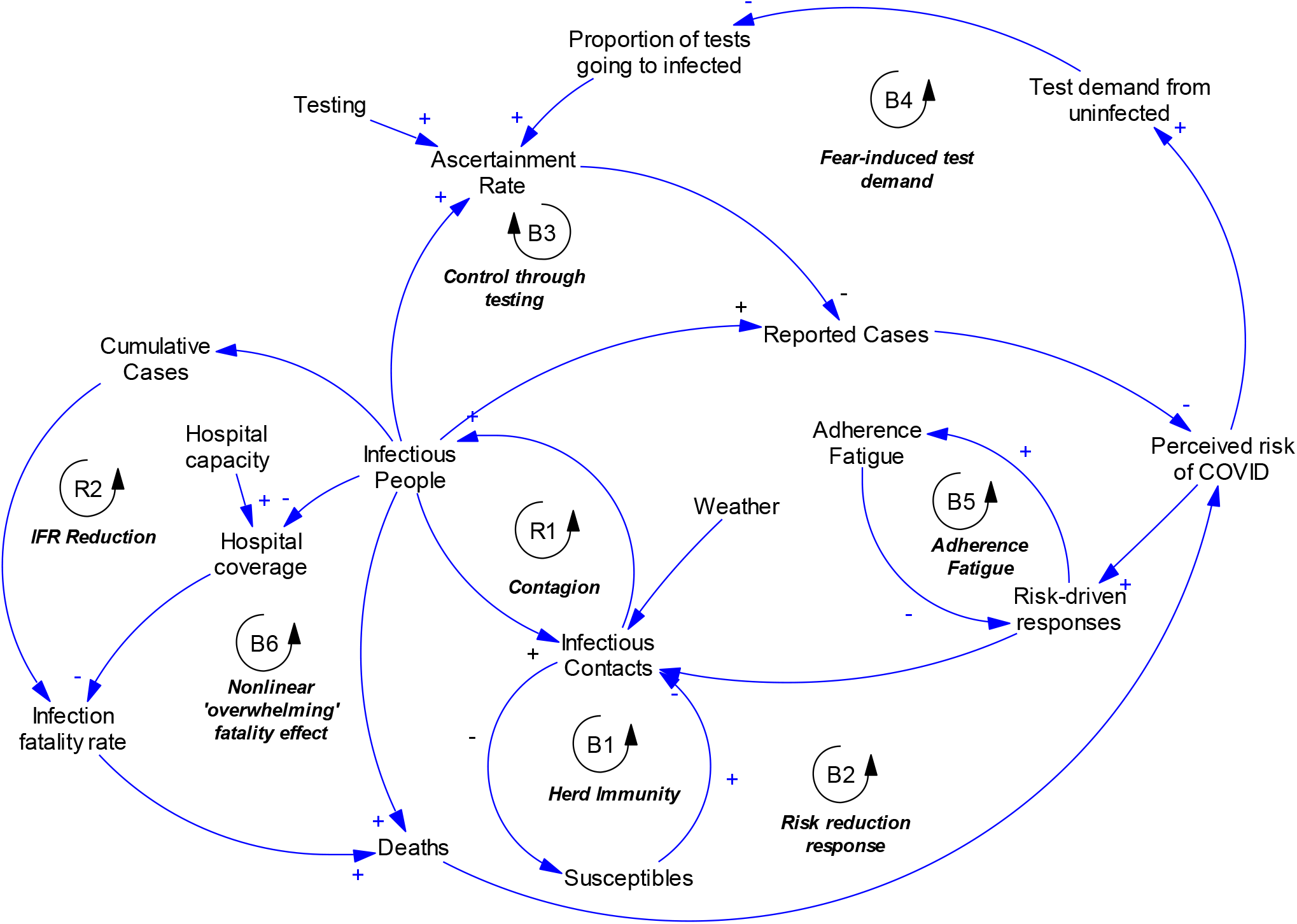
Overview of model’s mechanisms. Major feedback loops are identified as Balancing (Negative feedback; B) and Reinforcing (Positive feedback; R).

#### Modeling the Severity of Symptoms

COVID-19 infection varies in acuity, from asymptomatic to life-threatening. Disease acuity affects fatality risk and also testing and hospitalization decisions, which in turn affect official records of infection and fatality rates. Since movement between population groups via testing or hospitalization is itself a function of acuity, to allow for consistent inference of mean acuity across different population groups, we use an analytical framework to track acuity levels. The framework, which we adapted from prior research (*4*), obviates the need to disaggregate the population by different acuity levels (which would prohibitively raise the computational costs for estimation).

Specifically, we represent acuity using a zero-inflated Poisson distribution. This distribution combines two subpopulations – one with Poisson-distributed acuity levels with mean *Covid Acuity* (α_C_), and another *Additional Asymptomatic Fraction* with zero acuity, which is the zero-inflated component. The sum of those with zero acuity from the Poisson part of the population and the second group is the *Total Asymptomatic Fraction* (*p*_*a*_). We assume this asymptomatic group is not given priority in testing or hospitalization, and is not at risk of death. Thus they will always follow the 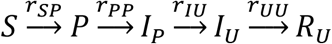 pathway. The pathways for the remaining population depend on acuity and its impacts on testing, hospitalization, and death. Note that the concept of acuity defined here only needs to have a monotonic relationship with tangible symptoms and risk factors and it does not have a one-to-one relationship with any real-world measure of acuity, and as such is better seen as a mathematical construct that informs modeling rather than a real-world variable with clinical definition.

From this framework two parameters, *a* and *α*_*C*_, are estimated as country specific parameters with limited variability across countries.

#### Testing

The testing sector reads the *Active Test Rate* (*T*_*t*_) for each country as exogenous input data (see appendix S3 for pre-processing details for this data). A fraction of the total test rate, typically small, is allocated to post-mortem testing of COVID-19 victims who have not been previously confirmed (*Post Mortem Tests Total, T*_*PM*_). Specifically, of the deaths of unconfirmed infectious individuals (whether hospitalized or not), a certain *Fraction of Fatalities Screened Post Mortem* (*n*_*PM*_) will be identified true post-mortem tests. We anchor the *n*_*PM*_ to *Fraction Covid Death In Hospitals Previously Tested* (*n*_*DCH*_). The rationale for this anchoring is that on the margin if there are many unidentified COVID patients in hospitals, the chances are that the system lacks enough testing capacity and thus post-mortem testing should also be less thorough:

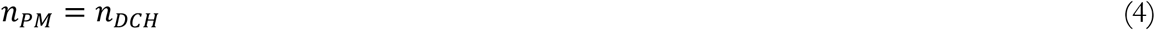

We experimented other functional forms with a free parameter connecting the two constructs, but following our conservative estimation principle decided against including that free parameter in the final model. We feared that absent clear observables to identify this additional parameter (e.g. on country-specific policies regulating post-mortem testing) the degree of freedom would improve the fit but potentially for the wrong reason.

The remaining *Testing Capacity Net of Post Mortem Tests* (*T*_*Net*_ = *T*_*t*_ −*T*_*PM*_) is allocated to test demand from two sources. First, symptomatic COVID patients leaving the pre-detection (*I*_*P*_) phase may seek testing (*Positive Candidates Interested in Testing Poisson Subset*: 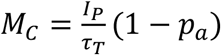). Second, COVID-negative individuals may seek testing due to various perceived risks and other conditions with overlapping symptoms such as common cold and influenza-like illnesses (*M*_*N*_, *Potential Test Demand from Susceptible Population*). This “negative” demand includes a *Baseline Daily Fraction Susceptible Seeking Tests* (*n*_*ST*_) of the population not previously tested positively (*N*_*U*_), and increases with the *Recent Detected Infections* (*T*_*PIR*_), which is an exponentially weighted moving average of *Positive Tests of Infected* (*T*_*PI*_). COVID-positive and COVID-negative sources of demand add up to create the overall *Testing Demand* (*M*_*T*_):

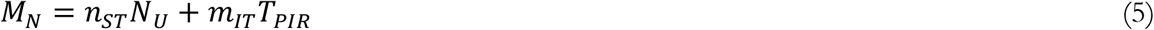

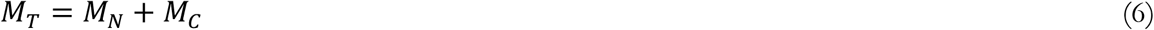

Where the *Multiplier Recent Infections to Test* (*m*_*IT*_), captures the sensitivity of negative test demand to recent infection reports.

To allocate the available tests (*T*_*Net*_) between these two sources of demand, we use an analytical logic that allocates testing based on symptom severity. Via self-selection and screening by testing centers, people who have more symptoms or other signals that correlate with COVID infection (e.g., high exposure risk) are more likely to be tested. We assume each unit of acuity increases the likelihood that an individual gets tested, based on a variable *Prob Missing Symptom, p*_*MS*_. This variable represents the probability that each acuity unit fails to convince the testing decision process to test a given individual, i.e. how selectively and sparingly tests are conducted. Specifically, in this model an individual with *k* acuity units is tested with probability:

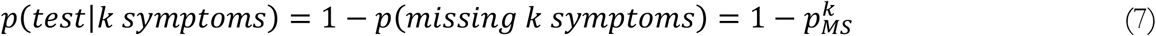

We assume the negative test demand is coming from a population with a Poisson-distributed, unit average acuity level (α_*N*_=1) for symptoms of non-COVID influenza-like illnesses. The test demand from COVID patients also comes from a Poisson distribution of acuity, but with mean α_C_. With the Poisson distribution and given a level of α and *p*_*MS*_, one can calculate the fraction of each demand source that would be tested:

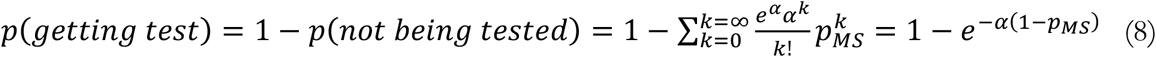

We therefore need to find the *p*_*MS*_ that allows test supply to match demand that is satisfied, specifically, by solving the following equation for *p*_*MS*_*\**:

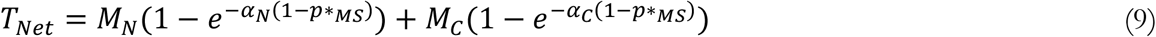

Figure S3 provides a graphical summary of the zero-inflated Poisson symptom and testing framework. In this figure testing outcomes are graphed for a population where 10% are COVID-positive, assuming that *Covid Acuity*, α_C_, is 6, and with two different levels of *p*_*MS*_ (=0.8 and 0.95). For this figure we also assume a 55% asymptomatic fraction for COVID patients. Even with testing that prioritizes patients with more symptoms, and despite the large difference in symptom frequency between COVID patients and negative cases, the majority of tests are allocated to negative cases with a few symptoms. COVID patients with multiple symptoms are likely to be identified if P_MS_ is not very large, but when total demand for testing (i.e. the sum of all bars with symptoms>0) is large, P_MS_, found from solving equation 9, may be close to 1, excluding many COVID patients with multiple symptoms and thus higher risks of fatality.

**Figure S3.**
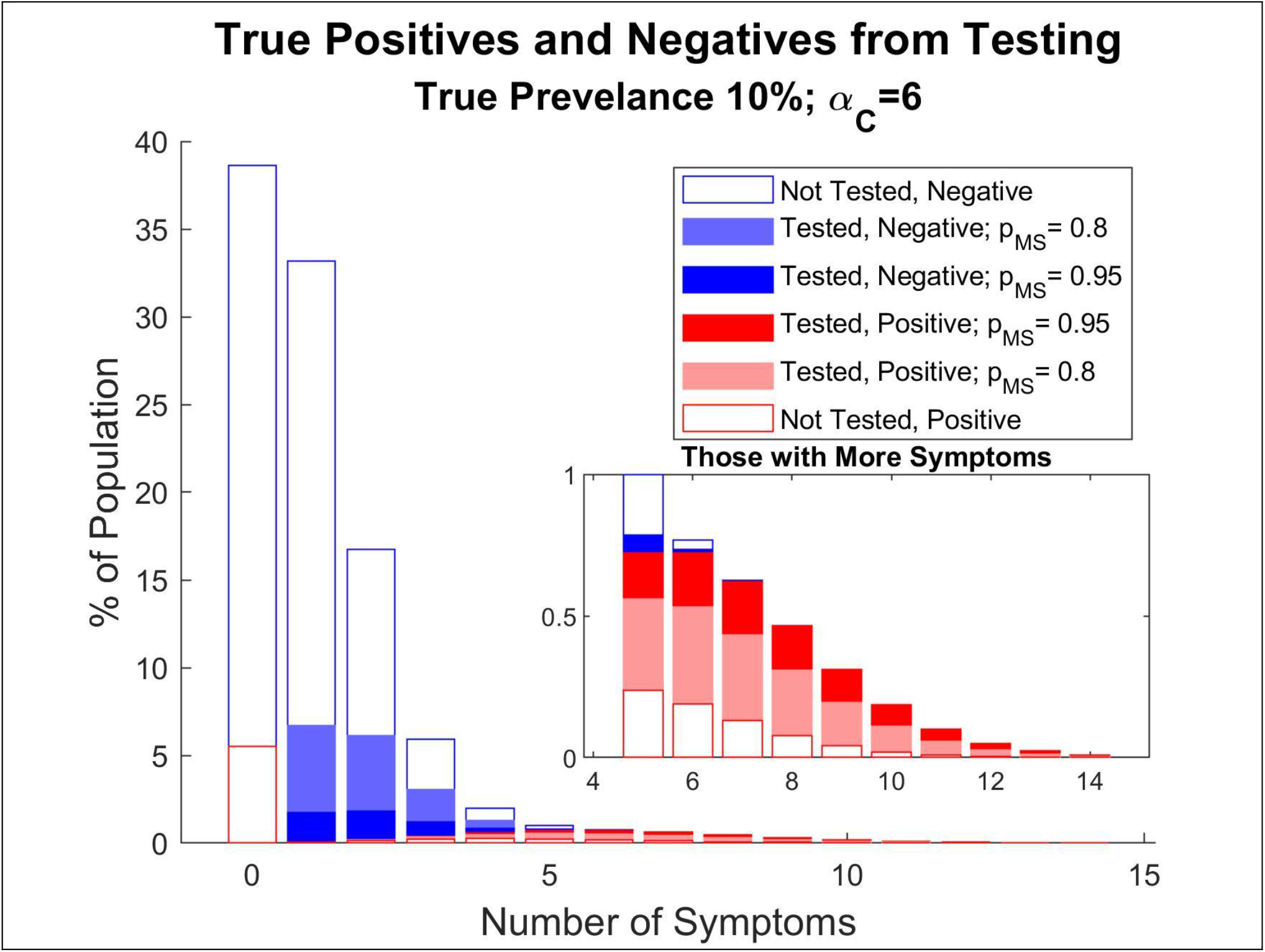
Schematic overview of zero-inflated Poisson process and test allocation. Red bars represent COVID-positive individuals and blue ones are COVID- negative. Asymptomatic fraction is assumed to be 55% for COVID patients with the symptomatic cases following a Poisson distribution with mean 6. Color coded bars signal fraction of tested individuals with different levels of probability of missing symptoms, P_MS_.

Having solved for *p\**_*MS*_ (numerically), we analytically calculate the average acuity level for those positively tested (α_CP_: *Average Acuity of Positively Tested*) and those either not tested or having received a false negative result (α_CN_). Specifically, if test sensitivity was 100%, the average acuity for those not tested would be:

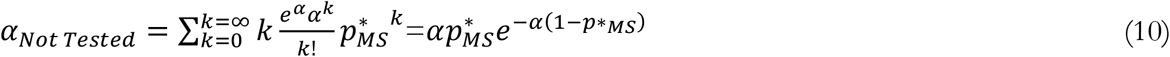

The acuity level for those tested could then be found based on the conservation of total acuity across those positively tested and those not. Starting with this basic specification we further account for the *Sensitivity of Covid Test (s*_*T*_) to calculate the values of α_CP_ and α_CN_. We parametrize sensitivity at 70%, which is the estimated sensitivity for the PCR-based tests used as the primary diagnosis method of current infections of COVID-19 (*5, 6*).

Overall, the testing rates that are determined by solving for 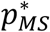, combined with sensitivity of tests, inform the fraction of COVID positive individuals transitioning from pre-detection (*I*_*P*_) to confirmed vs. unconfirmed states (*I*_*C*_ or *I*_*CH*_ vs. *I*_*U*_ or *I*_*UH*_), while the calculated α values inform the likelihood of hospitalization and fatality rates, as discussed next.

The testing sector includes the following two country level parameters that are estimated: *n*_*ST*_, *m*_*IT*_.

#### Hospitalization

The hospitalization sector of the model starts with each country’s *Nominal Hospital Capacity* (*h*_*N*_) in total hospital beds. In practice, geographic variation in hospital density and demand creates imperfect matching of available beds with cases of COVID-19 at any point in time, e.g. because some potential capacity is physically distant from current COVID hotspots. This imperfect matching means some of the nominal hospital capacity is effectively unavailable at any time, especially in larger, less densely populated countries. We therefore calculate *Effective Hospital Capacity* (*h*_*E*_) by considering geographic density of hospital beds (*Bed per Square Kilometer; d*_*H*_):

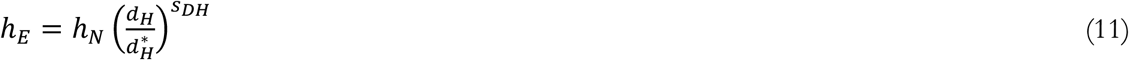

Where the 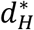 represents a large *Reference Hospital Density* of 6.06 beds per km^2^ (which is the value of *d*_*H*_ for South Korea). The parameter *s*_*DH*_ (*Impact of Population Density on Hospital Availability*) is estimated.

Effective capacity is allocated between *Potential Hospital Demand* (*H*_*CD*_) from COVID-19 cases and the regular demand for hospital beds from all other conditions (which we assume equals pre-pandemic effective hospital capacity). We assume that COVID-19 patients will have higher priority for hospitalization compared to regular demand. Specifically, we assume that fraction of regular demand allocated (*m*_*HR*_) would be the square of that for COVID demand 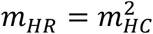, and solve the resulting hospital capacity allocation problem analytically:

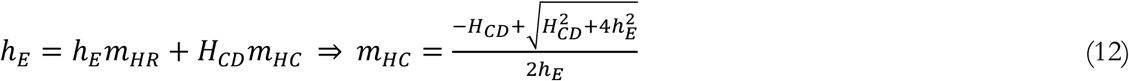

We determine the COVID demand for hospitalization based on a screening process similar to that for testing. Two types of COVID patients may seek hospitalization: those with confirmed test results and those without. The former are more likely to seek hospital treatment. We first calculate a parameter analogous to *p*_*MS*_ in the testing sector that informs the demand from confirmed COVID patients for hospitalization. This parameter, the *PMAS Confirmed for Hospital Demand (p*_*MHC*_) is determined based on acuity level of confirmed (α_CT_) and *Reference COVID Hospitalization Fraction Confirmed* (*r*_*H*_), an estimated parameter capturing the overall need for hospitalization among COVID patients:

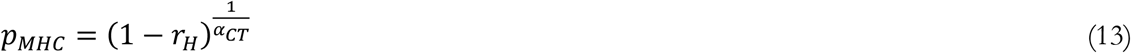

For unconfirmed COVID patients we scale the analogue of this parameter (*p*_*MHU*_) based on how much priority non-COVID patients generally receive:

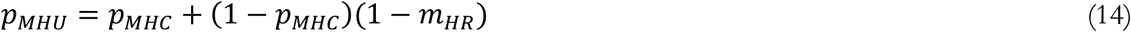

This formulation ensures that: 1) Confirmed COVID patients are more likely to be hospitalized, but also that 2) if there is ample hospital capacity (*m*_*HR*_∼1), then confirmed and unconfirmed COVID patients will receive similar priority for the same level of acuity. In short, the *p*_*M*._ values determine hospital demand by confirmed and unconfirmed COVID patients, which add up to *H*_*CD*_. The latter determines the fraction of hospital demand that is met. Analogous to the testing sector, this fraction along with demand determines the flow of individuals from the pre-detection (*I*_*P*_) state to hospitalized vs. non-hospitalized states (*I*_*CH*_ or *I*_*UH*_ vs. *I*_*C*_ or *I*_*U*_). Matching demand to allocated capacity also allows us to calculate the realized *Probability of Missing Acuity Signal at Hospitals* (*p\**_*M*_) for confirmed and unconfirmed patients. As in the testing sector, those probabilities let us approximate for the expected acuity levels for COVID patients in and out of hospital, as well as tested vs. not-tested, i.e. α_*CT*_, α_*CH*_, α_*U*_, and α_*UH*_. These average acuity levels in turn inform fatality rates for each group.

The hospital sector includes two country level estimated parameter with limited variation across countries: *s*_*DH*_ and *r*_*H*_.

### Infection Fatality Rates

For patients in each of the _*U, C, CH*_, and _*UH*_ groups we specify the *Infection Fatality Rate* (*f*), as:

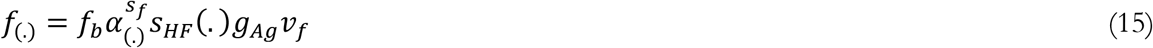

The parameter *Base Fatality Rate for Unit Acuity* (*f*_*b*_) sets the baseline for fatality rate. *Sensitivity of Fatality Rate to Acuity* (*s*_*f*_) determines how fatality changes with estimated acuity levels; more severe cases are expected to have higher fatality rates. Hospitalization reduces fatality rates, expressed as the relative *Impact of Treatment on Fatality Rate* (*s*_*HF*_); Finally, IFR reduction due to heterogenous responses (e.g. high risk groups becoming more cautious as cases accumulate), improved treatment with learning curves, and other drivers is captured in *Time variant change in fatality* (*v*_*f*_).

The *g*_*Ag*_ function incorporates the impact of age distribution on fatality rates. For age effect, we calculate a risk factor for each country. We use data from the World Bank on the age distribution of each country’s population in 10-year age strata to calculate an age-weighted average of the IFRs for COVID patients by 10-year age group reported in prior work (*7*). We normalize this age-weighted average IFR against its value for China, where the data on IFRs by age group were originally recorded. Normalizing in this way means the age effect is not sensitive to any systematic over- or under-estimation of the IFR in prior work, only to the relative risk by age group. The resulting normalized age effect ranges from 0.271 (Kenya, median age ∼20 years) to 2.368 (Japan, median age ∼48 years). Given the well-established impact of age on fatality, this factor is directly multiplied into the infection fatality equations.

Finally, we formulate the *v*_*f*_ factor as a function of cumulative cases to-date in each country using a standard learning curve formulation, bounded by a minimum multiplier that is 10% baseline, and starts to operate after cases reach 0.5% of population. The *Learning and Death Reduction Rate, l*_*IFR*_, is estimated for each country. Specifically:

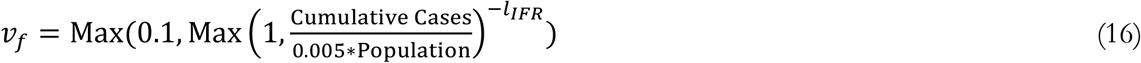

Overall, the fatality sector includes three parameters that are estimated at the country level, with limited variance across countries, those are: *f*_*b*_, *s*_*HF*_, and *s*_*f*_. A fourth country-level parameter, *l*_*IFR*_, is allowed to very more widely across different nations.

#### Note on comorbidities and fatality

We also explored including three comorbidities but found the estimates unreliable and therefore they are not included in the main specification of the model. Those comorbidities include obesity, chronic disease, and liver disease. The effects we explored for each were: 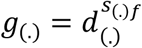, where we used the following country-level indicators from the World Health Organization (*8*), normalized by the average across all countries (*d*_(.)_):

For obesity: Prevalence of obesity among adults, BMI ≥30 (age-standardized estimate) (%)

For chronic health issues: Probability (%) of dying between age 30 and exact age 70 from any of cardiovascular disease, cancer, diabetes, or chronic respiratory disease

For liver disease: Liver cirrhosis, age-standardized death rates (15+), per 100,000 population

#### Risk Perception, Behavioral Responses, and Adherence Fatigue

In equation 3 we noted that *Contacts Relative to Normal* (*F*_*C*_) regulates infection rates. This factor ranges between a minimum (*Min Contact Fraction*; *c*_*Min*_) and 1 as a function of the impact of perceived risk on behaviors, F:

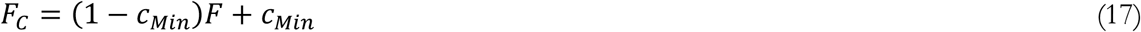

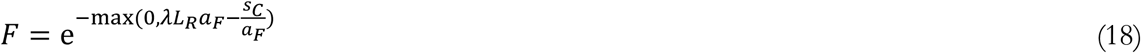

The impact of perceived risk on response uses an exponential function (eq 18) with exponent informed by *Perceived Risk of Life Loss* (*L*_*R*_), which is then moderated by a multiplier (*Dread Factor in Risk Perception*, λ) and *Impact of Adherence Fatigue* (*a*_*f*_). This moderated risk is compared to a Risk *Threshold for Response* 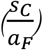, which itself responds to adherence fatigue.

*L*_*R*_ adjusts to an underlying *Indicated Risk of Life Loss* 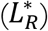 with a time constant that is asymmetric, i.e. *Time to Upgrade Risk* (*τ*_*RU*_) could be different from *Time to Downgrade Risk* (*τ*_*RD*_). The 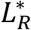 itself depends on *Perceived Hazard of Death* (*Z*_*DP*_) and a discount rate to turn daily costs to life-long ones (γ=0.03/year):

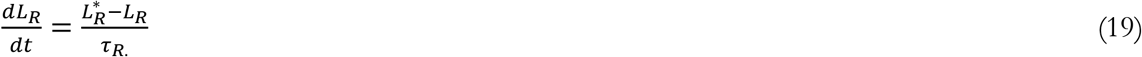

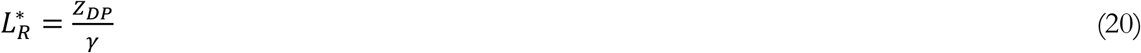

The *Perceived Hazard of Death* (*Z*_*DP*_) is an average of reported daily hazard of death (with the weight *Weight on Reported Probability of Infection, w*_*R*_) and true hazard for death which individuals may perceive through word of mouth and their social networks.

Finally, we formulate the *Impact of Adherence Fatigue* based on a 100-day exponential average of relative contacts, *Recent Relative Contacts* (*F*_*R*_) and a country-specific estimated parameter, *Strength of Adherence Fatigue* (s_a_):

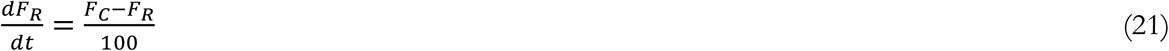

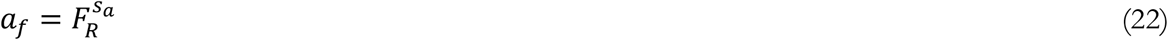

Overall, the risk perception and response sector includes the following six country-specific parameters that are estimated: *c*_*Min*_, *τ*_*RU*_, *τ*_*RD*_, λ, *w*_*R*_, *and s*_*a*_.

#### Vaccination

We include a simple vaccination sector in the model to inform policy analyses. This sector was not active in the estimation of the model and most of the analyses reported in the paper, but is operational for vaccination scenarios reported in future projections. It is formulated using the following assumptions:

- Vaccines are perfect in stopping transmission to vaccinated. Therefore they move individuals from the “Susceptible” stock to “vaccinated” where they remain for the rest of simulation.
- Individuals may opt not to vaccinate. A user of the model can specify a fraction of population not accepting the vaccine, and those individuals are assumed to be represented with the same fraction across different population stocks. The scenarios simulated in the paper assume this fraction is zero but the online simulator allows for changing that fraction.
- All individuals willing to vaccinate will get vaccinated regardless of their prior COVID infection status. However, vaccines are effective only on susceptible individuals, so those actively infected at the time of vaccination will not be affected by vaccine.
- Vaccination rate is set based on a user-specified vaccination period. The rate will ramp up linearly for a given fraction of this period, and then will remain constant for the remainder. The final rate is specified such that everybody will get the vaccine within the specified vaccination period. In the reported simulations, the ramp-up is assumed to be fast and the overall period is set to one year, starting from January 2021. In the online simulator the ramp up period is assumed to be half the overall vaccination period, and users can input the overall period.
- Vaccination could follow a priority plan in which higher-risk individuals are vaccinated first. In the model this mechanism is implemented by tracking a co-flow of acuity for all susceptible individuals. Vaccination is allowed to drain this acuity coflow with a rate that exceeds average acuity in the susceptible population by a user-specified ratio. In the reported scenarios we use a draining factor 1.5 times the average acuity in the stock of susceptibles. The average acuity in susceptible population would then be the *α*_*C*_ used in the formulations above (with initial *α*_*C*_ starting from the empirically estimated value), and will change dynamically in response to vaccination of elderly and other high-risk groups and the potentially faster draining of acuity coflow.

#### Summary of Key Equations and Parameters

Table S1 summarizes the main equations discussed in S1, providing the mapping between full variable names and the short forms. It also includes all estimated model parameters, as well as those specified based on prior research.

**Table S1.** Mapping between full variable names and their short form for the subset of variables and parameters discussed in S1. Also included are equations explained above and sources for other variables.

| Short form | Full variable name | Equation/Source |
| --- | --- | --- |
| $T_t$ | Active Test Rate | Data (See S4 for pre-processing details) |
| $m_{HC}$ | Allocated Fraction COVID Hospitalized | $\frac{-H_{CD} + \sqrt{H_{CD}^2 + 4h_E^2}}{2h_E}$ |
| $m_{HR}$ | Allocated Fration NonCOVID Hospitalized | $m_{HC}^2$ |
| $\alpha_{CP}$ | Average Acuity of Positively Tested | See full documentation |
| $f_b$ | Base Fatality Rate for Unit Acuity | Estimated |
| $n_{ST}$ | Baseline Daily Fraction Susceptible Seeking Tests | Estimated |
| $d_H$ | Bed per Square Kilometer | Data (8) |
| $m_T$ | Confirmation Impact on Contact | Estimated |
| $F_C$ | Contacts Relative to Normal | $e^{-\max(0, \lambda L_R a_F - \frac{S_C}{a_F})} (1 - c_{Min}) + c_{Min}$ |
| $\alpha_C$ | Covid Acuity | Estimated |
| $R_W$ | CRW | Use estimates from (1) |
| $g_{AG}$ | Demographic Impact on Fatality Relative to China | Use estimates based on (8, 9) |
| $\gamma$ | Discount Rate per Day | $8.2e^{-5}$ /Day |
| $\lambda$ | Dread Factor in Risk Perception | Estimated |
| $h_E$ | Effective Hospital Capacity | $h_N \left( \frac{d_H}{d_H^*} \right)^{S_{DH}}$ |
| $n_{DCH}$ | Fraction Covid Death In Hospitals Previously Tested | See full documentation |
| $n_{PM}$ | Fraction of Fatalities Screened Post Mortem | $n_{DCH}$ |
| $I_{CH}$ | Hospitalized Infectious Confirmed | See full documentation |
| $I_{UH}$ | Hospitalized Infectious Unconfirmed | See full documentation |
| $\tau_H$ | Hospitalized Resolution Time | 20 Days |
| $s_a$ | *Impact of Adherence Fatigue* | $F_R^{s_a}$ |
| $S_{DH}$ | Impact of Population Density on Hospital Availability | Estimated |
| $S_{HF}$ | Impact of Treatment on Fatality Rate | Estimated |
| $\tau_P$ | Incubation Period | 5 days |
| $L_R^*$ | Indicated Risk of Life Loss | $L_R^* = \frac{Z_{DP}}{\gamma}$ |
| $I_P$ | Infected pre Detection | See full documentation |
| $I_U$ | Infected Unconfirmed Post-Detection | See full documentation |
| $f(\cdot)$ | Infection Fatality Rate (·) | $f_b \alpha_{(\cdot)}^{sf} s_{HF}(\cdot) g_{Ag} v_f$ |
| $r_{SP}$ | Infection Rate | $C_I W \left( \frac{S}{N} \right)$ |
| $I_C$ | Infectious Confirmed Not Hospitalized | See full documentation |
| $C_I$ | Infectious Contacts | $\beta F_C (m_a (P^a + I_P^a + I_U^a) + I_P^s + I_U^s + P^s + I_{UH} + m_T (I_{CH} + I_C))$ |
| $l_{IFR}$ | *Learning and Death Reduction Rate* | Estimated |
| $c_{Min}$ | Min Contact Fraction | Estimated |
| $m_{IT}$ | Multiplier Recent Infections to Test | Estimated |
| $h_N$ | Nominal Hospital Capacity | Data |
| $\tau_T$ | Onset to Detection Delay | 5 Days |
| $T_0$ | Patient Zero Arrival Time | Estimated |
| $Z_{IP}$ | Perceived Hazard of Infection | See full documentation |
| $L_R$ | Perceived Risk of Life Loss | $\frac{dL_R}{dt} = \frac{L_R^* - L_R}{\tau_R}$ |
| $N$ | Population | Data (10) |
| $M_C$ | Positive Candidates Interested in Testing Poisson Subset | $\frac{I_P}{\tau_T} (1 - a)$ |
| $T_{PM}$ | Post Mortem Tests Total | See full documentation |
| $\tau_R$ | Post-Detection Phase Resolution Time | 10 Days |
| $H_{CD}$ | Potential Hospital Demand | See full documentation |
| $M_N$ | Potential Test Demand from Susceptible Population | $n_{ST} N_U + m_{IT} T_{PIR}$ |
| $T_{PI}$ | Positive Tests of Infected | See full documentation |
| $P$ | Pre-Symptomatic Infected | See full documentation |
| $p_{MHC}$ | PMAS Confirmed for Hospital Demand | $(1 - r_H)^{\frac{1}{\alpha_C}}$ |
| $p_{MHU}$ | PMAS Unconfirmed for Hospital Demand | $p_{MHC} + (1 - p_{MHC})(1 - m_{HR})$ |
| $p_{MS}$ | Prob Missing Symptom | From solution to equation 9 |
| $T_{PIR}$ | Recent Detected Infections | See full documentation |
| $r_H$ | Reference COVID Hospitalization Fraction Confirmed | Estimated |
| $\beta$ | Reference Force of Infection | Estimated |
| $d_H^*$ | Reference Hospital Density | Data (8) |
| $m_a$ | Multiplier Transmission Risk for Asymptomatic | Estimated |
| $F_R$ | *Recent Relative Contacts* | $\frac{dF_R}{dt} = \frac{F_C - F_R}{100}$ |
| $s_T$ | Sensitivity of Covid Test | 0.7 |
| $s_f$ | Sensitivity of Fatality Rate to Acuity | Estimated |
| $s_C$ | Sensitivity of Contact Reduction to Utility | Estimated |
| $s_W$ | Sensitivity to Weather | Estimated |
| $s_a$ | *Strength of Adherence Fatigue* | Estimated |
| S | Susceptible | See full documentation |
| $T_{Net}$ | Testing Capacity Net of Post Mortem Tests | $T_t - T_{PM}$ |
| $M_T$ | Testing Demand | $M_N + M_C$ |
| $\tau_{RD}$ | Time to Downgrade Risk | Estimated |
| $\tau_{RU}$ | Time to Upgrade Risk | Estimated |
| $v_f$ | Time Variant Change in Fatality | $\text{Max}(0.1, \text{Max}\left(1, \frac{\text{Cumulative Cases}}{0.005 * \text{Population}}\right)^{-l_{IFR}})$ |
| $p_a$ | Total Asymptomatic Fraction | Estimated |
| $U_L$ | Utility from Limited Activities | $e^{0.5sc}$ |
| $U_N$ | Utility from Normal Activities | $e^{sc\alpha_f(1-L_R)}$ |
| $w_R$ | Weight on Reported Probability of Infection | Estimated |
| W | Weather Effect on Transmission | $R_W^{sw}$ |

### S2 ESTIMATION METHOD

#### Overview of the Approach

The model we estimate is nonlinear and complex, and any estimation framework is unlikely to have clean analytical solutions or provable bounds on errors and biases. Therefore, in designing our estimation procedure we apply 3 guideposts: 1) Being conservative by incorporating uncertainties. 2) Avoid over-fitting; and 3) Enhance generalizability and robustness of estimates and projections. To these ends: we use a likelihood function that accommodates overdispersion and autocorrelation (negative binomial); we utilize a hierarchical Bayesian framework to couple parameter estimates across different countries which reduces the risk of over-fitting the data; and we use the conceptual definitions of parameters and their expected similarity across countries to inform the priors for the magnitude of that coupling across countries. Compared to more common choices in similar estimation settings (e.g. use of Gaussian likelihood functions), these choices tend to widen the credible regions for our estimates and reduce the quality of the fit between model and data. In return, we think the results may be more reliable for projection, more informative about the underlying processes, and better reflective of uncertainties in such complex estimation settings. We also conduct a validation test of our estimation framework using synthetic data in section S3.

The model is a deterministic system of ordinary differential equations with a set of known and unknown parameters. The known parameters are those specified based on the existing literature and do not play an active role in estimation. The unknown parameters can be categorized into those that vary across different countries and those that are the same across all countries (i.e. “general” parameters). The estimation method is designed to identify both the most likely value and the credible regions for the unknown parameters, given the data on reported cases and deaths (and for a subset of countries, the excess deaths). This is done through a combination of estimating the most likely parameter values in a likelihood based framework, and using Markov Chain Monte Carlo simulations to quantify the uncertainties in parameters and projections.

We first introduce the 3 different components of the likelihood function we use: the fit to time series data, the random effects component coupling country-level parameters, and the penalty for excess mortality. Then we explain the implementation details.

#### The Fit to Time Series for Cases and Deaths

Define model calculations for expected reported cases and deaths for country *i* as *μ*_*ij*_(*t*) (with index *j* specifying cases and deaths) and the observed data for those variables as *y*_*ij*_(*t*); the country-level vector of unknown parameters as ***θ***_***i***_ and the general unknown parameters as ***ϕ***. Note that ***θ***_***i***_ vector includes several parameters, each specifying an unknown model parameter, such as Impact of Treatment on Fatality, or Total Asymptomatic Fraction, for country *i*. The model can be summarized as a function *f* that produces predictions for expected cases and deaths for each country given the general and country-specific parameters:

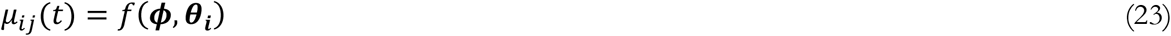

We use a negative binomial distribution to specify the likelihood of observing the y values given ***θ*** and ***ϕ***. Specifically, the logarithm of likelihood for observing the data series *y* given model predictions *μ*(***θ***, ***ϕ***) is:

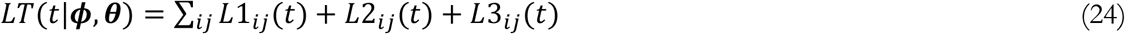

where (dropping time index for clarity):

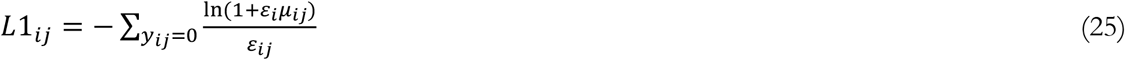

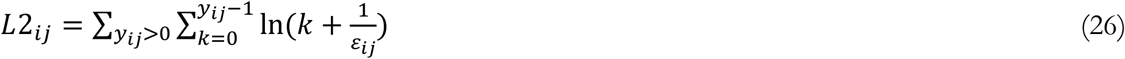

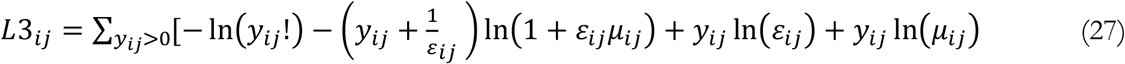

Summing the LT function over time provides the full (log) likelihood for the observed data given a parameterization of the model. The negative binomial likelihood function includes two parameters, μ and ε which determine the mean and the scaling/shape of the observed outcomes. The second parameter, ε, provides the flexibility needed fit outcomes with fat tails and auto-correlation. This parameter could itself be subject to search in the optimization process. Specifically, we assume that:

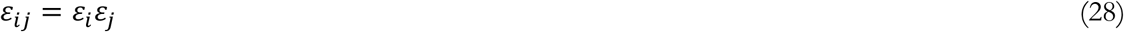

Thus we create a (set of) country specific parameter(s) (*ε*_*i*_) and two general parameters (*ε*_*j*_) which should be estimated along with the conceptual model parameters. The country level scale (*ε*_*i*_) implicitly assesses the reliability and inherent variability in country level reports, and the general ones inform the variability in case data vs. deaths. We augment the vectors ***ϕ*** and ***θ*** to include these scaling parameters as well.

#### Incorporating the coherence of parameters across countries

Up to this point we have not included any relationship among country specific parameters, ***θ***_***i***_. This independence assumption would allow parameters representing the same underlying concept to vary widely across different countries. Such treatment, by providing more flexibility, enhances the model’s fit to historical data. However, it ignores the conceptual link that exists for a given parameter across countries, potentially allowing the model to fit the data for the wrong reasons (i.e. using parameter values that do not correspond to meaningful real world concepts). The result would likely be less reliable and also not robust for future projections. We therefore define a Hierarchical Bayesian framework to account for the potential dependencies among model parameters. Specifically, we assume the same conceptual parameters (e.g. Impact of Treatment on Fatality), across different countries, are coming from an underlying normal distribution with an unknown mean (to be estimated) and a pre-specified prior for the standard deviation. This assumption is similar to the use of “Random Effect” models common in regression frameworks, though we deviate from canonical random effect models by pre-specifying the standard deviation. In fact it is possible to estimate the standard deviation across countries as well (and to obtain better fits to data by including the additional degrees of freedom), but adding those degrees of freedom ignores qualitatively relevant insights about the level of coupling across different countries for each parameter, and thus results may fit the data better but for the wrong reasons. For example, some parameters, such as Patient Zero Arrival Time, could be very different across countries, whereas parameters reflecting innate properties of the SARS-CoV-2 virus itself (e.g. *Total Asymptomatic Fraction* (*a*)) or those determining fatality (e.g. *Base Fatality Rate for Unit Acuity* (*f*_*b*_)) should be very similar across different countries. Allowing the model to determine the variance for the latter will lead to better fits: the model can find baseline fatality rates that easily match fatality variations across countries, and would expand the corresponding variance parameter accordingly. However, as a result the estimation algorithm will have too easy a job: it will not require a precise balancing between hospitalization, impact of acuity on fatality, and post-mortem testing decisions to fit fatality data. Thus, the estimates may well be less informative, or further from true underlying processes and the general characteristics of the disease which we care about. Overall, our implementation of a hierarchical Bayesian estimation framework to account for the coupling among the variables may reduce the apparent quality of fit but offer more robust results better informing the underlying mechanisms.

The implementation of this random effect introduces another element to the overall likelihood function:

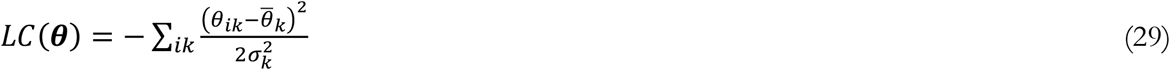

Here *θ*_*ik*_ represents the *k*^th^ parameter for country *i*, and 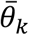 is the (estimated) average across countries for the *k*^th^ parameter. σ_k_ is the pre-specified allowable variability for the *k*^th^ parameter across different countries.

In setting these factors we chose small values for factors representing biological and natural processes, while adding more room for variation when human behaviors and perceptions were involved (See Table S2 for those settings). Specifying these standard deviation priors adds a subjective element to the estimation process. We note that subjective elements are ultimately indispensable in any modeling activity: from specifying the model boundary to the level of aggregation, use of various functional forms, and choice of likelihood functions, these choices are built on subjective assessments that experts bring to a modeling project. Absent our conceptually informed variability factors, we would need to make the assumption that country-level parameters are independent, or that our complex estimation process would correctly identify the true dependencies among those parameters. We think both those alternatives are inferior in the chosen method. So here we focus on transparently documenting and explaining those assumptions, and Supplement S3 provides a validation experiment. Table S2 summarizes the estimated model parameters, their estimated values (mean across countries and mean of Inter-Quartile Range) and the assumed variability factor (*σ*_*k*_) for each.

**Table S2.** Estimated model parameters, their estimated values (mean and standard deviation (std) across countries and the mean of Inter-Quartile Range (MIQR). Last column reports the variability allowances used to specify the coupling among country-level estimates. See equation 29 and related discussions above.

| Parameter Name | | Mean | StDev | MIQR | Variability Factor, $\sigma_k$ |
| --- | --- | --- | --- | --- | --- |
| $f_b$ | Base Fatality Rate for Unit Acuity** | 5.74E-04 | 7.30E-06 | 1.27E-05 | 1.00E-05 |
| $n_{ST}$ | Baseline Daily Fraction Susceptible Seeking Tests | 8.36E-04 | 5.70E-04 | 1.72E-04 | 0.0005 |
| $m_T$ | Confirmation Impact on Contact | 1.73E-01 | 1.24E-01 | 9.90E-02 | 0.1 |
| $\alpha_C$ | Covid Acuity** | 5.92E+00 | 1.21E-03 | 1.29E-02 | 0.01 |
| $\lambda$ | Dread Factor in Risk Perception* | 6.14E+03 | 1.28E+04 | 3.74E+03 | $10^{0.8}$ |
| SDH | Impact of Population Density on Hospital Availability | 1.70E-01 | 1.62E-01 | 9.23E-02 | 0.1 |
| SHF | Impact of Treatment on Fatality Rate | 4.58E-01 | 2.17E-01 | 6.65E-02 | 0.1 |
| lIFR | Learning and Death Reduction Rate | 7.07E-01 | 7.42E-01 | 1.67E-01 | 0.5 |
| c<sub>Min</sub> | Min Contact Fraction | 7.91E-02 | 4.99E-02 | 2.28E-02 | 0.03 |
| m<sub>IT</sub> | Multiplier Recent Infections to Test | 4.49E+01 | 2.38E+01 | 9.47E+00 | 30 |
| m<sub>a</sub> | Multiplier Transmission Risk for Asymptomatic** | 2.93E-01 | 2.55E-03 | 1.29E-02 | 0.02 |
| T<sub>0</sub> | Patient Zero Arrival Time | 9.90E+01 | 2.98E+01 | 4.59E+00 | Uniform |
| β | Reference Force of Infection | 4.07E-01 | 2.19E-01 | 3.34E-02 | 0.2 |
| r<sub>H</sub> | Reference COVID Hospitalization Fraction Confirmed | 6.32E-01 | 1.25E-01 | 8.65E-02 | 0.1 |
| s<sub>f</sub> | Sensitivity of Fatality Rate to Acuity** | 2.14E+00 | 2.35E-03 | 6.42E-03 | 0.005 |
| s<sub>C</sub> | Sensitivity of Contact Reduction to Utility | 3.17E+00 | 5.27E+00 | 1.35E+00 | 6 |
| s<sub>a</sub> | Strength of Adherence Fatigue | 1.24E+00 | 1.07E+00 | 1.91E-01 | 0.5 |
| τ<sub>RD</sub> | Time to Downgrade Risk* | 2.45E+02 | 1.88E+02 | 6.43E+01 | 10<sup>0.3</sup> |
| τ<sub>RU</sub> | Time to Upgrade Risk* | 3.83E+01 | 5.34E+01 | 1.33E+01 | 10<sup>0.2</sup> |
| a | Total Asymptomatic Fraction ** | 4.99E-01 | 9.15E-03 | 1.28E-02 | 0.03 |
| w<sub>R</sub> | Weight on Reported Probability of Infection | 4.52E-01 | 2.62E-01 | 2.05E-01 | 0.2 |
| s<sub>W</sub> | Sensitivity to Weather | 2.64E+00 | (not applicable for global parameter) |  |  |
| *Given the wide range and potential long tail for these parameters the Log<sub>10</sub> transformation is used in specifying the dispersion penalty (equation 27) and variability factors are reported as 10<sup>σ</sup>, where σ is used in equation 27. |  |  |  |  |  |
| ** These parameters are expected to be less variable across countries and thus are assigned small variability allowances compared to their mean. |  |  |  |  |  |

#### Excess mortality penalty

Finally, we include a likelihood-based penalty term to allow model predictions be informed by excess mortality data collected by various news agencies and researchers for a subset of countries in our sample. These data provide snapshots of excess mortality (compared to a historical baseline) for a window of time in each country. Subtracting from total excess mortality the COVID-19 deaths officially recorded in that window offers a data point for excess mortality not accounted for in official data (e_i_). We can calculate in the model the counter-part for this construct: the simulated mortality that is not included in the simulated reported COVID-19 deaths (*ē*_*i*_). There is uncertainty in these excess mortality data: the historical baselines used by various sources do not adjust for demographic change, excess mortality may be due to factors other than COVID-19, and some of it may be due to changes in healthcare availability and utilization motivated by COVID-19 but not directly attributable to the disease (for example when surgeries are delayed, hospitalization is avoided, or heart conditions are ignored). Excess mortality may also be reduced due to reduced traffic accidents (in light of physical distancing policies) and pollution related deaths. Given these uncertainties, we use the following penalty function to keep the simulated unaccounted excess mortality close to data:

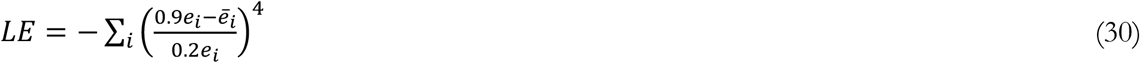

This penalty could be seen as a likelihood coming from the probability distribution 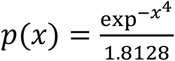 defined for all values of *x*. It assumes that in the most likely case for excess mortality, 90% of unaccounted mortality should be attributed to COVID-19 deaths, but that there is significant uncertainty around this, so some 20% variation across this figure is quite plausible (70%-110% of data). However, numbers outside of this range start to impose increasingly large penalties, so that very large deviation becomes unlikely.

Combining these three components, we obtain the full likelihood function used in the analysis:

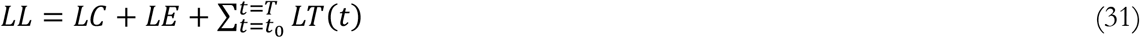

For each country we include the LT component from the first day they have reached 0.1% of their cumulative cases to-date, or a minimum of 50 cumulative cases. This excludes very early rates that are both unreliable and which, given very small estimated model predictions for infection, could lead to unreasonably large likelihood contributions.

#### Numerical Methods

The model includes a large number of parameters to be estimated: a general parameter for the impact of weather, 2 general parameters for *ε*_*j*_, and 22 parameters for each country that are coupled together based on the random effects framework described above. Out of those 1 parameter (per country) is for *ε*_*i*_ and the other 21 are informing various features of disease transmission, testing, hospitalization, and risk perception and response. With a sample of 92 countries, this would lead to 2027 parameters to be estimated. A direct optimization approach to this problem suffers from potential risk of getting stuck in local optima, and direct use of MCMC methods to find the promising regions of parameter space suffers from the curse of dimensionality. We therefore designed the following 4-step procedure to find more reliable solutions to both problems and the synthetic data exercise in S3 provides some evidence on the effectiveness of the method.

1. We estimate the model with the full parameter vector for a smaller number of countries with larger outbreaks (3-5 countries). We use the Powell direction search method implemented in Vensim™ simulation software for this step. The method is a local search approach though it has features that allows it to escape local optima in some cases. We restart the optimization from various random points in the feasible parameter space and track the convergence of those restarts to unique local peaks. We stop this process when we are repeatedly landing on the same local peaks in the parameter space. This procedure showed that local peaks do exist, but they are not many; for example, within 100 restarts we may find 2-4 distinct peaks, with one being distinctly better than others. This quasi-global peak provides a coherent set of starting points for ***ϕ*** and 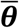 for next steps.
2. We go through iterations of the following two steps: A) Conduct country-specific optimizations with 50 restarts to find the vector of **θ**_*i*_ given the ***ϕ*** and 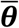 from first optimization or from the step B. B) Conduct a global optimization, including all countries but fixing ***θ***_*i*_ and optimizing on ***ϕ*** (and 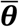; though that is simply the mean across country level parameters from previous round). We stop when iterations offer little improvement from one round to the next (less than 0.05% improvement in log-likelihood).
3. We conduct a full optimization allowing all parameters (***θ***_*i*_, ***ϕ*** and 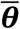) to change, starting from the point found in the last iteration of step 2. This step finds the exact peak on the likelihood landscape which is the best-fitting parameter set for the model.
4. For the MCMC, theoretically one should conduct the sampling from all model parameters in the full model. However, our experiments showed that the large dimensionality of the parameter space requires an infeasible number of samples to achieve adequate mixing and ensure reliable credible regions for parameters and projections. To overcome this challenge we note that the parameters of different countries are connected to each other only through ***ϕ*** and 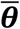, and these general parameters are rather insensitive to dynamics in each country. The insensitivity is due to the fact that a single country only contributes about 1% to the general parameters’ values, and within a typical MCMC the country-level parameters often can’t change more than 10% before the resulting samples become highly unlikely. Therefore, one can conduct an approximate country-level MCMC by fixing the general parameters at those from step 3, and only sampling from the ***θ***_***i***_ for each country. The MCMC algorithm used is one designed for exploring high dimensional parameter spaces using differential evolution and self-adaptive randomized subspace sampling (*11*). Using this method we obtain good mixing and stable outcomes (Robin-Brooks-Gelman PSFR convergence statistic remaining under 1.1) after about 600,000 samples (the burn-in period). We continue the MCMC for each country for another 400,000 samples and then randomly take a subsample of those points after the burn-in period for the next step.
5. The resulting subsamples for different countries from step 4 are assembled together to create a final sample of parameters for the full model to conduct projections and sensitivity analysis at the global scale. Uncertainties in the handful of global parameters is not identified in this procedure, but can be quantified by assessing the sensitivity of the global likelihood surface to changes in those parameters.

The process above is automated using a Python script that controls the simulation software (Vensim). We conduct the analysis using a parallel computing feature of Vensim on a Windows server with 48 cores. After compiling the simulation model into C++ code (which speeds up calculations significantly), and using a simulation time step of 0.25 days, it takes about 60 hours to complete the estimation for 91 countries, and almost two weeks to complete the full suite of sensitivity analyses reported in the paper. Full analysis code is available online at https://github.com/tseyanglim/CovidGlobal.

### S3 VALIDATION OF ESTIMATION FRAMEWORK

The complexity of model and the large number of parameters involved complicates the assessment of estimation method based on theoretical considerations alone. We therefore use a synthetic data experiment to build confidence in the estimation framework. Specifically, we first simulate the model using known parameters without using historical deaths and cases to provide a ‘ground-truthed’ set of synthetic data. We then apply the exact estimation framework used on the actual data to infer the parameters of the model from this synthetic dataset. Finally, we assess how well the estimated parameters correspond to the “true” values and how inclusive the estimated credible intervals are of the true parameters. The ability of the estimation framework to find the true parameter values, and consistent credible intervals, would increase our confidence that parameters estimated using actual data are also not particularly biased and that the credible intervals are informative. While repeating this procedure for multiple sets of synthetic data, with various parameterizations, is desirable, the computational costs in our setting make such an approach infeasible. Nevertheless, the large number of parameters estimated in a single full calibration exercise provides ample opportunities to test the precision of the method in the range of parameter values relevant in the actual data. The three steps of the process are discussed below.

### Generation of synthetic data

We used the model specified above, with the parameters estimated in the baseline analysis from actual data, to generate the synthetic data. Given the deterministic nature of the model, it would be easy for the estimation process to identify the model parameters should we use the exact outcome of the baseline simulation. To test the model in a more realistic scenario, therefore, we inject two different random noise time series into the model, effectively turning the data generation simulation model into a stochastic one with underlying noise processes not accurately captured in the estimation model (because of the autocorrelation in the driving noise). Specifically, we make the following two modifications to the model equations:

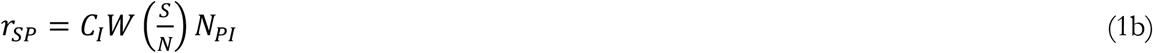

Where

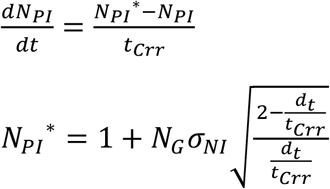

And

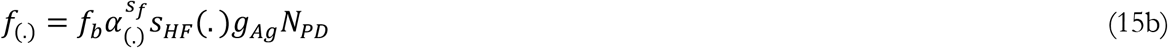

Where *N*_*P*._ are the noise terms changing infection and IFR rates. *N*_*PD*_ is formulated similar to *N*_*PI*_, with parameters *t*_*Crr*_ and *σ*_*ND*_. *N*_*G*_ is a standard Gaussian random number generator producing a new independent draw every time step of the simulation (*d*_*t*_) for each of the two noise streams separately and independently.

These equations specify two first order autocorrelated Normally distributed noise streams. The autocorrelation time constant, *t*_*Crr*_, is set to 10 days for both streams of noise. The *σ*_*NI*_ and *σ*_*ND*_ parameters are set to 0.1 (i.e. leading to standard deviation of noise around infections and deaths being 10% of the model generated baselines). As in the real world, the substantial correlation time leads to significant swings in the infection and death rates beyond those explained by model mechanisms.

We also add a “measurement” noise to both daily infections and deaths in synthetic data by drawing Negative Binomial random samples from the estimated distributions for each country at any given time and using those (rather than expected values) as the data in this estimation exercise.

To best replicate the features of actual data, the model uses actual country level data for test rates (which are exogenous inputs driving simulations) and various country level statistics such as population, population density, and age structure.

We record the data generated from this simulation for confirmed cases and deaths, corresponding to the data we have available to estimate the actual model. For each country we only record the data for the days in which we have a corresponding actual data point. We also record excess mortality counts for the subset of countries and periods for which we have such data. These three data items (two time series for confirmed infections and deaths and point estimates for excess mortalities in a subset of countries) are the inputs into the estimation process.

### Estimation using synthetic data

The synthetic data generated in the previous step is available on the project’s GitHub repository. This data is then used, following the estimation process discussed in S2, to find the model parameters. This step requires no other assumptions and follows the exact process used in the main analysis. Note that we start the estimation with uninformed (uniform with large ranges) priors on all parameters.

### Results and comparisons

Figure S5 reports the estimated parameters, their 95% credible intervals, and the true parameters across all 1932 (92 countries x 21 parameters each) country-level parameters of the model that impact outcomes. Overall, the estimation process successfully identifies the vast majority of parameters. For example the median distance between estimated and true values, as a percentage of the length of estimated 95% credible interval, is 21%. Moreover, the credible intervals envelope the true values rather consistently. Specifically, the 50% CI includes the true value in 32% of cases and this measure increases to 49%, 60%, 67%, and 75% for 80%, 90%, 95%, and 98% CIs respectively. The theoretical vs. actual intervals are showed in Figure S4.

**Figure S4.**
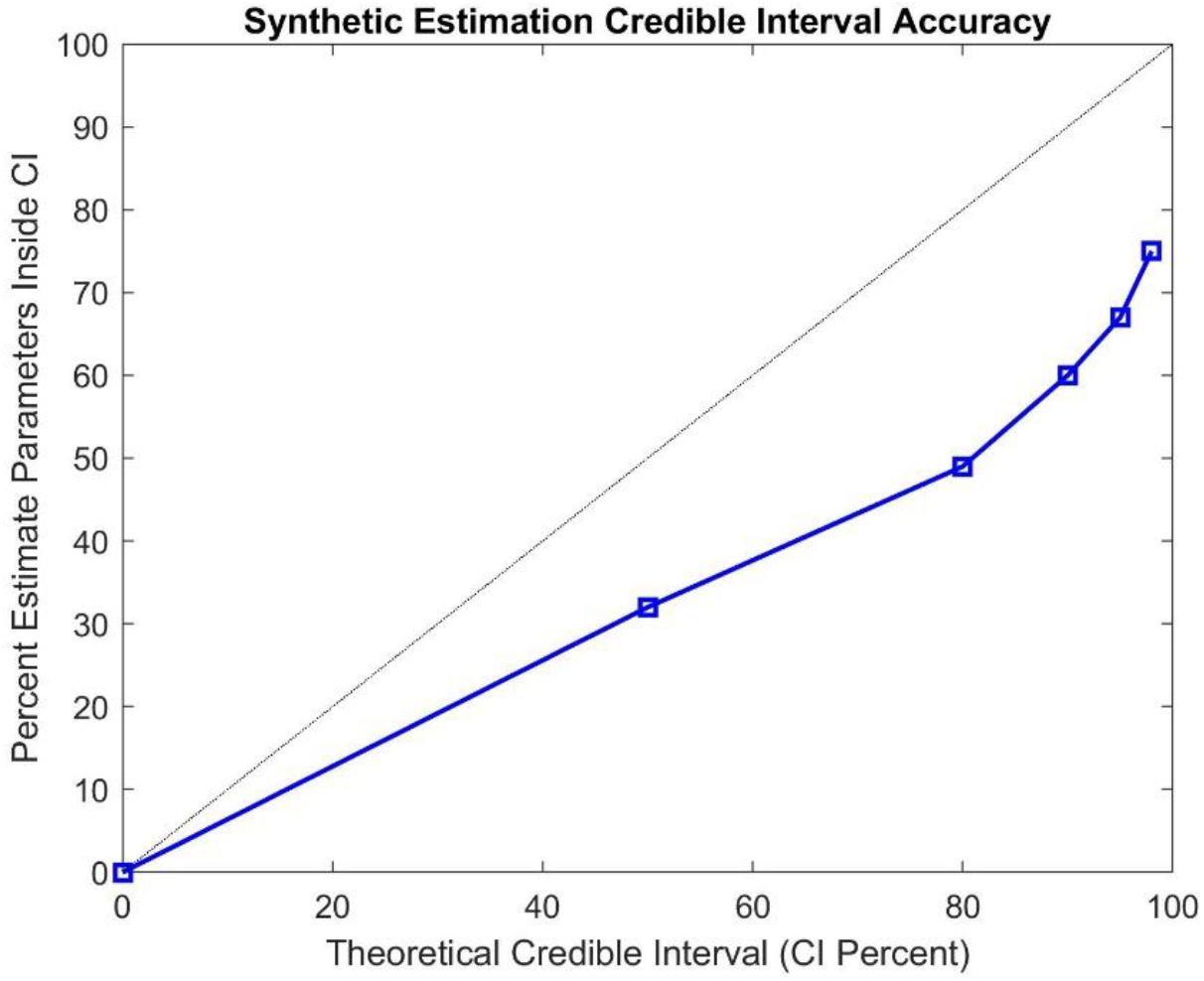
Theoretical vs. actual fraction of parameters enveloped by different Credible Interval percentiles.

While not identical to the expected theoretical values, these coverage levels are close, especially in the context of very large parameter spaces and complex estimation exercises where finding reliable CIs is often harder than estimating the parameters. Figure S5 also shows that some parameters are more likely to have imprecise confidence intervals than others. In fact, much of the imprecision in confidence intervals are due to two parameters, *Base Fatality Rate for Unit Acuity* and *Covid Acuity Relative to Flu* end up outside 95% confidence interval for all countries, despite estimated value being numerically very close to original value. We can’t rule out the existence of a local optima in the new estimated value driving the results. Moreover, for some parameters (e.g. f_b_, and *β*) the baseline estimated values could fall outside the 95% confidence intervals and are closer to the true values. These instances could point to asymmetric likelihood surfaces or the possibility that the MCMC chains may require larger samples for getting at true confidence intervals. Overall these results add to our confidence that the estimated credible intervals are in the right range, though some may be somewhat tighter than they should be.

**Figure S5.**
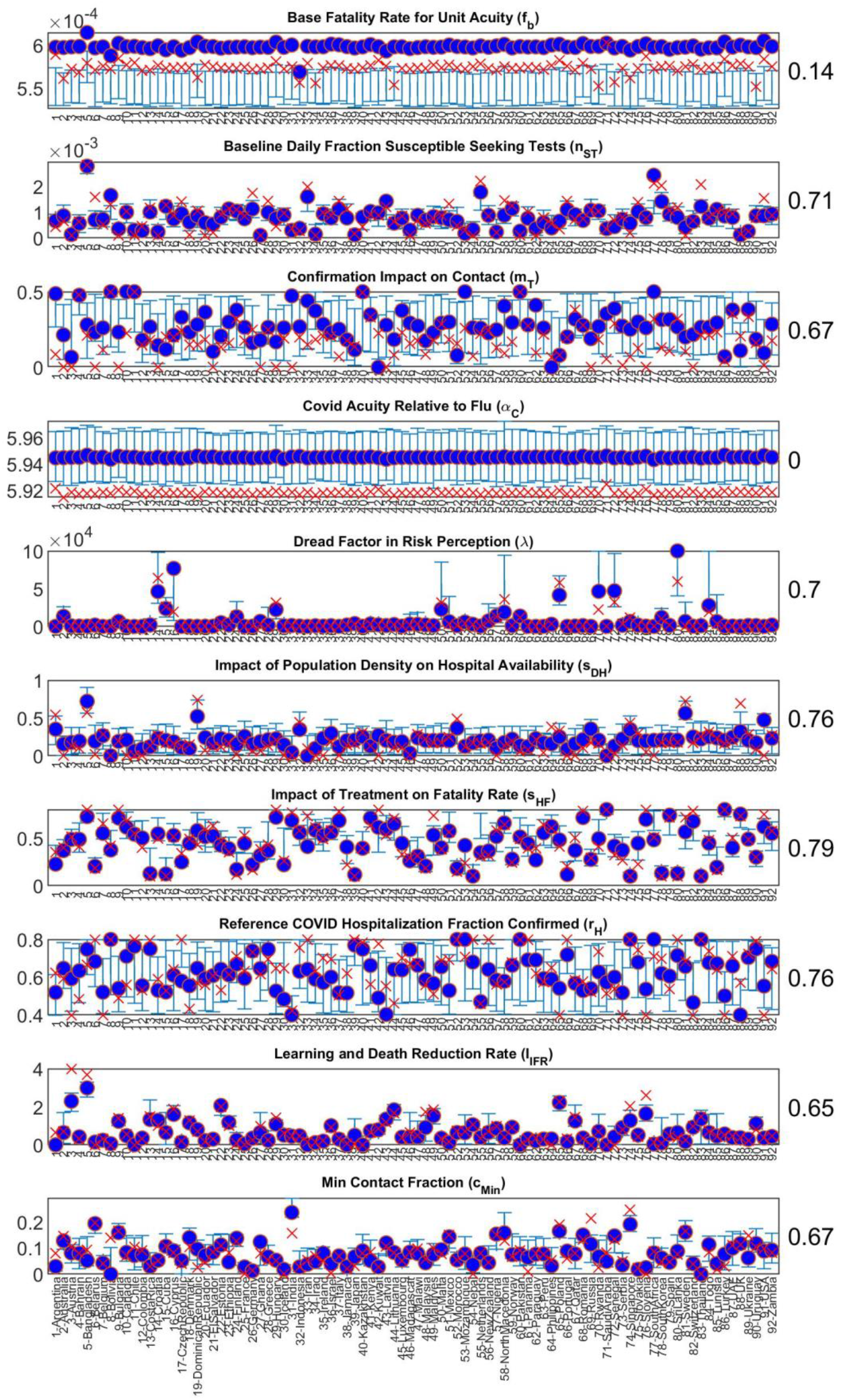
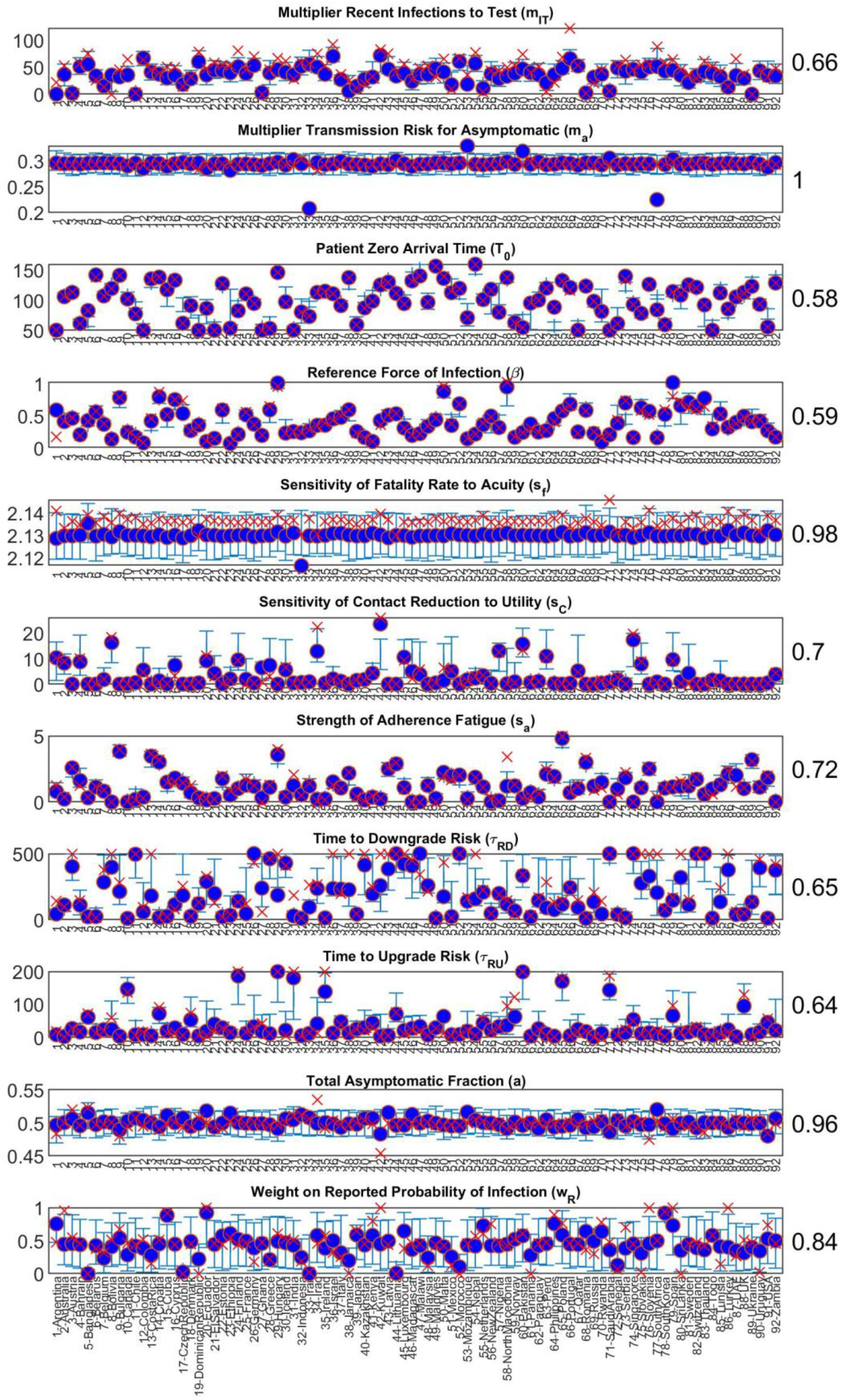
Country-level parameter estimates and 95% credible intervals from synthetic estimation exercise (blue circles and bars) compared with true values (red cross signs) across all parameters. Numbers on the right represent the fraction of true values enveloped by the 95% interval.

## S4 DATA PRE-PROCESSING

Getting contemporaneous, comprehensive, national-level data on Covid-19 is a challenge. The most widely-cited data aggregators, such as the Johns Hopkins Center for Systems Science and Engineering’s COVID-19 database (*12*), OurWorldInData portal (*13*), and the US-focused COVID Tracking Project (*14*), get their data from the same few official sources, such as the US Centers for Disease Control and Prevention (CDC), the European CDC (ECDC), and the World Health Organization (WHO). These official agencies in turn get their data from national and subnational public health authorities, which ultimately rely on reports from hospitals, clinics, and private and public health labs.

As a result, idiosyncrasies in the ground-level data collection processes permeate virtually all sources of aggregate data. Most notably, data collection involves time lags, which can differ from source to source. Daily death counts could reflect the date of actual death or the date a death is registered or reported; different UK government sources, for instance, use each of these metrics.^2^ Daily infection or case counts could include the total new cases reported *on* a given date, or the total cases confirmed *from* that date; the latter would result in some ‘backfill’ whereby case counts for previous days can continue to increase for some time as delayed confirmations come in. Daily counts of tests conducted could report samples collected, samples processed, results reported, or a mix of these; the US CDC, for instance, reports a mix of testing by date of sample collection and date of sample delivery to the CDC.^3^ Aside from differences in unit of measure (people vs. tests vs. samples), there may be different time lags involved as well. In addition to these idiosyncrasies, testing data in particular is also patchy for many countries, even as testing has become more widespread. The WHO does not report country-by-country testing, nor does the JHU Covid map outside the US. Furthermore, there are sometimes irregular delays in the reporting of test results, which can create occasional unexpected spikes in reported numbers of tests, infections, or both.^4^

Depending on the specifics of how daily infection and test counts are reported, there can in some cases be a disjunction between the two. Because confirmed case counts largely depend on positive test results, test and infection counts should be correlated – *ceteris paribus*, a day with a lot of samples collected for testing should see more confirmed cases attributed to it, while a day with no sample collection should see no cases. But since cases may not be reported by the date of the test, and tests may not be reported by the date of sample collection, officially reported numbers can get out of sync in either direction.

This problem is most salient when there are clear weekly cycles in daily rates. In most of the world, particularly western countries, daily test rates are far lower on weekends than during the week. As a result, infection numbers show a clear weekly cyclical component as well. But the weekly cycles in testing and infection numbers for a given country do not always line up. Our model explicitly accounts for the effect of testing on reported infections, but we do not explicitly model the country-level idiosyncrasies of reporting and how they vary between test data and infections. Instead we account for any such lags in pre-processing of the data to align testing and case data.

The weekly cycle occurs in many countries’ death rate data as well, where it presents a different problem. A weekly cycle in testing is a behaviourally realistic part of the data-generation process, as many labs, clinics, or other testing sites for instance may be closed on weekends. As testing provides the window on the state of confirmed infections, a comparable cycle in confirmed cases is to be expected as well. By linking case confirmations to testing, our model explicitly accounts for this limited visibility on the true state of the epidemic. However, a weekly cycle in death rates almost certainly reflects different limitations of the data-generation process, typically to do with hospital staffing,^5^ which we do not explicitly model. As such we need to address any weekly cycle in death rates through data pre-processing as well.

To deal with these challenges, we developed a multi-step algorithm to pre-process our data before feeding it into the model for calibration. The algorithm is described below. It was implemented in Python, largely using the Pandas and NumPy packages, and the code is available in full at: https://github.com/tseyanglim/CovidGlobal.

The algorithm proceeds country-by-country, following these steps on each country.

1. Examine daily cumulative test data; if data are insufficient (6 or fewer data points), drop country from the dataset.
2. Interpolate any missing daily cumulative test data points using a piecewise cubic Hermite interpolating polynomial (PCHIP) spline. If the first reported infection is before the first reported cumulative test, also extrapolate cumulative tests back to the date of first reported infection.
  a. Extrapolation to the date of first reported infection is necessary since both in the model and, to a large extent, in reality, reported infections require testing for confirmation.
  b. PCHIP spline interpolation yields a continuous monotonic function with a continuous first derivative, thus avoiding generating any anomalous rapid change in daily test rate.
  c. We used the implementation of PCHIP interpolation from the widely used SciPy package for Python.^6^
3. Calculate daily test rate as daily cumulative tests less the preceding day’s cumulative test total:

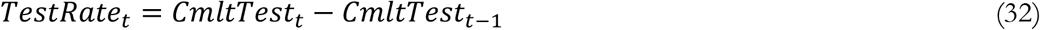
4. Examine the original daily cumulative test data to estimate how much of the calculated daily test rate is based on interpolated vs. original data.
  a. Daily test rates calculated based on mostly original data should be expected to include any weekly cycles or occasional irregularities that would also be reflected in daily infection counts. Conversely, daily test rates calculated from cumulative test counts that are largely interpolated would not be expected to fully reproduce any such cycles or irregularities, since the interpolation produces a relatively smooth function.
  b. As a rule of thumb, we examine the cumulative test data for the second half of the time from the first test to the latest test. If fewer than half the days in that window have original cumulative test data, we consider the test data to be ‘sparse’, requiring further processing.
5. If the test data are not sparse, account for any potential lag or other reporting delay differences between daily test rate and daily infection rate using a time-shift algorithm to estimate any such lags or delays from the data and shift the test rate time series accordingly. The time-shift algorithm ensures that any weekly cycles present in the daily infection rate data are reflected in the daily test rate data and aligned as best as possible on date, thereby accounting for the fact that model-generated reported infections depends on testing but with no time lag between test and result.
  a. First, identify the weekly component of the time series of daily infection rate and daily test rate using a seasonal-trend decomposition based on LOESS (STL) procedure.^7^
    i. STL deconstructs time series data into several components, including a trend and a seasonal component over a specified period (weekly, in this case) as well as a residual. STL is an additive decomposition, and has the advantage of allowing the seasonal component to change over time (rather than being a fixed pattern repeated exactly across the whole time series).
    ii. We used the STL implementation from the Statsmodels package for Python.^8^
  b. Shift the time series over a one-week range (from -2 to +4 days of lag between test and infection reporting), calculating the cross-correlation between the weekly seasonal component of the daily infection rate data and the daily test rate data for each time shift.
  c. Identify the time shift within this range that maximizes the cross-correlation between the infection rate and test rate data, and shift the test rate data accordingly.
6. If the test data are sparse, the spline interpolation will generally cut out some of any weekly cyclicality that may be present. Visual inspection of daily test rates for countries with sparse test data also shows large, irregular spikes in reported tests are not uncommon, without necessarily having concomitant irregular spikes in reported daily infection rates. As such, rather than attempting to eliminate differences in reporting lags through the time-shift algorithm described above, we instead apply a data-smoothing algorithm to both daily test rate and daily infection rate, in order to reduce any cyclicality and irregular spikes. This smoothing allows the calibration of the main model to focus on matching the underlying trends in the data.
7. In all cases, whether daily cumulative test data are sparse or not and whether infection and test rate data are smoothed or not, since weekly cycles in death data are reflective of reporting lags not captured in the model, daily death rate data is smoothed using the same algorithm.
8. The smoothing algorithm used is designed first to conserve the total number of reported cases (tests, infections, or deaths), and second to preserve some degree of variation in the time series, as some noise may be informative and retaining some is important to the calibration of the model.
  a. Starting from when the time series of daily rate (test or infection) exceeds a specified minimum value (5/day), calculate the rolling mean of the daily rate, using a centred moving window of 11 days.
  b. Calculate the residual between each day’s data point and the rolling mean for that day, and divide by the square root of the rolling mean, to get an adjusted deviation value:

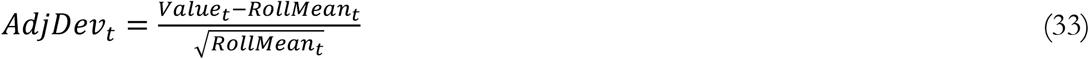
    i. Dividing by the square root of the rolling mean reflects a heuristic assumption that each daily rate (of infections, deaths, or tests) behaves as a Poisson process (StDev of Pois(λ) = λ^0.5^).
    ii. The functional result of this adjustment is that both *absolute* and *relative* magnitudes of deviations from the rolling mean are given some weight – large relative deviations when absolute values are small (and data are noisier) are not ignored, but neither do they outweigh larger absolute (but smaller relative) deviations that occur when the mean is large, which is important since most of the time series data are growing significantly over the time horizon of the model.
  c. Calculate thresholds for identifying dips and peaks in the data based on the median of the adjusted deviations, ± one median absolute deviation (MAD) of the adjusted deviations:

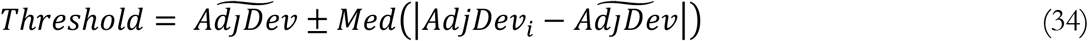
    i. Using the median absolute deviation to determine thresholds for peaks and dips is robust to outliers in the deviations, which do arise occasionally in the data.
    ii. A threshold width of one MAD is relatively narrow for outlier detection, but by inspection of the data, is about right for identifying most of the peaks and dips caused by weekly cycles in test, infection, and death rates, as well as larger outliers.
  d. Once thresholds are calculated, iterate through the data points in the time series first forward in time from oldest to newest, filling in any ‘dips’ (data points with adjusted deviations below the lower threshold), then backward in time from newest to oldest, smoothing out any ‘peaks’ (data points with adjusted deviations above the upper threshold) that remain. Repeat the process until all data points’ adjusted deviations are within the originally calculated thresholds for the time series.
    i. We infer that the underlying processes generating dips and peaks are somewhat different. Dips are generally the result of weekly cycles in the data, e.g. lower rates of testing or longer lags in death reporting that occur on weekends. Peaks arise to some extent due to the same weekly processes, e.g. some deaths that occur on weekends only being recorded at the start of the next week. However, some peaks, especially larger ones, may result from irregular random delays in reporting, such as large batches of tests being held up due to logistical issues and then getting processed all at once. As such the smoothing procedure for dips vs. peaks is slightly different.
  e. The dip-filling step fills a fraction of each dip (specified as a *smoothing factor*) by redistributing data counts based on a multinomial draw from the subsequent few days following each dip.
    i. First, calculate the amount to fill based on the deviation and the smoothing factor specified, in this case 0.67:

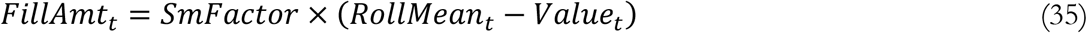
    ii. Calculate the amount redistributed from each of the following few (7) days *X*_*t*+1_, *X*_*t*+2_, … *X*_*t*+*k*_, *k* = 7, based on a multinomial distribution as follows:

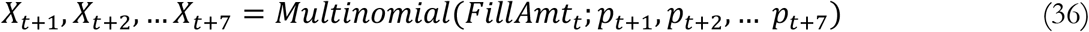 Where *p*_*t*+1_, *p*_*t*+2_, … *p*_*t*+7_ are calculated as:

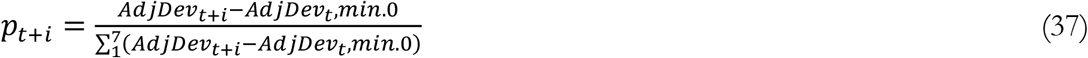
    iii. This formulation allows some redistribution from any of the subsequent few days whose adjusted deviations exceed the focal day’s adjusted deviation, but with more redistribution from days with higher adjusted deviations.
  f. The peak-smoothing step similarly redistributes a fraction of each peak, specified by the smoothing factor, to the *preceding* several days based on another multinomial draw.
    i. First, calculate the amount to redistribute similarly to the dip-filling step:

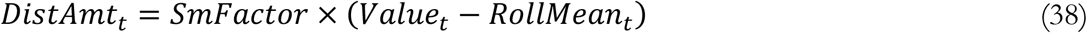
    ii. Calculate the amount redistributed *to* each of the preceding several (14) days *Y*_*t*−1_, *Y*_*t*−2_, … *Y*_*t*−*k*_, *k* = 14, based on a multinomial distribution as follows:

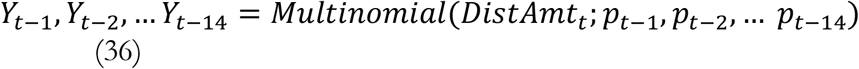 Where *p*_*t*−1_, *p*_*t*−2_, … *p*_*t*−14_ are calculated as:

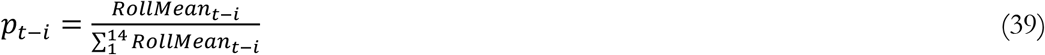
    iii. This formulation redistributes peaks to preceding days based on the calculated rolling mean counts of those days, on the assumption that the irregular delays that generate random spikes in counts are essentially random and equally likely to affect any given unit of data over a several-day span. As such, the probability that a unit showing up in a spike due to such delays comes from a given preceding day is simply proportional to the expected count for that day, as approximated by the rolling mean.
  g. By filling dips first before smoothing peaks, the combined algorithm largely addresses any peaks that are due primarily to weekly cycles during the dip-filling stage, such that remaining peaks that get smoothed tend to be the larger, irregular ones.

## S5 EXTENDED RESULTS

### Quality of fit measures

Table S3 reports two quality of fit metrics for different countries and different time series. The first four columns report Mean Absolute Error Normalized by Mean (MAEN) and the last four report the R-Squared measures. Errors for cumulative infection and deaths are followed by those for the new cases and deaths (flow variables).

**Table S3.** Measures of fit between data and simulations for different countries. Mean Absolute Error Normalized by Mean (MAEN) and R-Squared are reported for cumulative and new cases and deaths.

|  | MAEN |  |  |  | R-Squared |  |  |  |
| --- | --- | --- | --- | --- | --- | --- | --- | --- |
|  | Cumulative |  | Flow |  | Cumulative |  | Flow |  |
| Country | Infection | Death | Infection | Death | Infection | Death | Infection | Death |
| Argentina | 0.0934 | 0.0525 | 0.239 | 0.198 | 0.999 | 0.999 | 0.899 | 0.84 |
| Australia | 0.0277 | 0.288 | 0.483 | 0.65 | 0.998 | 0.97 | 0.691 | 0.72 |
| Austria | 0.239 | 0.335 | 0.576 | 0.59 | 0.991 | 0.962 | 0.852 | 0.943 |
| Bahrain | 0.11 | 0.388 | 0.421 | 0.947 | 0.988 | 0.944 | 0.414 | 0.0946 |
| Bangladesh | 0.0208 | 0.028 | 0.161 | 0.14 | 0.999 | 0.999 | 0.799 | 0.808 |
| Belarus | 0.109 | 0.123 | 0.182 | 0.494 | 0.996 | 0.972 | 0.918 | 0.466 |
| Belgium | 0.328 | 0.363 | 0.533 | 0.356 | 0.965 | 0.964 | 0.612 | 0.816 |
| Bolivia | 0.0515 | 0.0793 | 0.325 | 0.363 | 0.992 | 0.997 | 0.703 | 0.77 |
| Bulgaria | 0.0881 | 0.202 | 0.455 | 0.374 | 0.992 | 0.973 | 0.724 | 0.917 |
| Canada | 0.224 | 0.111 | 0.293 | 0.191 | 0.977 | 0.994 | 0.841 | 0.915 |
| Chile | 0.0728 | 0.0399 | 0.354 | 0.289 | 0.993 | 0.997 | 0.529 | 0.764 |
| Colombia | 0.0901 | 0.0421 | 0.207 | 0.173 | 0.999 | 0.997 | 0.868 | 0.853 |
| CostaRica | 0.057 | 0.064 | 0.464 | 0.332 | 0.999 | 0.996 | 0.372 | 0.717 |
| Croatia | 0.117 | 0.226 | 0.347 | 0.4 | 0.989 | 0.968 | 0.861 | 0.972 |
| Cuba | 0.0414 | 0.163 | 0.341 | 1.16 | 0.998 | 0.958 | 0.583 | 0.35 |
| Cyprus | 0.0795 | 0.441 | 0.452 | 1.32 | 0.99 | 0.781 | 0.649 | 0.257 |
| CzechRepublic | 0.178 | 0.0789 | 0.303 | 0.19 | 0.997 | 0.998 | 0.882 | 0.941 |
| Denmark | 0.0802 | 0.172 | 0.342 | 0.357 | 0.993 | 0.964 | 0.765 | 0.865 |
| DominicanRepublic | 0.0711 | 0.142 | 0.383 | 0.298 | 0.994 | 0.996 | 0.464 | 0.67 |
| Ecuador | 0.0576 | 0.0651 | 0.593 | 0.659 | 0.996 | 0.985 | 0.0681 | 0.153 |
| ElSalvador | 0.103 | 0.155 | 0.521 | 0.304 | 0.997 | 0.995 | 0.363 | 0.78 |
| Estonia | 0.0724 | 0.157 | 0.322 | 0.937 | 0.997 | 0.899 | 0.844 | 0.515 |
| Ethiopia | 0.0365 | 0.0424 | 0.238 | 0.26 | 0.999 | 0.998 | 0.806 | 0.789 |
| Finland | 0.426 | 0.0647 | 0.43 | 0.515 | 0.962 | 0.991 | 0.602 | 0.737 |
| France | 0.325 | 0.895 | 0.577 | 2.16 | 0.919 | 0.794 | 0.376 | 0.265 |
| Germany | 0.233 | 0.0288 | 0.298 | 0.267 | 0.98 | 0.992 | 0.87 | 0.807 |
| Ghana | 0.183 | 0.149 | 0.701 | 1.35 | 0.995 | 0.992 | 0.383 | 0.106 |
| Greece | 0.0696 | 0.0436 | 0.379 | 0.205 | 0.993 | 1 | 0.793 | 0.961 |
| Hungary | 0.0975 | 0.105 | 0.332 | 0.218 | 0.998 | 0.997 | 0.823 | 0.976 |
| Iceland | 0.0558 | 0.241 | 0.405 | 1.87 | 0.996 | 0.887 | 0.737 | 0.0427 |
| India | 0.0316 | 0.0141 | 0.147 | 0.116 | 1 | 1 | 0.915 | 0.929 |
| Indonesia | 0.0867 | 0.0358 | 0.225 | 0.176 | 0.999 | 0.998 | 0.813 | 0.829 |
| Iran | 0.0768 | 0.0476 | 0.306 | 0.207 | 0.989 | 0.997 | 0.689 | 0.72 |
| Iraq | 0.0897 | 0.118 | 0.233 | 0.314 | 0.994 | 0.984 | 0.855 | 0.572 |
| Ireland | 0.161 | 0.209 | 0.403 | 0.353 | 0.971 | 0.966 | 0.571 | 0.934 |
| Israel | 0.113 | 0.16 | 0.653 | 0.453 | 0.968 | 0.979 | 0.336 | 0.395 |
| Italy | 0.356 | 0.019 | 0.336 | 0.185 | 0.965 | 0.998 | 0.873 | 0.951 |
| Jamaica | 0.119 | 0.159 | 0.339 | 0.742 | 0.997 | 0.995 | 0.735 | 0.485 |
| Japan | 0.386 | 0.258 | 0.38 | 0.461 | 0.989 | 0.982 | 0.731 | 0.467 |
| Kazakhstan | 0.0458 | 0.109 | 0.578 | 0.468 | 0.996 | 0.996 | 0.157 | 0.676 |
| Kenya | 0.0498 | 0.0869 | 0.359 | 0.389 | 0.998 | 0.986 | 0.646 | 0.65 |
| Kuwait | 0.0412 | 0.139 | 0.24 | 0.392 | 0.996 | 0.992 | 0.621 | 0.373 |
| Latvia | 0.113 | 0.758 | 0.302 | 0.671 | 0.999 | 0.92 | 0.881 | 0.779 |
| Lithuania | 0.0828 | 0.205 | 0.355 | 0.435 | 0.993 | 0.99 | 0.777 | 0.85 |
| Luxembourg | 0.251 | 0.43 | 0.716 | 0.731 | 0.993 | 0.954 | 0.416 | 0.443 |
| Madagascar | 0.163 | 0.11 | 0.561 | 0.789 | 0.996 | 0.999 | 0.594 | 0.46 |
| Malawi | 0.332 | 0.132 | 0.802 | 0.649 | 0.998 | 0.988 | 0.421 | 0.687 |
| Malaysia | 0.0712 | 0.131 | 0.251 | 0.554 | 0.999 | 0.982 | 0.889 | 0.599 |
| Maldives | 0.0853 | 0.446 | 0.403 | 1.32 | 0.993 | 0.974 | 0.52 | 0.00805 |
| Malta | 0.0713 | 0.373 | 0.397 | 0.763 | 0.993 | 0.97 | 0.704 | 0.559 |
| Mexico | 0.127 | 0.041 | 0.378 | 0.189 | 0.999 | 0.997 | 0.353 | 0.689 |
| Morocco | 0.0627 | 0.0689 | 0.292 | 0.214 | 0.995 | 0.996 | 0.782 | 0.88 |
| Mozambique | 0.032 | 0.0886 | 0.383 | 0.89 | 0.998 | 0.996 | 0.551 | 0.232 |
| Nepal | 0.0732 | 0.236 | 0.335 | 0.378 | 0.993 | 0.972 | 0.685 | 0.639 |
| Netherlands | 0.242 | 0.155 | 0.232 | 0.206 | 0.991 | 0.987 | 0.953 | 0.909 |
| NewZealand | 0.338 | 0.216 | 0.72 | 1.87 | 0.851 | 0.688 | 0.71 | 0.178 |
| Nigeria | 0.494 | 0.116 | 0.603 | 0.428 | 0.987 | 0.988 | 0.275 | 0.571 |
| NorthMacedonia | 0.0823 | 0.0547 | 0.349 | 0.247 | 0.997 | 0.998 | 0.748 | 0.907 |
| Norway | 0.0411 | 0.0825 | 0.375 | 0.623 | 0.995 | 0.975 | 0.646 | 0.715 |
| Pakistan | 0.0998 | 0.025 | 0.362 | 0.276 | 0.982 | 0.998 | 0.611 | 0.777 |
| Panama | 0.0944 | 0.0882 | 0.387 | 0.386 | 0.981 | 0.982 | 0.528 | 0.245 |
| Paraguay | 0.0871 | 0.0511 | 0.229 | 0.177 | 0.999 | 0.998 | 0.863 | 0.925 |
| Peru | 0.319 | 0.0694 | 0.541 | 0.425 | 0.987 | 0.987 | 0.174 | 0.47 |
| Philippines | 0.0588 | 0.0305 | 0.28 | 0.274 | 0.995 | 0.999 | 0.682 | 0.704 |
| Poland | 0.126 | 0.129 | 0.303 | 0.223 | 0.991 | 0.997 | 0.867 | 0.928 |
| Portugal | 0.372 | 0.543 | 0.373 | 0.398 | 0.963 | 0.982 | 0.867 | 0.83 |
| Qatar | 0.164 | 0.239 | 0.347 | 0.916 | 0.983 | 0.974 | 0.829 | 0.306 |
| Romania | 0.0623 | 0.0489 | 0.355 | 0.204 | 0.993 | 0.998 | 0.675 | 0.845 |
| Russia | 0.0284 | 0.0197 | 0.0551 | 0.076 | 1 | 0.999 | 0.989 | 0.983 |
| Rwanda | 0.102 | 0.239 | 0.562 | 1.39 | 0.99 | 0.979 | 0.412 | 0.161 |
| SaudiArabia | 0.0669 | 0.0821 | 0.39 | 0.243 | 0.988 | 0.992 | 0.683 | 0.755 |
| Senegal | 0.0494 | 0.0601 | 0.353 | 0.664 | 0.995 | 0.991 | 0.545 | 0.333 |
| Serbia | 0.281 | 0.177 | 0.548 | 0.558 | 0.909 | 0.928 | 0.916 | 0.833 |
| Singapore | 0.199 | 2.25 | 0.68 | 3.64 | 0.966 | 0.837 | 0.4 | 0.0197 |
| Slovakia | 0.155 | 0.0833 | 0.386 | 0.367 | 0.994 | 0.995 | 0.755 | 0.821 |
| Slovenia | 0.289 | 0.582 | 0.423 | 0.617 | 0.994 | 0.931 | 0.818 | 0.83 |
| SouthAfrica | 0.196 | 0.117 | 0.282 | 0.21 | 0.99 | 0.998 | 0.842 | 0.909 |
| SouthKorea | 0.0722 | 0.136 | 0.346 | 0.599 | 0.987 | 0.969 | 0.849 | 0.409 |
| Spain | 0.22 | 0.167 | 0.418 | 0.344 | 0.986 | 0.985 | 0.672 | 0.733 |
| SriLanka | 0.0977 | 1.17 | 0.361 | 0.963 | 0.996 | 0.784 | 0.809 | 0.567 |
| Sweden | 0.377 | 0.121 | 0.808 | 0.397 | 0.546 | 0.967 | 0.00298 | 0.669 |
| Switzerland | 0.137 | 0.347 | 0.528 | 0.414 | 0.996 | 0.993 | 0.608 | 0.866 |
| Thailand | 2.92 | 5.44 | 2.19 | 5.7 | 0.881 | 0.994 | 0.151 | 0.621 |
| Togo | 0.13 | 0.163 | 0.561 | 1.54 | 0.991 | 0.985 | 0.176 | 0.0396 |
| Tunisia | 0.687 | 0.821 | 0.844 | 0.92 | 0.963 | 0.907 | 0.329 | 0.618 |
| Turkey | 0.212 | 0.0809 | 0.637 | 0.243 | 0.852 | 0.98 | 0.106 | 0.909 |
| UAE | 0.122 | 0.195 | 0.408 | 0.382 | 0.986 | 0.981 | 0.534 | 0.484 |
| UK | 0.292 | 0.282 | 0.383 | 0.283 | 0.976 | 0.966 | 0.767 | 0.839 |
| Ukraine | 0.0565 | 0.0664 | 0.237 | 0.19 | 0.996 | 0.996 | 0.84 | 0.896 |
| Uruguay | 0.0481 | 0.0878 | 0.276 | 1.08 | 0.993 | 0.988 | 0.952 | 0.366 |
| USA | 0.0716 | 0.0306 | 0.23 | 0.146 | 0.991 | 0.999 | 0.936 | 0.857 |
| Zambia | 0.0841 | 0.106 | 0.725 | 0.808 | 0.994 | 0.984 | 0.258 | 0.386 |

Figure S6 shows the visualization of fit between data and simulations for all the countries in our sample. These graphs include data and model outputs for reported new cases (blue; left axis in thousands per day) and deaths (red; right axis in thousands per day) starting from the beginning of the epidemic in each country until 22 December 2020.

**Figure S6.**
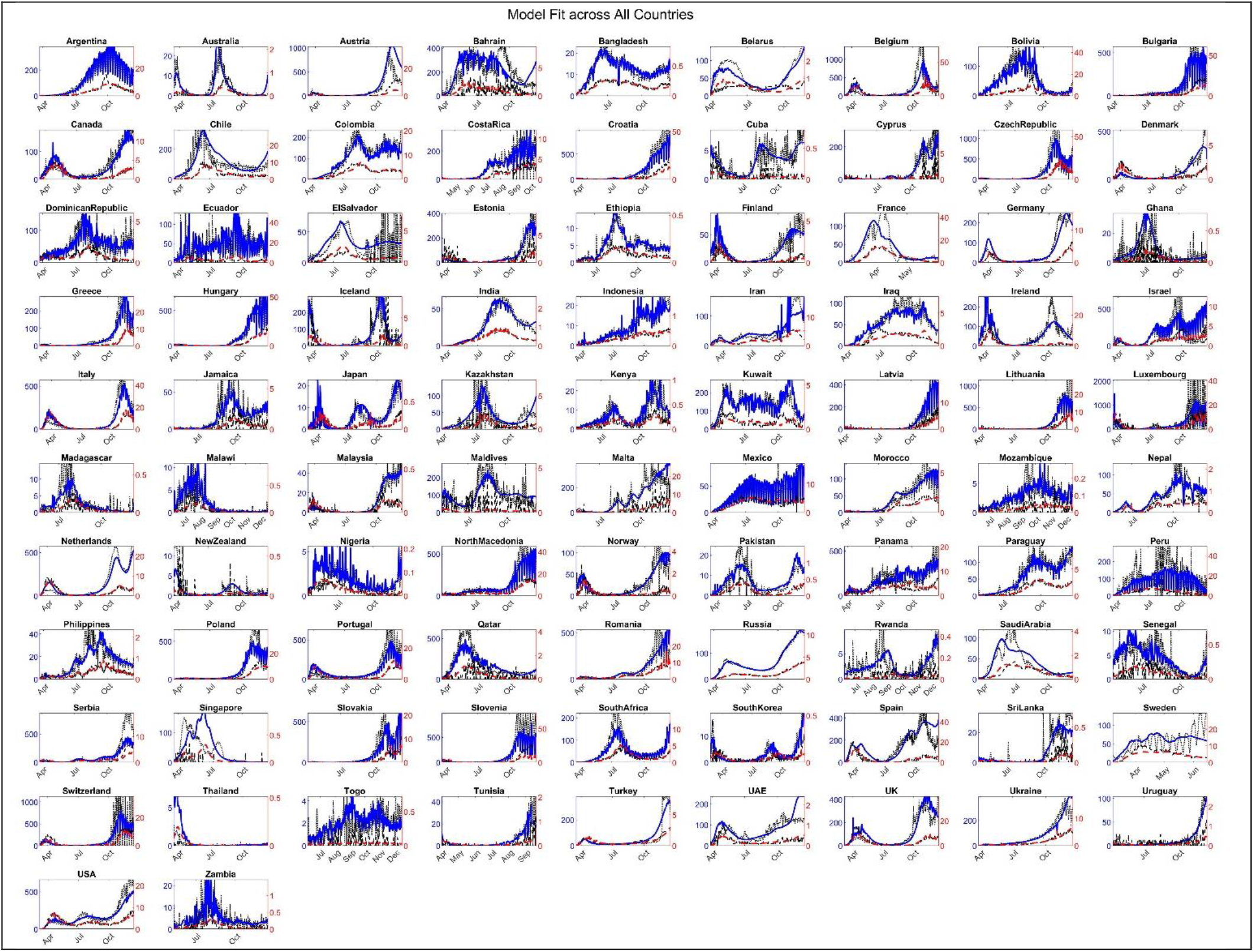
Comparison of data and simulation. New cases in blue (left axis, in thousands per day) and new deaths (red, right axis).

### Estimates for true magnitude of epidemic

Estimates for true cumulative cases (blue; left axis in millions) and deaths (red; right axis in thousands) across different countries up to 22 December 2020 are reported in Figure S7.

**Figure S7.**
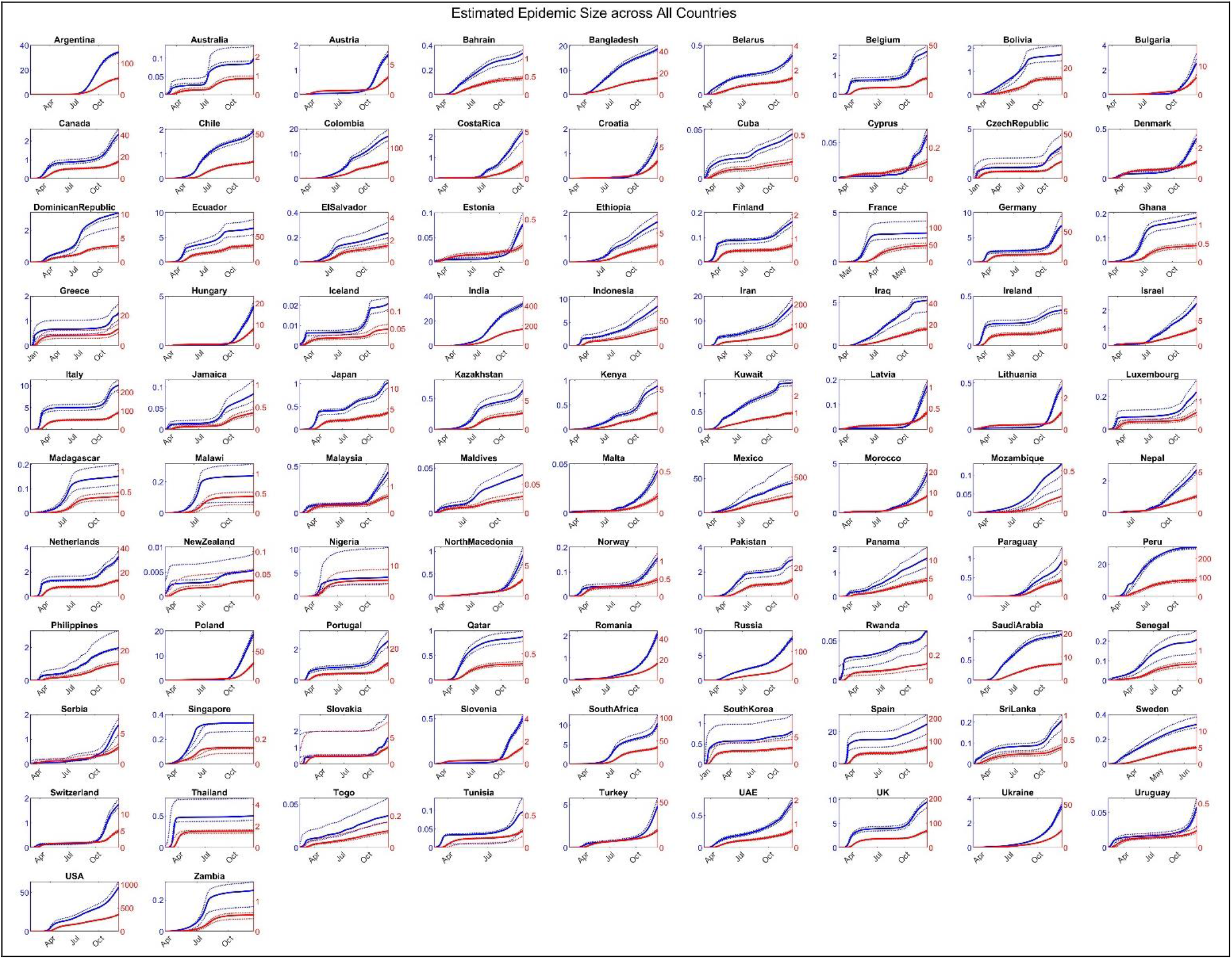
Estimates and 95% credible intervals for true magnitude of epidemic. Cumulative cases (blue, left axis in millions) and cumulative deaths (red, right axis, in thousands)

### Excess deaths

Figure S8 shows the ratio of estimated excess deaths, i.e. COVID-19 fatalities not reported as such, to reported excess deaths, i.e. deaths over historical baseline not accounted for by reported COVID-19 deaths, for the countries for which such data are available.

**Figure S8.**
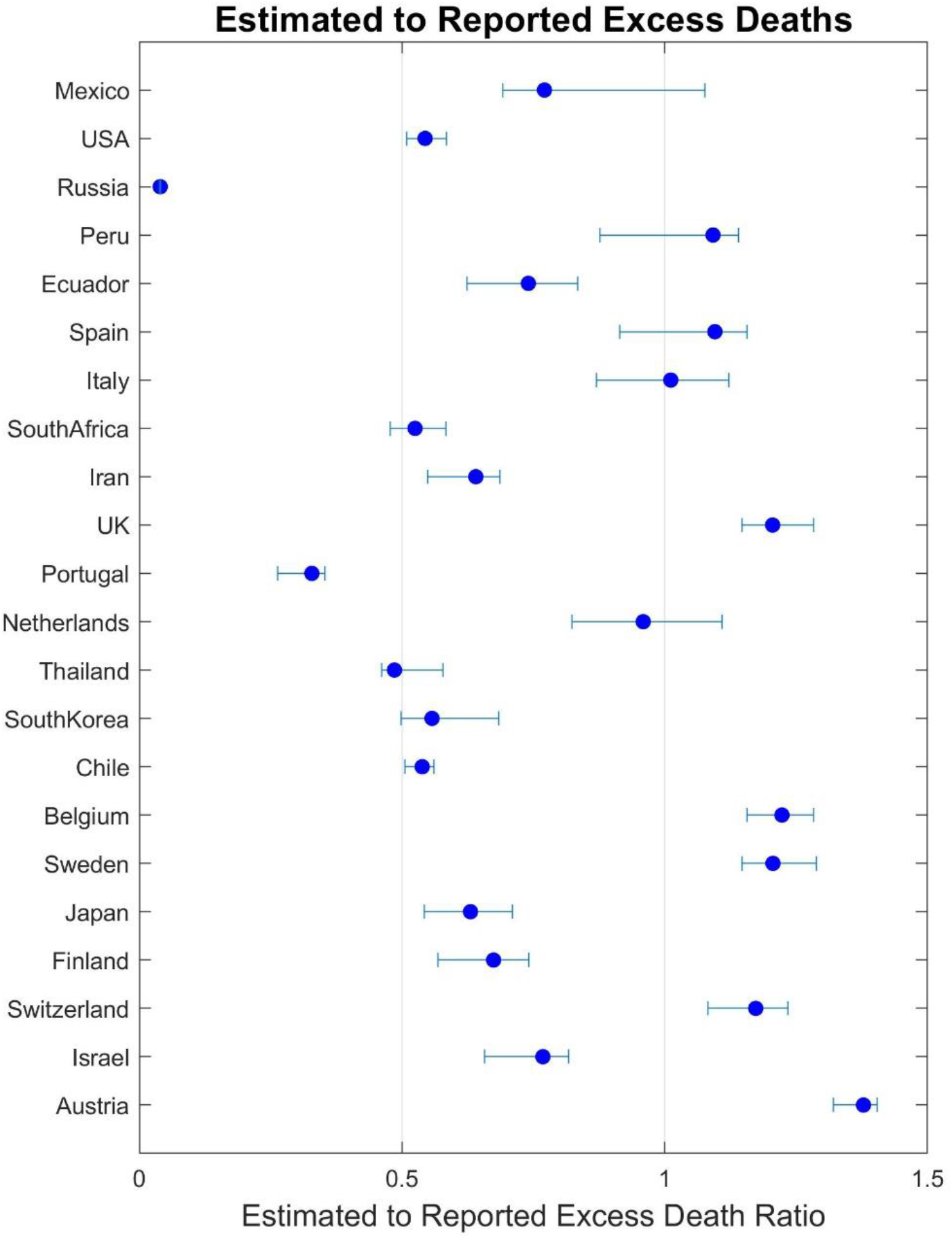
Ratio of estimated excess deaths to reported excess deaths for countries for which this data was available.

### Maximum reproduction number

*Figure S9* shows the initial reproduction number (R_E_) occurring in each country. Reproduction numbers are changing dynamically and transient dynamics may lead to larger than equilibrium numbers if maximum R_E_ values were used. We therefore use the 90^th^ percentile of simulated reproduction number in this graph. Also note the large credible intervals for these estimates. This range is partly driven by what exact point of the curve is represented by the 90^th^ percentile. It is also due to the inherent uncertainty when both reproduction number and behavioral and policy responses are estimated: one can have smaller initial R_E_ and smaller response functions, or larger values for both, and stay consistent with the data, specially because early in the epidemic ascertainment rates are very low and data is not very informative about the true magnitude. Moreover, given the recording of R_E_ values at their (often initial) high values, they may reflect non-representative subpopulations or events. For example the high impact of weather conditions on transmission rates could notably alter Maximum R_E_ for some countries depending on weather conditions at the time of first wave.

**Figure S9.**
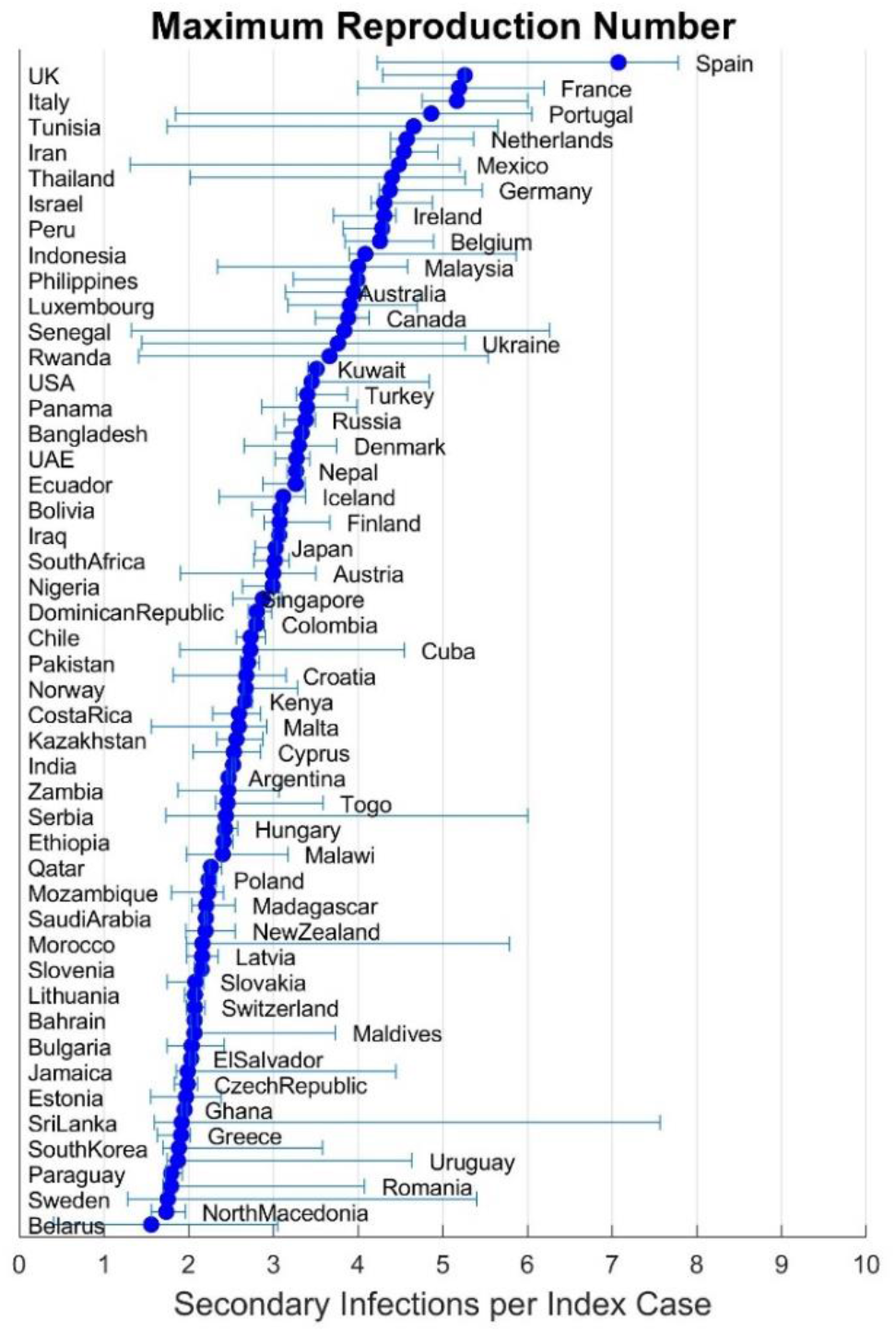
Maximum reproduction number R_E_ for each country’s outbreak

### Time to herd immunity

Figure S10 shows estimated time to herd immunity across nations. These estimates are based on time it takes before 60% of population has been infected by COVID-19. Depending on the basic reproduction number in each location and the heterogeneity in contacts, the 60% threshold will not be an exact value for most countries, but offers a reasonable intuition for the ranges of time involved and could be adjusted with a linear scaling to other thresholds. Two estimates are offered, one for the estimated number based on current true infection rates, and another based on the peak infection rates experienced in that country. The two may be the same if the current rates are the peak rates.

**Figure S10.**
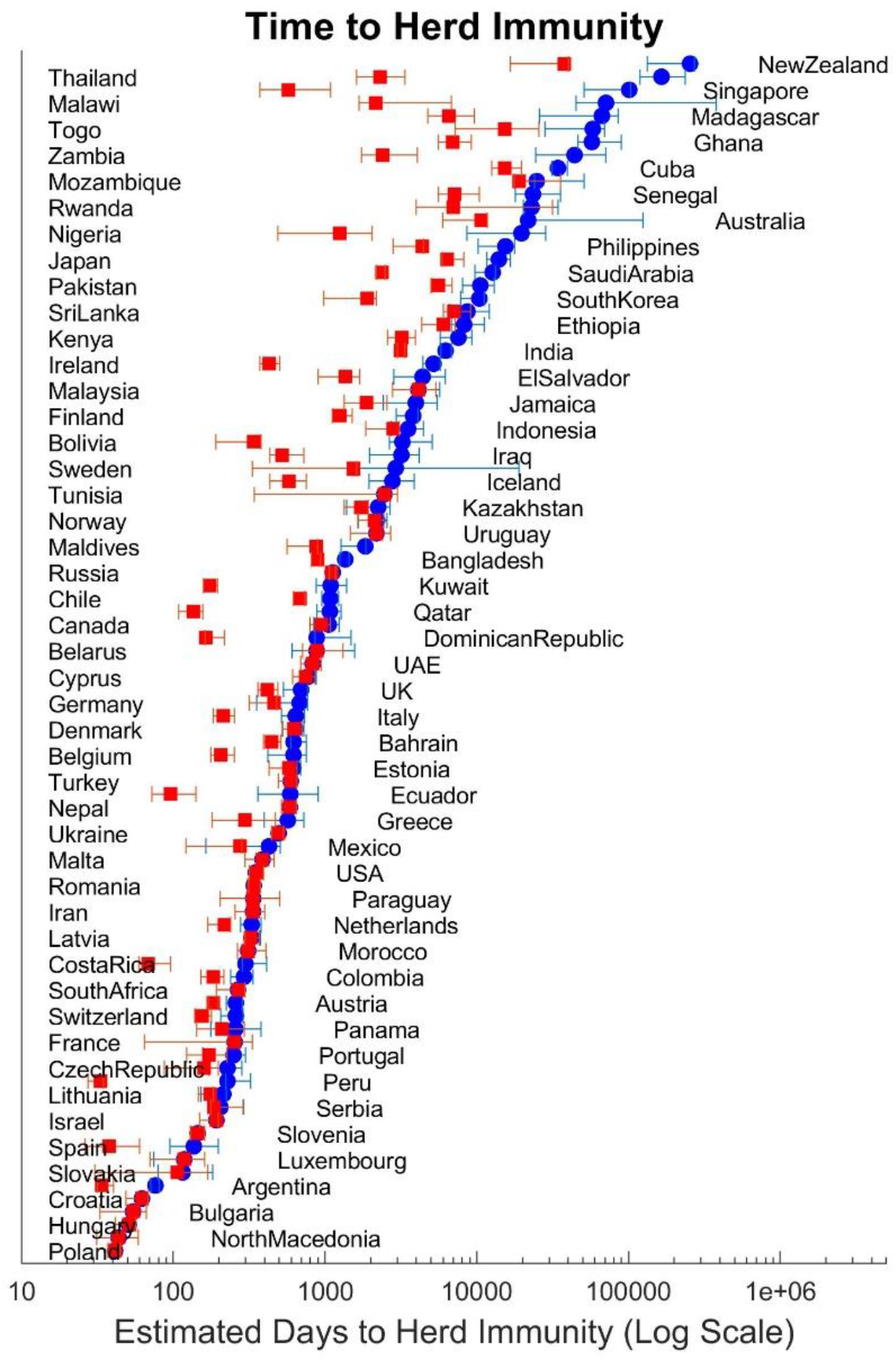
Time to 80% cumulative infection based on current infection rates (blue circles) and peak infection rates to-date (red squares) in days (log scale).

### Parameter estimates

Figure S11 reports most likely estimates for the vector of country-specific parameters (***θ***_***i***_). The figure also includes 95% credible intervals for these parameter estimates.

**Figure S11.**
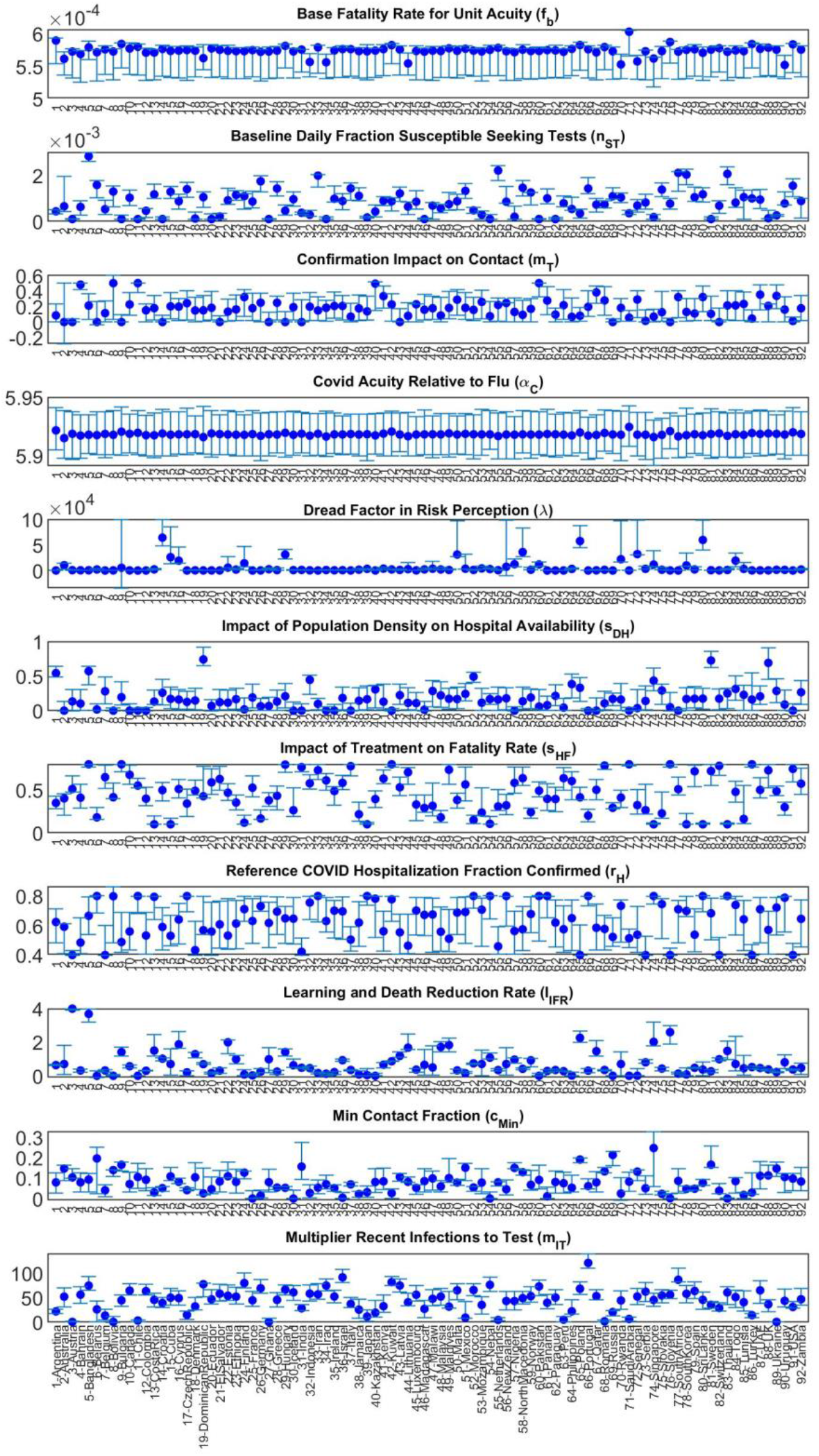
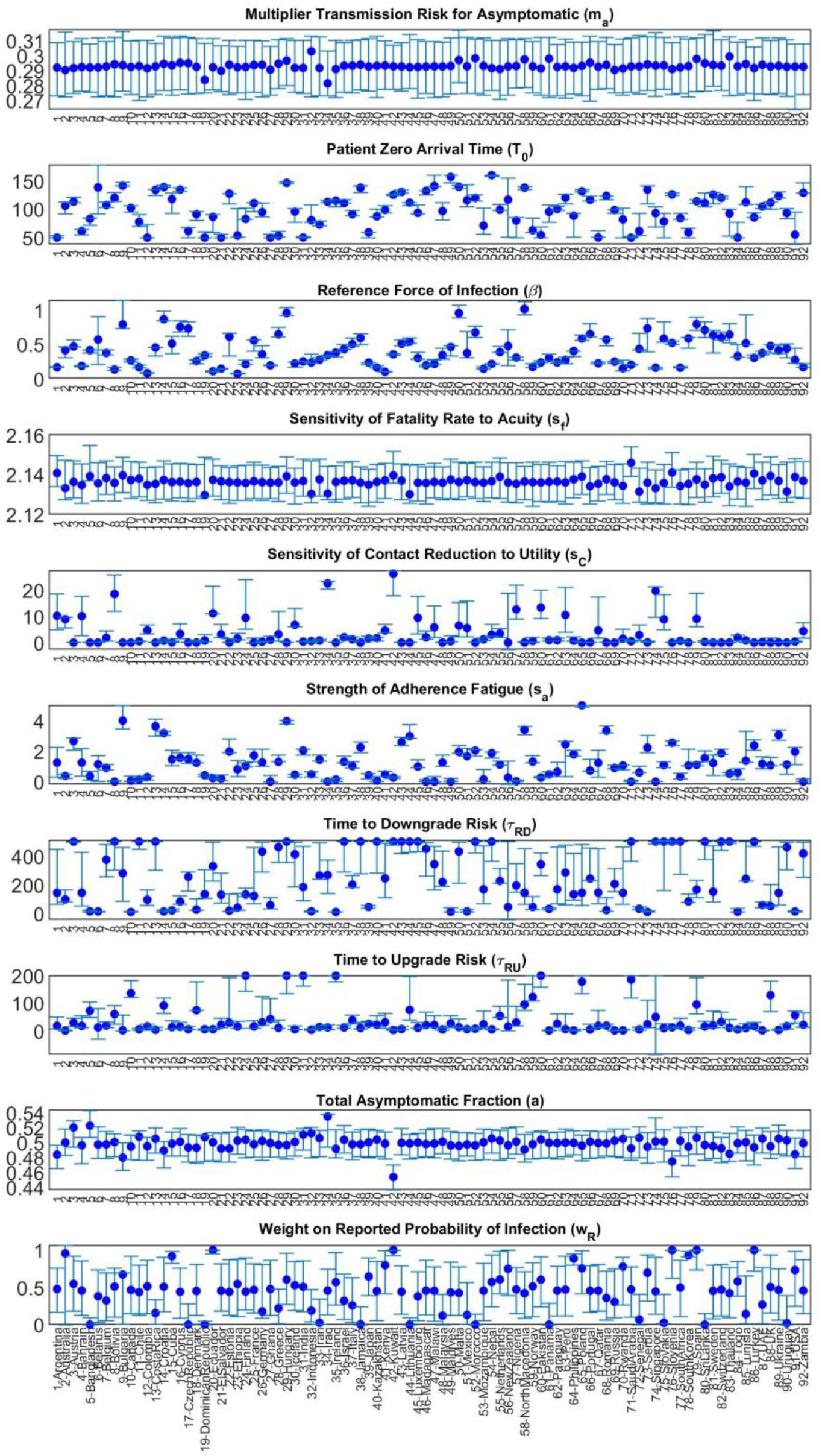
Parameter estimates and 95% credible regions for country-specific parameters.

### Future projections

Figure S12 reports country-level projections for new cases and deaths based on the scenario I (no changes in estimated parameters; no vaccination; testing fixed at values observed for 22 December 2020; consistent with those reported in Figure 7 in the main paper).

In scenarios II and IV in the main paper (not shown here) we change responsiveness through:

- Increasing *Time to Downgrade Risk* (*σ*_*RD*_) by 20%
- Shifting *Sensitivity of Contact Reduction to Utility* (*s*_*C*_) by 20%.
- Increasing *Dread Factor in Risk Perception* (*λ*) by 20%.

Note that these changes primarily change contacts as a function of perceived risk, but do not necessarily entails fewer contacts overall. Vaccination scenario setups are discussed in vaccination sector (under S1).

**Figure S12.**
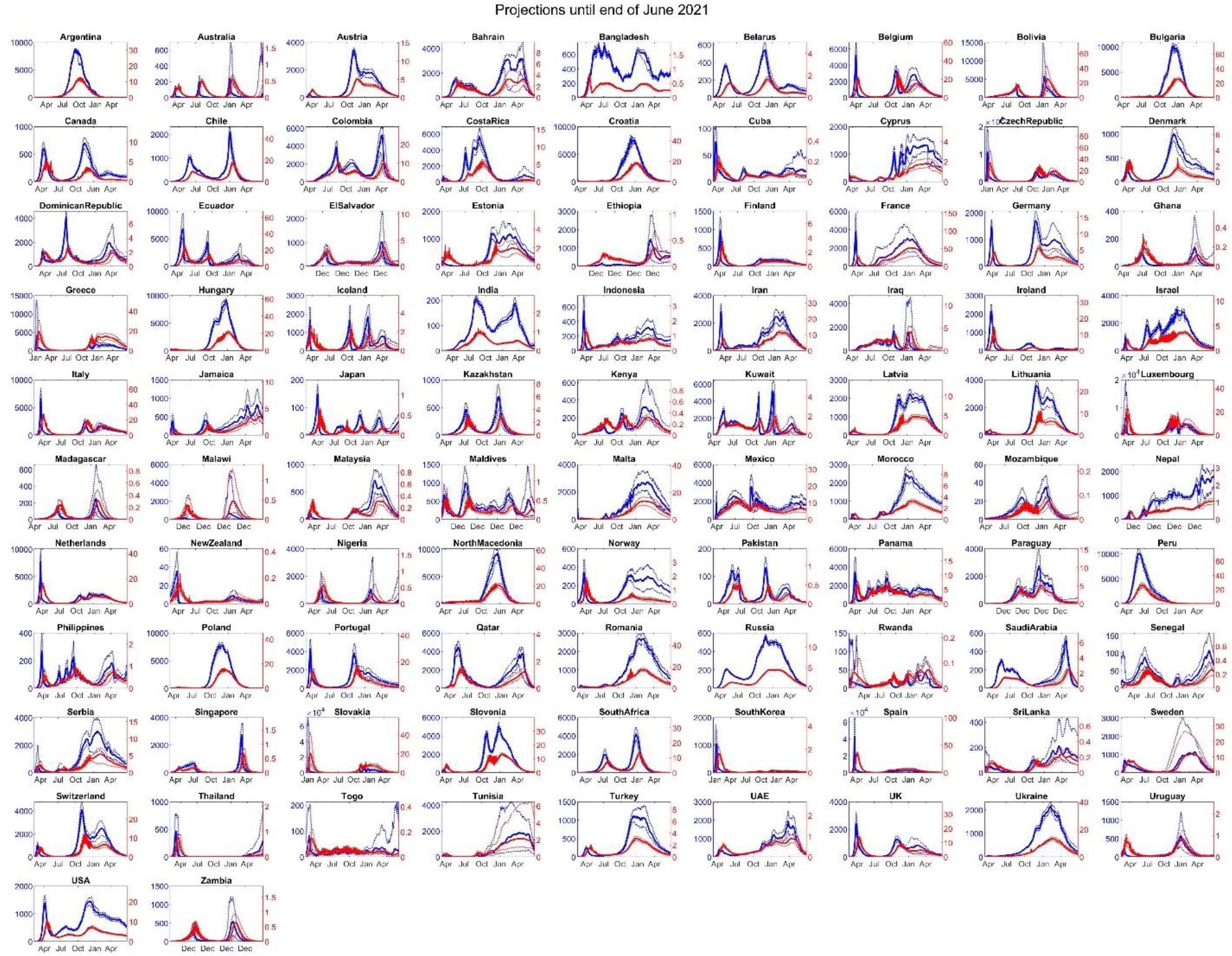
Country level projections until Spring 2021 in scenario I. Daily cases (in thousands, blue, left axis) and daily deaths (red, right axis) are graphed.

## S6 OUT OF SAMPLE PREDICTION TEST

We conducted an out of sample prediction test of the model by comparing the quality of fit for model projections for future data not used in model estimation against the version of the model using that data. We calculate the quality of fit for projections of that model (the “early model”) for data later released for the period 30 September 2020-22 December 2020. Those projections are reported in Figure S13 and are directly comparable with Figure S6. Note that we use the actual testing rates to drive the model for this prediction interval. Inspection of this graph points to various outcomes across countries, ranging from close fit for the prediction interval to a few with major discrepancies. For example, the model was able to predict the emergence of a second wave, before it was detectable in the infection data, for Belarus, Russia, and UK and the model predicted the third waves in Iran, Israel and USA well. On the other hand, among others, we overestimated the Fall trajectory of epidemic in India and under-estimated that in Turkey.

The discrepancies arise from both the baseline gaps between the model and data and emerging features of the epidemic in the prediction interval. The baseline gaps typically appear because our method enforces a strong coupling among countries. For instance, keeping IFR parameters similar across countries, the model cannot explain the unexpectedly low fatality rates in Qatar and Singapore (which could be due to outbreaks among younger immigrant worker communities). Moreover, the model is unable to accommodate changes in responses that are not detectable in the historical data (e.g. missing the magnitude of the “second wave” when first wave is not yet complete and thus the parameters for risk perception and response are not fully identifiable).

**Figure S13.**
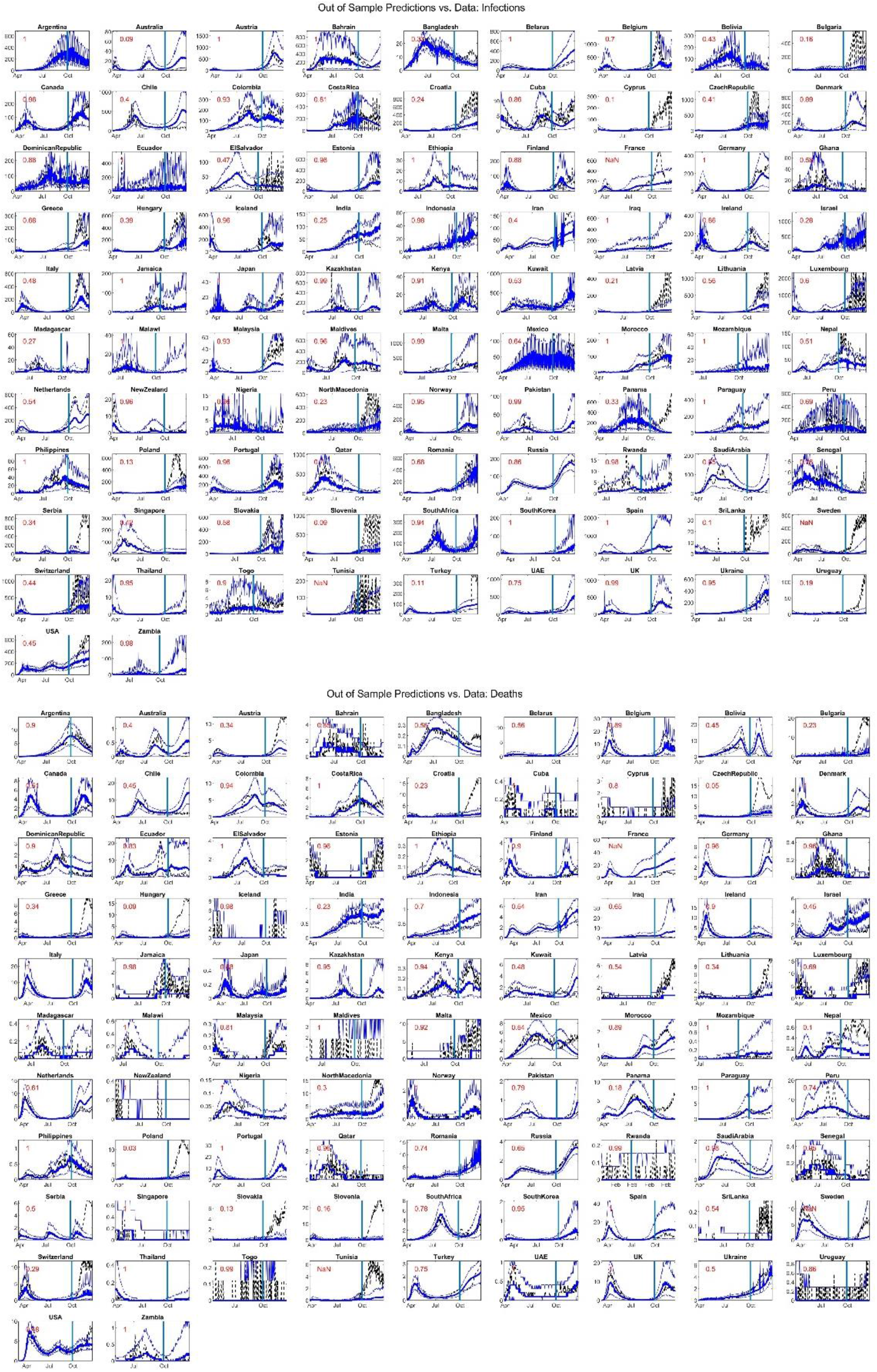
Predictions from the model fitted with data until 30 September 2020 for the 30 September 2020-22 December 2020 interval. The start of prediction interval is marked with a horizonal blue line. Red numbers represent the fraction of data falling within the 95% prediction intervals.

Assessing the quality of fit requires some benchmark to compare against. Defining external benchmarks in the case of current model is complicated because, to our knowledge, no other model has attempted to simultaneously match infection and fatality data across this large set of nations and offer future projections. Therefore, we focus on an internal benchmark using quality of fit for the version of the model using the data from the prediction interval for estimation. Specifically, we compare the fit measures from the early model (MAEN scores for reported infection rates and death rates) against the same fit measures coming from the model estimated until 22 December 2020. The updated version uses the data in the prediction interval and thus is likely to have a better fit than the early version of the model. The ratio of MAEN values between these two models informs the speed with which fit quality deteriorates, with values closer to one suggesting robust long-term predictions. Overall the mean fit ratio is 0.516, loosely speaking suggesting that projections lose their accuracy by about a half over 1.5 months. Table S4 reports those values across different nations.

**Table S4.**
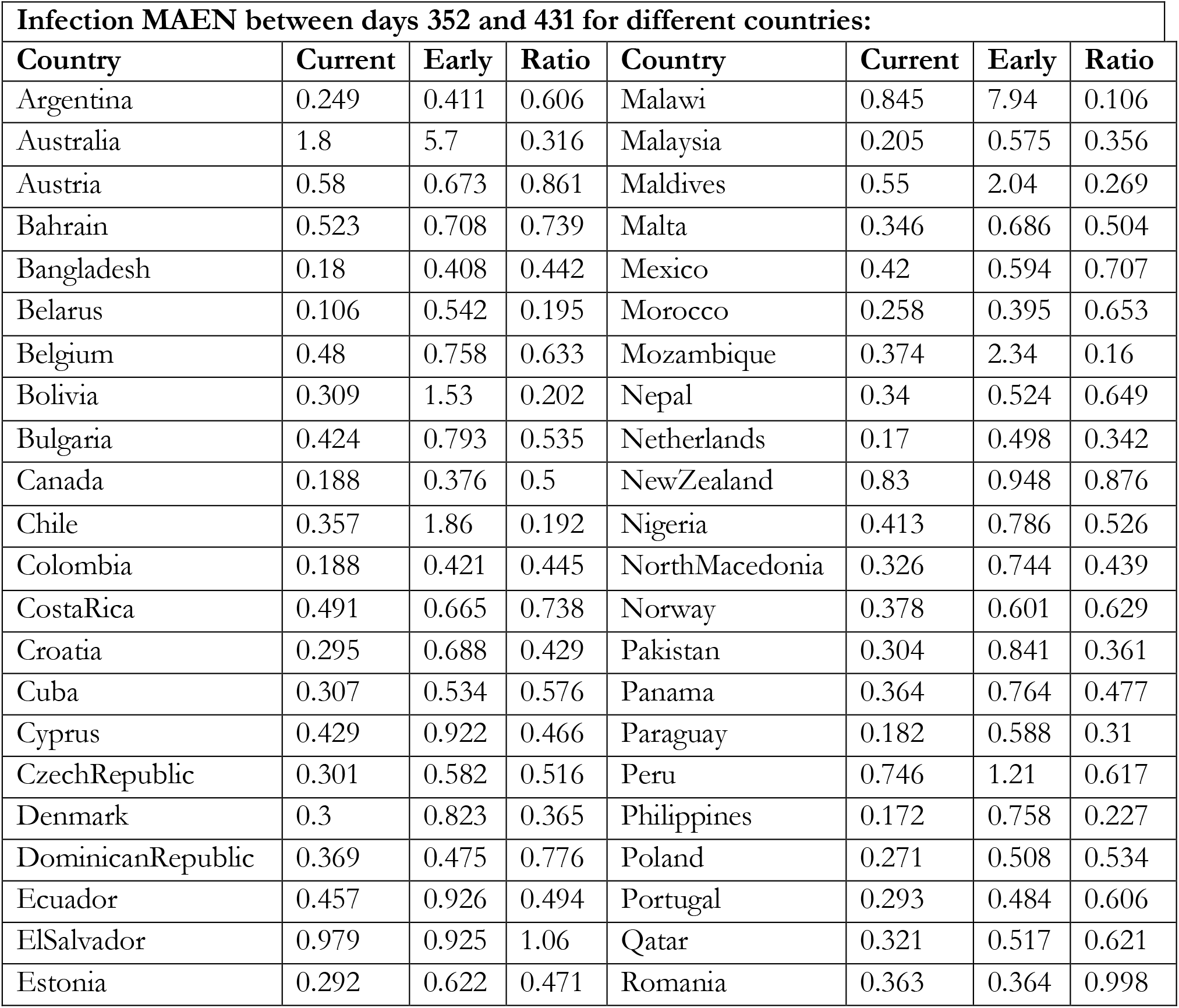
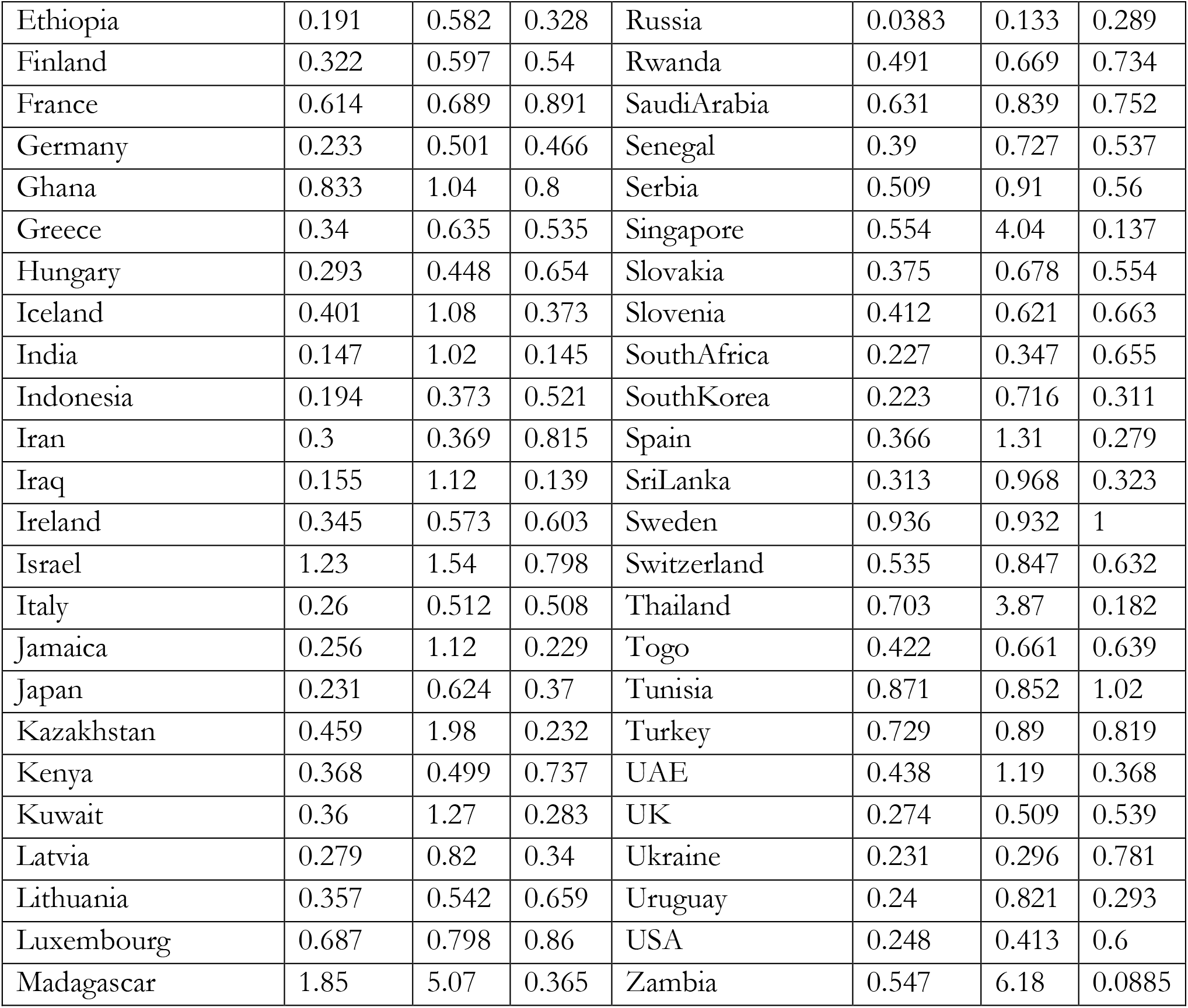
Quality of fit for future projections compared to when data is available. Mean Absolute Error Normalized by Mean (MAEN) for daily infection rates in the early model, current model (which uses data from prediction interval of 30 September 2020-22 December 2020), and the ratio of current model’s MAEN to the early one.

## S7 SENSITIVITY ANALYSIS

### Impact of Cross-country Parameter Variances

#### Setup

Our estimation method uses a random effects framework, which couples the parameters across different countries, specifying them as instances of some underlying distribution with a given variance. Those variance factors, explained in S2, were specified to incorporate the authors’ judgement on how different each parameter may be across different countries, based on the nature of those parameters. For instance, parameters representing physiological or virological constructs should generally vary less than those representing socio-cultural and behavioral responses. In principle, one could propose other variance factors. Assuming very large variances would essentially decouple the models for different countries, while shrinking variances towards zero will force all parameters to be the same across countries. In this section we assess the sensitivity of results to changes in those variance factors. Specifically, we re-do the analysis when all variance factors are scaled by a factor of 4, or 0.25. We re-estimate the model in each case and measure how much the following 12 outcome measures (organized into 3 groups) change compared to the baseline estimates as a result:

a. Country level projections for 1) Actual to reported case ratio; 2) Actual to reported death ratio; 3) Projected infection rate at the end of Winter 2021; 4) Projected death rate at the end of winter 2021;
b. Country level MAEN values for daily infection rates and death rates;
c. Aggregate (across all countries) cumulative infections, deaths, and IFR, both on 22 December 2020 and at the end of June 2021.

The first two groups of measures are country specific, so we report them for all countries, followed by their averages, and then the aggregate outcomes.

#### Results

In Table S5 and Table S6 results from these two experiments are reported. As expected, increasing allowed variances enables the model to offer a better fit to data (i.e. reduces MAEN values, thus mostly negative values for fit statistics in the first experiment). Other sensitivities remain relatively small for most countries, showing few systematic changes in model’s predictions as in response to changes in the cross-country parameter variances. However, trajectories for a handful of countries are sensitive to these variance factors: Australia, Bolivia, Bulgaria, Costa Rica, Croatia, El Salvador, Ghana, Hungry, Iraq, Maldives, Mozambique, New Zealand, North Macedonia, Paraguay, Poland, Portugal, Qatar, Rwanda, Singapore, Slovakia, Thailand, Togo. In these cases one would expect a separate country-specific estimation to give results that may be qualitatively different from those we find, and thus caution should be exercised.

**Table S5.** Impact of increasing assumed cross-country parameter variances by a factor of four. All reported outcomes measure percentage change in the given metric from baseline analysis.

| Sensitivities to 4x change in variances |  |  |  |  |  |  |
| --- | --- | --- | --- | --- | --- | --- |
| Country | Case Undercount Ratio | Death Undercount Ratio | Final Infection Rate | Final Death Rate | MAEN Infection | MAEN Death |
| Argentina | -2.4 | 1.51 | -27.8 | -16.3 | -0.598 | -2.66 |
| Australia | 6.54 | 0.789 | -50.3 | -42 | 2.49 | -1.59 |
| Austria | 2.63 | 0.651 | -2.88 | -1.32 | 2.45 | -0.305 |
| Bahrain | 5.76 | -0.985 | -5.13 | -10 | -4.97 | -0.321 |
| Bangladesh | 6.1 | 4.42 | 3.17 | 3.65 | -1.54 | -0.712 |
| Belarus | -1.66 | -3.49 | -18.4 | -37.2 | -13 | -16 |
| Belgium | 4.09 | 2.05 | -13.9 | -14.6 | 4.32 | -6.1 |
| Bolivia | 5.57 | 4.14 | 19.1 | 9.16 | -0.33 | -0.893 |
| Bulgaria | 8.34 | 11.9 | -66 | -46.2 | -3.14 | -22.2 |
| Canada | 6.7 | 1.05 | 15 | -4.68 | -1.15 | -3.25 |
| Chile | 0.313 | 0.54 | -0.0162 | 0.796 | 1.16 | 0.591 |
| Colombia | 5.15 | 3.15 | 0.141 | 2.24 | 4.86 | -1.09 |
| Costa Rica | -0.241 | -0.176 | -3.48 | -4.25 | 0.194 | -0.298 |
| Croatia | -11.8 | 1.1 | 38.1 | 0.771 | -7.22 | -13 |
| Cuba | 7.23 | 6.38 | 87.6 | -1.47 | -1.66 | 0.218 |
| Cyprus | -1.57 | 3.07 | 10.7 | 23.8 | 1.09 | 0.377 |
| Czech Republic | 7.3 | 1.7 | -1.66 | -11.8 | -4.06 | -3.12 |
| Denmark | -1.15 | 1.15 | -7.63 | -2.69 | 0.428 | -2.93 |
| Dominican Republic | 3.83 | 2.53 | -3.73 | -3.39 | -3.87 | 1.22 |
| Ecuador | -2.44 | -0.457 | 0.773 | 6.77 | -1.21 | -0.779 |
| El Salvador | 13.7 | 3.6 | 382 | 113 | -3.69 | -5.77 |
| Estonia | -1.18 | 4.28 | 8.43 | 3.91 | 0.817 | -0.0563 |
| Ethiopia | 18.4 | 8.4 | 19.4 | 34.8 | -0.683 | -4.19 |
| Finland | 2.53 | -0.747 | 13.9 | 8.82 | -9.07 | -2.22 |
| France | 8.58 | 0.654 | -21.3 | -25.6 | -3.83 | -2.8 |
| Germany | 13 | 4.94 | 8.23 | -1.72 | -1.53 | -6.32 |
| Ghana | 0.0961 | -2.99 | 883 | 111 | -1.21 | -1.51 |
| Greece | 10.6 | 6.14 | -6.19 | -9.94 | 2.45 | -4.32 |
| Hungary | 7.05 | 8.15 | -15.8 | -11.7 | -6.2 | -17.9 |
| Iceland | 2.05 | 0.375 | -22.8 | -9.36 | -1.8 | 0.348 |
| India | -1.16 | 0.507 | -5.71 | -2.22 | -0.93 | -1.02 |
| Indonesia | 10 | 1.42 | 33.7 | 1.88 | -0.47 | -3.17 |
| Iran | 6.56 | 1.76 | -7.62 | -13.2 | -0.223 | 0.033 |
| Iraq | -0.25 | -0.115 | -78.7 | 20 | -0.507 | -0.305 |
| Ireland | 7 | 6.63 | 89.5 | 43.8 | 2.51 | -11.6 |
| Israel | 9.8 | 3.51 | -26.9 | -26.5 | -7.7 | -0.895 |
| Italy | 5.97 | -0.44 | -1.51 | -8.66 | -6.67 | -8.15 |
| Jamaica | 3.13 | 0.218 | -21.5 | -14.6 | -6.78 | -0.978 |
| Japan | -0.977 | -1.84 | 8.04 | -5.16 | 2.87 | -0.776 |
| Kazakhstan | -0.859 | 0.344 | 15.9 | 4.75 | -0.22 | -3.47 |
| Kenya | -3.65 | -0.158 | -20.8 | -5.16 | -0.158 | -2.52 |
| Kuwait | 2.05 | 3.51 | -48.2 | -45.2 | -3.62 | -0.351 |
| Latvia | 5.58 | 4.18 | -7.61 | -14.6 | -4.86 | -3.08 |
| Lithuania | 1.5 | 0.727 | -3.58 | 2.33 | 0.646 | -2.17 |
| Luxembourg | 5.02 | 3.85 | 7.44 | -7.68 | -1.84 | -3.58 |
| Madagascar | -9.76 | -11.6 | 17.9 | 28.1 | 1.94 | -0.36 |
| Malawi | -4.34 | -2.08 | 40.9 | -14.7 | -2.95 | -9.22 |
| Malaysia | 10.3 | -1.89 | -1.39 | 0.931 | -3.8 | -3.55 |
| Maldives | -2.84 | 5.95 | 6.46 | 18 | -2.01 | 1.42 |
| Malta | 8.72 | -0.132 | -3.44 | 0.666 | -8.25 | -3.44 |
| Mexico | 8.31 | -2.17 | 1.96 | -15 | -0.358 | -1.11 |
| Morocco | 5.98 | 2.1 | -5.88 | -6.73 | -0.46 | -1.18 |
| Mozambique | -20.1 | -10.1 | -65.2 | -39.3 | -3.78 | -0.717 |
| Nepal | 5.87 | 4.08 | 4.76 | -7.05 | -2.08 | -3.19 |
| Netherlands | -0.153 | 3.17 | -4.7 | -15.2 | -17.4 | -3.14 |
| NewZealand | 2.52 | -13.6 | -24.4 | -29.2 | -5.02 | -7.2 |
| Nigeria | 2.96 | 1.02 | -27.6 | -28.2 | 0.401 | 0.138 |
| NorthMacedonia | 2.42 | 3.39 | -14.6 | -3.5 | -3.11 | -3.8 |
| Norway | 1.6 | 1.58 | -11 | -9.97 | 0.0685 | -2.2 |
| Pakistan | 14.8 | 3.45 | 18.9 | -0.586 | 0.482 | -5.33 |
| Panama | 1.36 | 0.608 | 8.6 | 5.81 | -1.02 | -2.92 |
| Paraguay | 28.4 | 3.56 | -45.4 | -63.6 | 1.4 | -2.88 |
| Peru | -0.539 | -1.06 | 27.7 | 30 | 0.736 | 0.308 |
| Philippines | -1.03 | -0.0881 | 24.5 | 30 | -0.0283 | -1.88 |
| Poland | 3.48 | 9.08 | -20.9 | -21.4 | 3.81 | -15.6 |
| Portugal | 24.3 | -0.662 | -49.4 | -66.7 | 2 | -29.3 |
| Qatar | 0.149 | 3.89 | -50.7 | -32.9 | -9.35 | 3.78 |
| Romania | 0.458 | 0.485 | 5.1 | 3.72 | 1.87 | -2.34 |
| Russia | -0.149 | 0.186 | -7.83 | -6.79 | -3.05 | -0.208 |
| Rwanda | 4.32 | -0.00907 | -34.6 | -29.5 | -0.834 | -0.494 |
| SaudiArabia | -6.25 | 4.24 | -4.14 | 4.87 | -3.88 | 2.26 |
| Senegal | 2.66 | 1.03 | 5.4 | 2.5 | -1.32 | -0.238 |
| Serbia | 17.9 | 7.21 | -29 | -20.5 | 2.59 | -4.21 |
| Singapore | 13.1 | 6.47 | 344 | 167 | -39.2 | -8.7 |
| Slovakia | 9.65 | 0.864 | -44.2 | -50.3 | -0.368 | -3.87 |
| Slovenia | 1.57 | 8.69 | -4.1 | 7.36 | -3.13 | -4.98 |
| SouthAfrica | 11.9 | 3 | -14.8 | -11.3 | 1.34 | -6.72 |
| SouthKorea | 25.2 | 3.89 | -12.8 | -13.9 | -2.6 | -5.92 |
| Spain | 3.52 | 0.503 | -9.22 | 1.67 | -4.95 | -2.09 |
| SriLanka | 8.71 | 9.87 | 21 | 18.5 | -2.96 | -1.03 |
| Sweden | -5.71 | 0.964 | 12.3 | 12.9 | 1.56 | 2.37 |
| Switzerland | -4.17 | 2.43 | -0.919 | 5.64 | 0.00177 | -3.64 |
| Thailand | 17.8 | -2.4 | 100 | 131 | -30.5 | -40.4 |
| Togo | -8.88 | -4.63 | 189 | -32.9 | -3.3 | -0.166 |
| Tunisia | 40.9 | 21.2 | 72.7 | 18.8 | -3 | -1.88 |
| Turkey | 3.9 | -5.47 | -6.91 | -42.2 | 0.335 | -5.23 |
| UAE | 3.35 | 0.36 | -25.6 | -31.4 | -17.4 | -10 |
| UK | -0.541 | 2.77 | 4.79 | -1.36 | -4.06 | -14.8 |
| Ukraine | 1.53 | 0.371 | -3.97 | -19 | -1.38 | -1.23 |
| Uruguay | 0.629 | 0.0577 | -10.4 | -1.42 | 0.927 | -0.185 |
| USA | -1.65 | 1.15 | -2.69 | 0.315 | -6.02 | -0.526 |
| **Average** | **4.31** | **1.78** | **16.9** | **-1.49** | **-2.7** | **-3.98** |
| **Global Percentage Changes and Elasticities** |  |  |  |  |  |  |
|  | **Cases Early** | **Deaths Early** | **IFR Early** | **Cases Proj.** | **Deaths Proj.** | **IFR Proj.** |
| Global | 3.31 | 0.892 | -2.14 | 3.55 | -0.917 | -4.41 |

**Table S6.** Impact of decreasing assumed cross-country parameter variances by a factor of four. All reported outcomes measure percentage change in the given metric from baseline analysis.

| Sensitivities to 0.25x change in variances |  |  |  |  |  |  |
| --- | --- | --- | --- | --- | --- | --- |
| Country | Case Undercount Ratio | Death Undercount Ratio | Final Infection Rate | Final Death Rate | MAEN Infection | MAEN Death |
| Argentina | 0.0032 | -1.36 | 81 | 72.6 | 0.919 | -0.0141 |
| Australia | 3.01 | -2.07 | 62 | 40.5 | -0.812 | 1.58 |
| Austria | -4.18 | -4.18 | 17.4 | 4.89 | -2.17 | 4.15 |
| Bahrain | -4.72 | -1.82 | 7.7 | 9.64 | 3.71 | -0.432 |
| Bangladesh | -8.91 | -3.49 | -4.02 | -4.68 | 19.4 | 12.1 |
| Belarus | 0.182 | 5.85 | 14.4 | 13.5 | 0.671 | 15.2 |
| Belgium | -2.66 | -5.38 | 13.8 | 5.09 | -5.81 | 7.07 |
| Bolivia | -5.01 | -1.56 | -52 | -26 | -1.35 | 7.59 |
| Bulgaria | -7.48 | -12.2 | 404 | 73.1 | 4.82 | 32.5 |
| Canada | -8.34 | -2.11 | -5.7 | 15.1 | -0.343 | 19.7 |
| Chile | 0.638 | -1.1 | -3 | -2.84 | -0.304 | -0.582 |
| Colombia | -7.24 | -3.38 | -1.81 | -4.02 | -6.18 | 0.364 |
| CostaRica | -8.21 | -5.22 | 54.9 | 208 | -0.475 | -3.69 |
| Croatia | 2.38 | -2.77 | -65.2 | -55.4 | 4.45 | 14 |
| Cuba | -9.97 | 2.21 | 11.2 | 29.2 | 5.01 | 5.26 |
| Cyprus | 3.52 | -3.83 | 7.45 | -4.84 | -1.31 | -0.502 |
| CzechRepublic | -2.58 | 1.85 | -2.27 | 8.77 | 12.3 | 3.07 |
| Denmark | 1.45 | -1.47 | 6.75 | 4.58 | 0.531 | 7.28 |
| DominicanRepublic | 2.59 | 2.72 | -5.52 | -7.23 | 10.3 | 5.39 |
| Ecuador | 5.13 | 2.67 | -32.5 | -31.4 | 1.85 | 1.56 |
| ElSalvador | 4.71 | -0.495 | 26.8 | 12.6 | 8.22 | 4.38 |
| Estonia | 4.31 | -5.45 | 19.2 | -0.147 | 0.186 | 1.23 |
| Ethiopia | -10.6 | -3.01 | -23.4 | -23 | 1.95 | 7.59 |
| Finland | -2.05 | 5 | 6.41 | 14.1 | 14.4 | 7.5 |
| France | -12.6 | -13.3 | 28 | 23.1 | -2.97 | -18.3 |
| Germany | -7.69 | -4.65 | -5.2 | -0.402 | -0.297 | 3.5 |
| Ghana | 47.1 | 35.7 | -58.8 | -39.9 | -7.31 | 13.7 |
| Greece | -6.14 | -1.51 | 4.37 | 9.37 | -4 | 9.83 |
| Hungary | -12.8 | -14.1 | 145 | 54.5 | 6.25 | 48.9 |
| Iceland | -2.75 | 5.01 | 55.8 | 43.6 | 1.76 | 2.24 |
| India | 2.68 | -0.794 | -0.0667 | 0.299 | 5.87 | 1.84 |
| Indonesia | 9.53 | 10.9 | -12.6 | 0.469 | 3.93 | 4.27 |
| Iran | -12.2 | -1.77 | 18.2 | 40.9 | 4.42 | 2.03 |
| Iraq | -1.54 | 0.124 | 1280 | 220 | 0.0163 | 1.18 |
| Ireland | -8.19 | -4.05 | -14.3 | -8.38 | -1.83 | 9.61 |
| Israel | -2.57 | -2.86 | 17.4 | 14.6 | 11.2 | 1.71 |
| Italy | -5.05 | 0.128 | 0.804 | 6.79 | 4.64 | 7.65 |
| Jamaica | -7.29 | -3.05 | 26.5 | -25.2 | 14.1 | 5.11 |
| Japan | 2.06 | 5.34 | 27.5 | 11.5 | -7.99 | 1.91 |
| Kazakhstan | 2.14 | 1.06 | -43.5 | -36.2 | 1.14 | 7.05 |
| Kenya | -2.51 | -4.49 | 24 | 9.31 | 0.0898 | 3.43 |
| Kuwait | -3 | -7.44 | -51.5 | -43.3 | 10.6 | 3.69 |
| Latvia | -2.96 | -6.57 | 29.7 | 7.88 | 17.5 | 5.12 |
| Lithuania | -1.68 | -4.73 | 8.72 | -6.46 | -1.22 | 8.81 |
| Luxembourg | -2.47 | -1.47 | 2.31 | 13.5 | 1.9 | 5.21 |
| Madagascar | 1.74 | 15.6 | -55.9 | -37.2 | 2.21 | 6.21 |
| Malawi | -4.68 | 3.35 | -19.4 | 3.6 | -1.32 | 6.77 |
| Malaysia | -6.36 | 8.01 | -31.9 | -28.8 | 6.76 | 6.84 |
| Maldives | 12.5 | -5.48 | 191 | -2.97 | 8.62 | -0.978 |
| Malta | -0.808 | -2.83 | 0.0203 | -18 | 15.9 | 4.25 |
| Mexico | 4.45 | 12 | 3.91 | 17.3 | -2.08 | 6.14 |
| Morocco | -15.1 | 1.06 | 12 | 5.07 | 0.934 | 5.96 |
| Mozambique | 0.515 | 13.9 | 103 | 117 | 0.995 | 6.68 |
| Nepal | -6.76 | -0.677 | -0.023 | -1.49 | 5.02 | 11.9 |
| Netherlands | -2.32 | -2.01 | -3.84 | -1.02 | 7 | 8.4 |
| NewZealand | -7.31 | 24.3 | 269 | 73.5 | 7.28 | 13.6 |
| Nigeria | 4.01 | 0.931 | -1.94 | 33.3 | -0.171 | -1.5 |
| NorthMacedonia | -18.1 | -3.59 | 241 | 162 | 4.72 | 6.86 |
| Norway | -3.18 | -2.46 | 9.2 | 19.1 | -0.327 | 2.96 |
| Pakistan | -7.15 | -4.5 | -19 | -2.03 | 9.23 | 9.83 |
| Panama | 2.54 | 5.11 | -8.52 | -6.08 | 4.57 | 12 |
| Paraguay | -7.1 | -0.318 | 21.3 | 38 | -1.29 | 2.99 |
| Peru | 0.943 | 2.62 | -40.2 | -38.6 | 0.246 | 2.4 |
| Philippines | 13.4 | 2.05 | 70.1 | 54.9 | 4.43 | 6.87 |
| Poland | -13.9 | -12.8 | 212 | 138 | 2.88 | 32.8 |
| Portugal | -15 | -2.65 | 19.5 | 49.1 | -9.08 | 11.8 |
| Qatar | -8.86 | -7.39 | -13.9 | -18 | 1.52 | -2.18 |
| Romania | -2.37 | -0.667 | -13.2 | -1.71 | -3.38 | 6.69 |
| Russia | 0.622 | -0.501 | 16.1 | 15.4 | 14.3 | 1.72 |
| Rwanda | -11.1 | 0.649 | 144 | 111 | 9.8 | 2.37 |
| SaudiArabia | 2.87 | -2.52 | -26 | -8.98 | -5.29 | 5.63 |
| Senegal | -4.36 | -3.47 | -4.1 | -6.93 | 0.538 | 0.731 |
| Serbia | -8.06 | -4.08 | 8.66 | 18.4 | 1.51 | 6.16 |
| Singapore | 12 | 24.9 | -91.6 | -72.7 | 24.4 | 24.5 |
| Slovakia | -12.2 | 2.86 | 84.1 | 129 | 0.854 | 7.3 |
| Slovenia | -7.98 | -9.61 | 43.6 | 13.8 | 0.709 | 6.76 |
| SouthAfrica | -13.6 | -2.55 | -6.48 | 2.99 | -3.25 | 17.9 |
| SouthKorea | -20 | -5.71 | 9.12 | 27.1 | 4.94 | 9.99 |
| Spain | 11.4 | 1.46 | -38.8 | -47.9 | 18.8 | 9.43 |
| SriLanka | -11.5 | 0.297 | -10.1 | -29.5 | 2.93 | 5.82 |
| Sweden | 9.33 | -8.45 | -21.5 | -33 | -2.01 | 3.42 |
| Switzerland | 1.58 | -6.69 | 3.85 | -16.2 | 0.784 | 11.8 |
| Thailand | 10.6 | 9.39 | -1.08 | 71.8 | 49.9 | 38.5 |
| Togo | -11.3 | -9 | 463 | -28.9 | 6.87 | -0.477 |
| Tunisia | -12.2 | 6.43 | 58.7 | -2.13 | 0.903 | -0.345 |
| Turkey | -1.71 | -1.53 | -4.86 | -2.57 | 1.65 | 6.99 |
| UAE | 5.22 | 1.05 | 16.4 | 12.7 | 18.1 | -1.96 |
| UK | -10.6 | -4.67 | 10.4 | 24.9 | -11.3 | 23.7 |
| Ukraine | -0.855 | -0.811 | -1.31 | 11.8 | 1.05 | 1.14 |
| Uruguay | -0.885 | 0.233 | 2.91 | 7.16 | -0.93 | -0.215 |
| USA | 1.52 | -0.786 | -4.56 | -3.82 | 9.35 | 2.71 |
| **Average** | **-2.54** | **-0.301** | **39.1** | **15.1** | **3.69** | **6.87** |
| **Global Percentage Changes and Elasticities** |  |  |  |  |  |  |
|  | **Cases Early** | **Deaths Early** | **IFR Early** | **Cases Proj.** | **Deaths Proj.** | **IFR Proj.** |
| Global | -1.83 | -1.02 | 0.318 | -1.57 | 0.275 | 2.07 |

### Sensitivity to Parametric Assumptions

#### Setup

We conducted sensitivity analysis changing all the major pre-specified (i.e. not estimated) model parameters to assess 1) How sensitive key results are to those parametric assumptions. 2) How overall model fit to data changes with changing those general parameters. In this analysis we changed each parameter by +/- 5%, and calculated the elasticities of the 12 outcome measures discussed in the previous section with respect to each parameter. Those elasticities are calculated as fractional change in the outcome measure divided by fractional change in the input parameter. As such, they are dimensionless, with values below one indicating minor to modest sensitivities.

Table S7 reports the parameters over which we conducted the sensitivity analysis, their base values, and an overview of the results where averages over country-level outcomes and fit measures are reported (first row) along with aggregate outcomes (second row; for a total of 12 outcomes per parameter in two rows). We report full country-level sensitivity results for the reported outcomes online, in the GitHub repository for the research. All reported numbers are elasticity values.

#### Results

Overall elasticities remain modest (Table S7) but also include more notable cases where results for a specific country are rather sensitive to one parameter or the other. Note that the mechanisms generating these elasticities are complex, as they emerge from new calibration of the model and thus incorporate various compensatory mechanisms and feedback effects. Thus we do not attempt to explain detailed country-level elasticities (reported on GitHub). With that caveat in mind, we provide an overview of the results here and tables on GitHub provide more in-depth outcomes.

The average residence time in hospitals has an impact on death rates and IFR, as increasing the residence time reduces available beds and exacerbates hospital shortages but also increased disease period in hosptial allows for more infection inside hospitals (note that in the sensitivity analysis the change in hospital residence time is not coupled with changing the disease duration outside of hospital). Increasing incubation period may modestly increase death under-count ratio and projected death rates, but slightly reduce projected infection rates; results for infection and death numbers are sensitive for a few countries due to shifts in overall parameters as a result of changing incubation period. Impact of Onset-to-detection delay is somewhat similar. Post-detection resolution time slightly increases cases and deaths as people spend more time in infectious states. Relative risk of transmission by presymptomatics would increase undercounts, infections and deaths, with a stronger impact on case under-count (thus reducing IFR). Increasing this parameter would also slightly increase the gap between the model and case data (MAEN). Finally, increasing the sensitivity of COVID test would reduce undercounts in cases and deaths and thus bring down total cases and deaths by stronger behavioral responses.

**Table S7.** Overview of parametric sensitivity results, reported as **elasticities (fractional change in outcome divided by fractional change in input parameter)**. Each parameter is followed by its base value, and two rows of outcomes identified on the top. First row includes averages of elasticities for 4 outcomes and 2 fit measures across 92 countries. Second row includes elasticity measures calculated over all simulated populations on 22 December 2020 and 30 June 2021.

| *Parameter* | **Ave Case Undercount Ratio** | **Ave Death Undercount Ratio** | **Ave 6/30/21 Infection Rate** | **Ave 6/30/21 Death Rate** | **Ave MAEN Infection Rate** | **Ave MAEN Death Rate** |
| --- | --- | --- | --- | --- | --- | --- |
|  | **Dec 22 Cml. Cases** | **Dec 22 Cml. Deaths** | **Historical IFR** | **6/30/21 Cml. Cases** | **6/30/21 Cml. Deaths** | **6/30/21 Cml. IFR** |
| *Hospitalized Resolution Time (20 days)* | -0.00889 | 0.0105 | 0.355 | 0.501 | -0.00379 | 0.0116 |
|  | 0.0173 | -0.0227 | -0.0456 | -0.0574 | -0.0684 | -0.0163 |
| *Incubation Period (5 days)* | 0.142 | 0.00899 | 0.0789 | 0.192 | 0.0117 | -0.0163 |
|  | 0.231 | 0.105 | -0.104 | 0.207 | 0.141 | -0.0688 |
| *Onset to Detection Delay (5 days)* | 0.12 | 0.0298 | 0.135 | 0.219 | 0.00522 | -0.0257 |
|  | 0.169 | 0.0254 | -0.117 | 0.182 | 0.0836 | -0.1 |
| *Post-Detection Phase Resolution Time (10 days)* | 0.000354 | -0.0037 | 0.00163 | 0.0044 | -0.0002 | 0.00281 |
|  | -0.0069 | -0.00299 | 0.00388 | -0.00112 | 0.00556 | 0.00741 |
| *Relative Risk of Transmission by Presymptomatic (1)* | 0.259 | -0.00558 | -0.398 | -0.558 | 0.0125 | 0.0334 |
|  | 0.19 | 0.0162 | -0.189 | 0.12 | -0.043 | -0.175 |
| *Sensitivity of COVID Test (0.7)* | -0.421 | -0.509 | -0.183 | -0.398 | 0.0395 | -0.0364 |
|  | -0.287 | -0.51 | -0.241 | -0.316 | -0.488 | -0.159 |

### Country level outcome elasticities for model parameters

See the online Github repository at https://github.com/tseyanglim/CovidGlobal for country-level outcome tables.

#### Sensitivity of results to exclusion of major countries

We repeated the analysis in three additional setups, excluding the top five countries by population (India, USA, Indonesia, Pakistan, Nigeria), by true infections to date (USA, Mexico, Iran, Peru, Indonesia), and by reported infections to date (USA, Russia, India, UK, Spain) from the estimation and analysis. These analyses inform the sensitivity of overall findings to data from specific countries.

In each case we report the *percentage* of change in the country level and aggregate outcome measures (those defined and discussed above). Table S*8* summarizes the results; full country-level outcome tables are available on the online Github repository at https://github.com/tseyanglim/CovidGlobal.

##### Results

Overall, the impact of excluding the top countries from analysis on historical fit and outcome measures is modest: fit quality does not change by more than 1.2% in 95% of country-outcome combinations. Moreover, the historical under-reporting ratios remain largely unchanged (changing by no more than 1.3% overall for 95% of country-outcome combinations). Cases and deaths summed up over the sample naturally change when excluding the larger countries or those with more infections (the bottom rows for each analysis). Long-term country level projections (items 3 and 4 in odd rows) change little (less than 2%) for most countries, however, a few show notable variations, suggesting high sensitivity to their response functions such that minor changes in parameters (which change due to the coupling with other nations) could significantly alter future projected outcomes. Those countries with significant sensitivity include: Australia, Cuba, El Salvador, Iceland, Kuwait, New Zealand, and Togo.

**Table S8.** Sensitivity of outcomes (in percentage change from baseline) to exclusion of top five countries by true infection, population, and reported infection. Each analysis is specified by the removed countries and followed by two rows of outcomes identified on the top. First row includes averages of 4 outcomes and 2 fit measures across 92 countries. Second row includes measures calculated over all simulated populations on 22 December 2020 and 30 June 2021.

| **Removed Countries** | **Ave Case Undercount Ratio** | **Ave Death Undercount Ratio** | **Ave 6/30/21 Infection Rate** | **Ave 6/30/21 Death Rate** | **Ave MAEN Infection Rate** | **Ave MAEN Death Rate** |
| --- | --- | --- | --- | --- | --- | --- |
|  | **Dec 22 Cml. Cases** | **Dec 22 Cml. Deaths** | **Historical IFR** | **6/30/21 Cml. Cases** | **6/30/21 Cml. Deaths** | **6/30/21 Cml. IFR** |
| *Top True Infections: USA, Mexico, Iran, Peru, Indonesia* | -0.011 | 0.0164 | 0.0743 | -0.117 | -0.0969 | 0.0701 |
|  | -33.5 | -38.2 | -5.94 | -32.4 | -35.2 | -4.43 |
| *Top Populations: India, USA, Indonesia, Pakistan, Nigeria* | -0.0178 | 0.00581 | 0.0606 | 0.0535 | -0.0374 | 0.0373 |
|  | -22.8 | -27.9 | -6.95 | -24.8 | -25.9 | -1.96 |
| *Top Reported Infections: USA, Russia, India, UK, Spain* | 0.183 | 0.0948 | 0.19 | 0.0557 | 0.0315 | -0.176 |
|  | -28.4 | -34.9 | -9.4 | -27.5 | -32 | -5.91 |

## S8 COMPLETE MODEL DOCUMENTATION

Below we provide complete model equations and units. The model, in the .mdl format, which can be opened using the Vensim simulation software, or the free Vensim model reader) is available with this appendix as well, and online at https://github.com/tseyanglim/CovidGlobal. Most of the equations are self-explanatory. The “[…]” notation is used to subscript variables over a set of members. For example, the subscript “Rgn” is used to identify different countries. Therefore [Rgn] indicates that a variable is defined separately for each member of the set “Rgn”. Other subscript ranges used in the equations are:

expnt: Used for numerically solving the probability of missing symptoms equation. pdim: Used for setting policy levels for a few variables.

Priors: Used for implementing the random effects estimation components. Each estimated parameter is mapped into an element of this subscript to simplify vector-based calculations.

Series: The data series (Infection, Death, Recovery).

TstSts: The test status including those confirmed (‘Tested’) and those unconfirmed (‘Notest’).

Variables units are provided for most equations. Those missing units are ones subscripted over different variable types or using equations that (utilizing power or log functions) cannot have consistent units.

### Complete equations and units

1. a[Rgn] = XIDZ (Potential Test Demand from Susceptible Population[Rgn], Positive Candidates Interested in Testing Poisson Subset Adj[Rgn], 1) Units: dmnl
2. AbsPrcErr[Rgn,Series] = if then else (DataIncluded[Rgn] = 0, :NA:, ZIDZ (abs (FlowResiduals[Rgn,Series]), DataFlowOverTime[Rgn,Series])) Units: dmnl
3. AbsStd[Priors] = 0.2, 0.3, 0.1, 0.2, 0.0002, 0.2, 10, 0.03, 6, 0.1, 0.1, 0.1, 0.8, 0.1, 1e-05, 10, 0.01, 0.005, 0.01, 10, 10, 10, 0.01, 0.5, 0.5 Units: **undefined**
4. Active Test Rate[Rgn] = if then else (Time < New Testing Time, DataTestRate[Rgn], External Test Rate[Rgn]) Units: Person/Day
5. ActiveAve[PriorEndoAve] = INITIAL(InputAve[PriorEndoAve] * (1 - SW EndoAve) + SW EndoAve * CalcAve[PriorEndoAve])
6. ActiveAve[PMT] = InputAve[PMT] Units: **undefined**
7. Activities Allowed by Government[Rgn] = 1 Units: dmnl [0,1,0.01]
8. Additional Asymptomatic Fraction Init[Rgn] = Additional Asymptomatic Relative to Symptomatic Init[Rgn] / (1 + Additional Asymptomatic Relative to Symptomatic Init[Rgn]) Units: dmnl
9. Additional Asymptomatic Post Detection[Rgn] = Weighted Infected Post Detection Gate[Rgn] * Additional Asymptomatic Relative to Symptomatic[Rgn] / (1 + Additional Asymptomatic Relative to Symptomatic[Rgn]) Units: Person
10. Additional Asymptomatic Relative to Symptomatic[Rgn] = ZIDZ (Total Asymptomatic Fraction Net[Rgn] - exp (− Covid Acuity[Rgn]), 1 - Total Asymptomatic Fraction Net[Rgn]) Units: dmnl
11. Additional Asymptomatic Relative to Symptomatic Init[Rgn] = INITIAL(ZIDZ (Total Asymptomatic Fraction Init Net[Rgn] - exp (− Covid Acuity Relative to Flu Init Net[Rgn] * Flu Acuity), 1 - Total Asymptomatic Fraction Init Net[Rgn])) Units: dmnl
12. Adherence Fatigue Time = 100 Units: Day
13. AdvCntrs[Rgn] = 1 Units: dmnl
14. All Recovery[Rgn] = Recovery of Confirmed[Rgn] + Recovery of Untested[Rgn] + sum (Hospital Discharges[Rgn,TstSts!]) Units: Person/Day
15. Allocated Fraction COVID Hospitalized[Rgn] = min (1, (− Expected Positive Poisson Covid Patients[Rgn] + Sqrt (Expected Positive Poisson Covid Patients[Rgn] ^ 2 + 4 * Effective Hospital Capacity[Rgn] * Effective Hospital Capacity[Rgn])) / (2 * Effective Hospital Capacity[Rgn])) Units: dmnl
16. Allocated Fration NonCOVID Hospitalized[Rgn] = SMOOTHI (Allocated Fraction COVID Hospitalized[Rgn] ^ 2, Hospital Adj T, 1) Units: dmnl
17. alp[Rgn,Infection] = min (maxAlp, ialp * alpR[Rgn])
18. alp[Rgn,Death] = min (1, dalp * alpR[Rgn])
19. alp[Rgn,Test] = min (1, talp * alpR[Rgn]) Units: dmnl
20. alpR[Rgn] = 1 Units: dmnl
21. AntiVaxxerFrac[Rgn] = 0 Units: dmnl
22. Area of Region[Rgn] = GET VDF CONSTANTS(Constant Data File, ‘DataConstants[Rgn]’, 5) Units: Km*Km
23. Average Acuity Hospitalized[Rgn,Tested] = Average Acuity of Positively Tested[Rgn] * XIDZ ((1 - Probability of Missing Acuity Signal at Hospitals[Rgn,Tested] * Fraction Poisson not Hospitalized[Rgn,Tested] ^ 2), 1 - Fraction Poisson not Hospitalized[Rgn,Tested], 2 * Probability of Missing Acuity Signal at Hospitals[Rgn,Tested])
24. Average Acuity Hospitalized[Rgn,Notest] = ZIDZ (Average Acuity of Untested Poisson Subset[Rgn] * (1 - Probability of Missing Acuity Signal at Hospitals[Rgn,Notest] * Fraction Poisson not Hospitalized[Rgn,Notest] ^ 2), 1 - Fraction Poisson not Hospitalized[Rgn,Notest]) Units: dmnl
25. Average Acuity in Susceptible[Rgn] = ZIDZ (Sympthoms in Susceptible[Rgn], Susceptible[Rgn]) Units: dmnl
26. Average Acuity Not Hospitalized[Rgn,Notest] = ZIDZ (Average Acuity Not Hospitalized Poisson[Rgn,Notest] * Infectious not Tested or in Hospitals Poisson[Rgn], “Infected Unconfirmed Post-Detection”[Rgn])
27. Average Acuity Not Hospitalized[Rgn,Tested] = Average Acuity Not Hospitalized Poisson[Rgn,Tested] Units: dmnl
28. Average Acuity Not Hospitalized Poisson[Rgn,Tested] = Max (0, Probability of Missing Acuity Signal at Hospitals[Rgn,Tested] * Average Acuity of Positively Tested[Rgn] * Fraction Poisson not Hospitalized[Rgn,Tested])
29. Average Acuity Not Hospitalized Poisson[Rgn,Notest] = Max (0, Probability of Missing Acuity Signal at Hospitals[Rgn,Notest] * Average Acuity of Untested Poisson Subset[Rgn] * Fraction Poisson not Hospitalized[Rgn,Notest]) Units: dmnl
30. Average Acuity of Positively Tested[Rgn] = Covid Acuity[Rgn] * XIDZ ((1 - Prob Missing Symptom[Rgn] * Fraction Interested not Tested[Rgn] ^ 2), 1 - Fraction Interested not Tested[Rgn], 2 * Prob Missing Symptom[Rgn]) Units: dmnl
31. Average Acuity of Untested Poisson Subset[Rgn] = ZIDZ (Poisson Subset Reaching Test Gate[Rgn] * Covid Acuity[Rgn] - Positive Tests of Infected[Rgn] * Average Acuity of Positively Tested[Rgn], Poisson Subset Not Tested Passing Gate[Rgn]) Units: dmnl
32. b[Rgn] = ZIDZ (Testing on Living[Rgn] - Positive Candidates Interested in Testing Poisson Subset Adj[Rgn] - Potential Test Demand from Susceptible Population[Rgn], Positive Candidates Interested in Testing Poisson Subset Adj[Rgn]) Units: dmnl
33. Base Fatality Rate for Unit Acuity[Rgn] = 0.0006 Units: dmnl
34. Base Fatality Rate for Unit Acuity Net[Rgn] = INITIAL(Base Fatality Rate for Unit Acuity[Rgn] * (1 - SW Gen[BsFtRt]) + SW Gen[BsFtRt] * InputAve[BsFtRt]) Units: dmnl [0,0.01]
35. BaseError = 5 Units: Person
36. Baseline Cumulative Cases for Learning = 0.005 Units: dmnl
37. Baseline Daily Fraction Susceptible Seeking Tests[Rgn] = 0.001 Units: 1/Day
38. Baseline Fatality Multiplier[Rgn] = INITIAL(Demographic Impact on Fatality Relative to China[Rgn] * Base Fatality Rate for Unit Acuity Net[Rgn] * Liver Disease Impact on Fatality[Rgn] * Obesity Impact on Fatality[Rgn] * Chronic Impact on Fatality[Rgn]) Units: dmnl [0,0.1]
39. Baseline Risk of Transmission by Asymptomatic[Rgn] = INITIAL(Baseline Transmission Multiplier for Untested Symptomatic * Multiplier Transmission Risk for Asymptomatic Net[Rgn]) Units: dmnl
40. Baseline Transmission Multiplier for Untested Symptomatic = 1 Units: dmnl
41. Bed per Square Kilometer[Rgn] = INITIAL(Nominal Hospital Capacity[Rgn] / Area of Region[Rgn]) Units: Person/(Km*Km)
42. Beds per Thousand Population[Rgn] = GET VDF CONSTANTS(Constant Data File, ‘DataConstants[Rgn]’, 11) Units: dmnl
43. CalcAve[Priors] = INITIAL(sum (RegionalInputs[Priors,Rgn!]) / ELMCOUNT(Rgn)) Units:**undefined**
44. cft[Rgn,p2] = lnymix[Rgn,p2]
45. cft[Rgn,p3] = lnymix[Rgn,p3] - lnymix[Rgn,p2]
46. cft[Rgn,p4] = (ln (min (100, Max (1e-06, ZIDZ (lnymix[Rgn,p4] - lnymix[Rgn,p2], lnymix[Rgn,p3] - lnymix[Rgn,p2]) / ln (2))))) Units: dmnl
47. Chng Cml Dth Untst Untrt[Rgn] = Deaths of Symptomatic Untested[Rgn] - Post Mortem Test Rate[Rgn] * Frac Post Mortem from Untreated[Rgn] Units: Person/Day
48. Chronic Death Rate[Rgn] = GET VDF CONSTANTS(Constant Data File, ‘DataConstants[Rgn]’, 17) Units: dmnl
49. Chronic Impact on Fatality[Rgn] = INITIAL((Chronic Death Rate[Rgn] / MeanChronic) ^ Sens Chronic Impact Net[Rgn]) Units: dmnl
50. Cml Death Frac In Hosp[Rgn] = XIDZ (Cumulative Deaths at Hospital[Rgn,Tested] + Cumulative Deaths at Hospital[Rgn,Notest], Cumulative Deaths[Rgn], 1) Units: dmnl
51. Cml Death fraction in hospitals large enough = sum (if then else (Cml Death Frac In Hosp[Rgn!] < MinHspDTresh, 1, 0)) Units: dmnl
52. Cml Death Hsp Inc[Rgn,Tested] = Hospitalized Infectious Deaths[Rgn,Tested] + PostMortemCorrection[Rgn]
53. Cml Death Hsp Inc[Rgn,Notest] = Hospitalized Infectious Deaths[Rgn,Notest] - PostMortemCorrection[Rgn] Units: Person/Day
54. Cml Known Death Frac Hosp[Rgn] = XIDZ (Cumulative Deaths at Hospital[Rgn,Tested], Cumulative Deaths[Rgn], 1) Units: dmnl
55. CmltErrPW = 2 Units: dmnl
56. CmltPenaltyScl = 0 Units: dmnl
57. CmltToInclude[Series] = 0, 0, 0 Units: dmnl
58. Confirmation Impact on Contact[Rgn] = 0.002 Units: dmnl
59. Confirmed Recovered[Rgn] = INTEG(Recovery of Confirmed[Rgn], 0) Units: Person
60. Constant Data File :IS: ‘CovidModelInputs - ConstantData.vdf’
61. Contacts Relative to Normal[Rgn] = min (Voluntary Reduction in Contacts[Rgn], Activities Allowed by Government[Rgn]) Units: dmnl
62. Continue without Testing[Rgn] = Reaching Testing Gate[Rgn] - Symptomatic Infected to Testing[Rgn] - Untested symptomatic Infected to Hospital[Rgn] Units: Person/Day
63. Count Missed Death[Rgn] = if then else (Excess Death Start Count[Rgn] = :NA:, 0, if then else (Time >= Excess Death Start Count[Rgn] :AND: Time <= Excess Death End Count[Rgn], Cml Death Hsp Inc[Rgn,Notest] + Chng Cml Dth Untst Untrt[Rgn], 0)) Units: Person/Day
64. Covid Acuity[Rgn] = Flu Acuity * Covid Acuity Relative to Flu Net[Rgn] Units: dmnl
65. Covid Acuity Relative to Flu[Rgn] = 6 Units: dmnl
66. Covid Acuity Relative to Flu Init Net[Rgn] = INITIAL(Covid Acuity Relative to Flu[Rgn] * (1 - SW Gen[Acty]) + SW Gen[Acty] * InputAve[Acty]) Units: dmnl
67. Covid Acuity Relative to Flu Net[Rgn] = Average Acuity in Susceptible[Rgn] / (1 - Additional Asymptomatic Fraction Init[Rgn]) Units: dmnl
68. Covid Poisson Fraction in Hospital[Rgn] = ZIDZ (Total Covid Hospitalized[Rgn], Infectious not Tested or in Hospitals Poisson[Rgn] + Infectious Confirmed Not Hospitalized[Rgn] + Total Covid Hospitalized[Rgn]) Units: dmnl
69. CRW[Rgn] Units: dmnl
70. Cumulative Cases[Rgn] = INTEG(New Cases[Rgn], 0) Units: Person
71. Cumulative Confirmed Cases[Rgn] = INTEG(SimFlowOverTime[Rgn,Infection], 0) Units: Person
72. Cumulative Confirmed Recovered[Rgn] = Confirmed Recovered[Rgn] + Cumulative Recovered at Hospitals[Rgn,Tested] Units: Person
73. Cumulative Death Fraction[Rgn] = ZIDZ (Cumulative Deaths[Rgn], Cumulative Deaths[Rgn] + Cumulative Recoveries[Rgn]) Units: dmnl
74. Cumulative Deaths[Rgn] = INTEG(Death Rate[Rgn], 0) Units: Person
75. Cumulative Deaths at Hospital[Rgn,TstSts] = INTEG(Cml Death Hsp Inc[Rgn,TstSts], 0) Units: Person
76. Cumulative Deaths of Confirmed[Rgn] = INTEG(SimFlowOverTime[Rgn,Death], 0) Units: Person
77. Cumulative Deaths of Confirmed Untreated[Rgn] = INTEG(Deaths of Confirmed[Rgn] + Post Mortem Test Untreated[Rgn], 0) Units: Person
78. Cumulative Deaths Untested Untreated[Rgn] = INTEG(Chng Cml Dth Untst Untrt[Rgn], 0) Units: Person
79. Cumulative Fraction Total Cases Hospitalized[Rgn] = ZIDZ (sum (Cumulative Deaths at Hospital[Rgn,TstSts!] + Cumulative Recovered at Hospitals[Rgn,TstSts!] + Hospitalized Infectious[Rgn,TstSts!]), Cumulative Cases[Rgn]) Units: dmnl
80. Cumulative Missed Death[Rgn] = INTEG(Count Missed Death[Rgn], 0) Units: Person
81. Cumulative Negative Tests[Rgn] = INTEG(Negative Test Results[Rgn], 0) Units: Person
82. Cumulative Recovered at Hospitals[Rgn,TstSts] = INTEG(Hospital Discharges[Rgn,TstSts], 0) Units: Person
83. Cumulative Recoveries[Rgn] = INTEG(All Recovery[Rgn], 0) Units: Person
84. Cumulative Tests Conducted[Rgn] = INTEG(SimTestRate[Rgn], 0) Units: Person
85. Cumulative Tests Data[Rgn] = INTEG(TstInc[Rgn], 0) Units: Person
86. Current Test Rate per Capita[Rgn] = INITIAL(if then else (DataLastTestRate[Rgn] = :NA:, 0, DataLastTestRate[Rgn] / Population[Rgn])) Units: 1/Day
87. dalp = 0.1 Units: dmnl
88. Data Excess Deaths[Rgn] = GET VDF CONSTANTS(Constant Data File, ‘DataConstants[Rgn]’, 15) Units: Person
89. DataAttentionTime[Rgn] = GET VDF CONSTANTS(Constant Data File, ‘DataConstants[Rgn]’, 9) Units: Day
90. DataCmltDeath[Rgn] Units: Person
91. DataCmltInfection[Rgn] Units: Person
92. DataCmltOverTime[Rgn,Infection] :RAW: := DataCmltInfection[Rgn]
93. DataCmltOverTime[Rgn,Death] :RAW: := DataCmltDeath[Rgn]
94. DataCmltOverTime[Rgn,Test] = DataCmltTest[Rgn] Units: Person
95. DataCmltTest[Rgn] Units: Person
96. DataFlowDeath[Rgn] Units: Person/Day
97. DataFlowInfection[Rgn] Units: Person/Day
98. DataFlowOverTime[Rgn,Infection] :RAW: := DataFlowInfection[Rgn]
99. DataFlowOverTime[Rgn,Death] :RAW: := DataFlowDeath[Rgn]
100. DataFlowOverTime[Rgn,Test] :RAW: := DataTestRate[Rgn] Units: Person/Day
101. DataFlowRecovery[Rgn] :RAW: Units: Person/Day
102. DataIncluded[Rgn] = if then else (Max (Cumulative Deaths[Rgn], Max (Cumulative Confirmed Cases[Rgn], DataCmltOverTime[Rgn,Infection])) > ThrsInc[Rgn], 1, 0) * DataLimitFromTime[Rgn] Units: dmnl
103. DataLastTestRate[Rgn] = INITIAL(GET DATA AT TIME (DataTestRate[Rgn], min (LastTestDate[Rgn], New Testing Time))) Units: Person/Day
104. DataLimitFromTime[Rgn] = if then else (Time > StopDataUseTime[Rgn], 0, 1) Units: dmnl
105. DataTestCapacity[Rgn] Units: Person/Day
106. DataTestRate[Rgn] Units: Person/Day
107. Day of First Case Report in JHU Database = 99 Units: Day
108. Days per Year = 365 Units: Day/Year
109. Death Rate[Rgn] = Deaths of Confirmed[Rgn] + Deaths of Symptomatic Untested[Rgn] + sum (Hospitalized Infectious Deaths[Rgn,TstSts!]) Units: Person/Day
110. DeathFractionCounted[Rgn] = if then else (DataCmltOverTime[Rgn,Death] = :NA:, 0, ZIDZ (DataCmltOverTime[Rgn,Death], Cumulative Deaths[Rgn])) Units: dmnl
111. Deaths of Confirmed[Rgn] = Tested Untreated Resolution[Rgn] * Fatality Rate Untreated[Rgn,Tested] Units: Person/Day
112. Deaths of Symptomatic Untested[Rgn] = Infectious not Tested or in Hospitals Poisson[Rgn] / “Post-Detection Phase Resolution Time” * Fatality Rate Untreated[Rgn,Notest] Units: Person/Day
113. Delay Order = 1 Units: dmnl
114. Demographic Impact on Fatality Relative to China[Rgn] = GET VDF CONSTANTS(Constant Data File, ‘DataConstants[Rgn]’, 12) Units: dmnl
115. Di[Rgn,Series] = DataFlowOverTime[Rgn,Series] Units: Person/Day
116. Different Infectious Counted[Rgn] = “Pre-Symptomatic Infected”[Rgn] + Infected pre Detection[Rgn] + (Additional Asymptomatic Post Detection[Rgn] + “Poisson Not-tested Asymptomatic”[Rgn]) + (Infectious not Tested or in Hospitals Poisson[Rgn] - “Poisson Not-tested Asymptomatic”[Rgn]) + Infectious Confirmed Not Hospitalized[Rgn] + sum (Hospitalized Infectious[Rgn,TstSts!]) Units: Person
117. Discount Rate Annual = 0.03 Units: 1/Year [1e-05,0.2]
118. Discount Rate per Day = INITIAL(Discount Rate Annual / Days per Year) Units: 1/Day
119. Dread Factor in Risk Perception[Rgn] = 25 Units: dmnl [0,10000]
120. Dread Factor in Risk Perception Net[Rgn] = if then else (Response Policy Time On < Time, (1 + Response Policy Weight) * Dread Factor in Risk Perception[Rgn], Dread Factor in Risk Perception[Rgn]) * Impact of Adherence Fatigue[Rgn] Units: dmnl
121. Dread Factor Policy = 3000 Units: dmnl
122. Effective Hospital Capacity[Rgn] = Nominal Hospital Capacity[Rgn] * Normalized Hospital density[Rgn] ^ Impact of Population Density on Hospital Availability[Rgn] Units: Person
123. eps = 0.001 Units: Person/Day
124. Excess Death End Count[Rgn] = GET VDF CONSTANTS(Constant Data File, ‘DataConstants[Rgn]’, 14) Units: Day
125. Excess Death Mean Frac = 0.9 Units: dmnl
126. Excess Death Range Frac = 0.2 Units: dmnl
127. Excess Death Rate Error[Rgn] = if then else (Data Excess Deaths[Rgn] < 50, 0, ZIDZ (Cumulative Missed Death[Rgn] - Excess Death Mean Frac * Data Excess Deaths[Rgn], Excess Death Range Frac * Data Excess Deaths[Rgn]) ^ 4) Units: dmnl
128. Excess Death Start Count[Rgn] = GET VDF CONSTANTS(Constant Data File, ‘DataConstants[Rgn]’, 13) Units: Day
129. Expected Positive Poisson Covid Patients[Rgn] = sum (Potential Hospital Demand[Rgn,TstSts!]) * “Post- Detection Phase Resolution Time” Units: Person
130. expnt : (p2-p4)
131. External Test Rate[Rgn] = Population[Rgn] * Policy Test Rate[Rgn] Units: Person/Day
132. Extrapolated Estimator[Rgn] = if then else (Covid Acuity Relative to Flu Net[Rgn] > 1, cft[Rgn,p2] + cft[Rgn,p3] * (Covid Acuity Relative to Flu Net[Rgn] - 1) ^ cft[Rgn,p4], lnymix[Rgn,p2]) Units: dmnl
133. Fatality Rate Treated[Rgn,TstSts] = min (1, Baseline Fatality Multiplier[Rgn] * TimeVar Impact of Treatment on Fatality[Rgn] * Average Acuity Hospitalized[Rgn,TstSts] ^ Sensitivity of Fatality Rate to Acuity Net[Rgn]) Units: dmnl
134. Fatality Rate Untreated[Rgn,TstSts] = min (1, Baseline Fatality Multiplier[Rgn] * Average Acuity Not Hospitalized Poisson[Rgn,TstSts] ^ Sensitivity of Fatality Rate to Acuity Net[Rgn] * Time variant change in fatality[Rgn]) Units: dmnl
135. Final Test Rate Per Capita[Rgn] = INITIAL(Current Test Rate per Capita[Rgn] + Weight Max in Test Goal * Max (0, (Max Test Rate per Capita - Current Test Rate per Capita[Rgn]))) Units: 1/Day
136. FINAL TIME = 444 Units: Day [50,182,1]
137. FlowResiduals[Rgn,Series] = if then else (DataFlowOverTime[Rgn,Series] = :NA:, :NA:, DataFlowOverTime[Rgn,Series] - MeanFlowOverTime[Rgn,Series]) Units: Person/Day
138. FlowToInclude[Series] = 1, 1, 0 Units: dmnl
139. Flu Acuity = 1 Units: dmnl
140. Flu Acuity Relative to Covid[Rgn] = Flu Acuity / Covid Acuity[Rgn] Units: dmnl
141. Frac Post Mortem from Untreated[Rgn] = SMOOTHI (ZIDZ (Deaths of Symptomatic Untested[Rgn], Deaths of Symptomatic Untested[Rgn] + Hospitalized Infectious Deaths[Rgn,Notest]), Post Mortem Test Delay, 1) Units: dmnl
142. frac rampup = 0.5 Units: dmnl
143. FracNotVaccinated Susceptible[Rgn] = Susceptible[Rgn] / (Initial Population[Rgn] - Vaccinated[Rgn]) Units: dmnl
144. FracThresh = 0.001 Units: dmnl
145. Fraction Covid Death In Hospitals Previously Tested[Rgn] = ZIDZ (Hospitalized Infectious Deaths[Rgn,Tested], sum (Hospitalized Infectious Deaths[Rgn,TstSts!])) Units: dmnl
146. Fraction Covid Hospitalized Positively Tested[Rgn] = ZIDZ (Hospitalized Infectious[Rgn,Tested], Total Covid Hospitalized[Rgn]) Units: dmnl
147. Fraction Infected[Rgn] = Cumulative Cases[Rgn] / Initial Population[Rgn] Units: dmnl
148. Fraction Interested not Tested[Rgn] = 1 - ZIDZ (Total Test on Covid Patients[Rgn], Positive Candidates Interested in Testing Poisson Subset[Rgn]) Units: dmnl
149. Fraction Interseted not Correctly Tested[Rgn] = 1 - (1 - Fraction Interested not Tested[Rgn]) * Sensitivity of COVID Test Units: dmnl
150. Fraction of Additional Symptomatic[Rgn] = Additional Asymptomatic Relative to Symptomatic[Rgn] / (1 + Additional Asymptomatic Relative to Symptomatic[Rgn]) Units: dmnl
151. Fraction of Fatalities Screened Post Mortem[Rgn] = Indicated Fraction Post Mortem Testing[Rgn] * Switch for Government Response[Rgn] Units: dmnl
152. Fraction of Population Hospitalized for Covid[Rgn] = Total Covid Hospitalized[Rgn] / Population[Rgn] Units: dmnl
153. Fraction Poisson not Hospitalized[Rgn,Tested] = exp (− Average Acuity of Positively Tested[Rgn] * (1 - Probability of Missing Acuity Signal at Hospitals[Rgn,Tested]))
154. Fraction Poisson not Hospitalized[Rgn,Notest] = exp (− Average Acuity of Untested Poisson Subset[Rgn] * (1 - Probability of Missing Acuity Signal at Hospitals[Rgn,Notest])) Units: dmnl
155. Fraction Seeking Test[Rgn] = 1 Units: dmnl
156. Fraction Tests Positive[Rgn] = ZIDZ (Positive Tests of Infected[Rgn], Testing on Living[Rgn]) Units: dmnl
157. Fraction Tests Positive Data[Rgn] = min (1, ZIDZ (DataFlowInfection[Rgn], Active Test Rate[Rgn])) Units: dmnl
158. Global Cases = sum (Cumulative Cases[Rgn!]) Units: Person
159. Global Deaths = sum (Cumulative Deaths[Rgn!]) Units: Person
160. Global IFR = ZIDZ (Global Deaths, Global Cases - sum (Different Infectious Counted[Rgn!])) Units: dmnl
161. Government Response Start Time[Rgn] = INITIAL(DataAttentionTime[Rgn] + Day of First Case Report in JHU Database) Units: Day
162. Herd Immunity Fraction = 0.6 Units: dmnl
163. Hospital Adj T = 1 Units: Day
164. Hospital Admission Infectious[Rgn,TstSts] = Hospital Admits All[Rgn,TstSts] Units: Person/Day
165. Hospital Admit Ratio[Rgn,TstSts] = XIDZ (Hospital Admits All[Rgn,TstSts], Potential Hospital Demand[Rgn,TstSts], 1) Units: dmnl
166. Hospital Admits All[Rgn,Tested] = Hospital Demand from Tested[Rgn] * Allocated Fraction COVID Hospitalized[Rgn]
167. Hospital Admits All[Rgn,Notest] = Hospital Demand from Not Tested[Rgn] * Allocated Fraction COVID Hospitalized[Rgn] Units: Person/Day
168. Hospital Demand from Not Tested[Rgn] = Poisson Subset Not Tested Passing Gate[Rgn] * (1 - exp (− Average Acuity of Untested Poisson Subset[Rgn] * (1 - PMAS Unconfirmed for Hospital Demand[Rgn]))) Units: Person/Day
169. Hospital Demand from Tested[Rgn] = Positive Tests of Infected[Rgn] * (1 - exp (− Average Acuity of Positively Tested[Rgn] * (1 - PMAS Confirmed for Hospital Demand[Rgn]))) Units: Person/Day
170. Hospital Discharges[Rgn,TstSts] = (1 - Fatality Rate Treated[Rgn,TstSts]) * Hospital Outflow Covid Positive[Rgn,TstSts] Units: Person/Day
171. Hospital Outflow Covid Positive[Rgn,TstSts] = Hospitalized Infectious[Rgn,TstSts] / Hospitalized Resolution Time Units: Person/Day
172. Hospitalized CFR Cumulative[Rgn,TstSts] = ZIDZ (Cumulative Deaths at Hospital[Rgn,TstSts], Cumulative Deaths at Hospital[Rgn,TstSts] + Cumulative Recovered at Hospitals[Rgn,TstSts]) Units: dmnl
173. Hospitalized Infectious[Rgn,TstSts] = INTEG(Hospital Admission Infectious[Rgn,TstSts] - Hospitalized Infectious Deaths[Rgn,TstSts] - Hospital Discharges[Rgn,TstSts], 0) Units: Person
174. Hospitalized Infectious Deaths[Rgn,TstSts] = Fatality Rate Treated[Rgn,TstSts] * Hospital Outflow Covid Positive[Rgn,TstSts] Units: Person/Day
175. Hospitalized Resolution Time = 20 Units: Day
176. Hospitalized True CFR[Rgn] = ZIDZ (sum (Hospitalized Infectious Deaths[Rgn,TstSts!]), sum (Hospital Outflow Covid Positive[Rgn,TstSts!])) Units: dmnl
177. Hospitalized True CFR Cumulative[Rgn] = ZIDZ (sum (Cumulative Deaths at Hospital[Rgn,TstSts!]), sum (Cumulative Deaths at Hospital[Rgn,TstSts!] + Cumulative Recovered at Hospitals[Rgn,TstSts!])) Units: dmnl
178. ialp = 0.1 Units: dmnl
179. Impact of Adherence Fatigue[Rgn] = Recent Relative Contacts[Rgn] ^ (Strength of Adherence Fatigue[Rgn] * if then else (Time to Stop Adherence Fatigue > Time, 1, SWadhFtg)) Units: dmnl
180. Impact of Population Density on Hospital Availability[Rgn] = 0.72 Units: dmnl
181. Impact of Treatment on Fatality Rate[Rgn] = 0.32 Units: dmnl
182. Incubation Period = 5.6 Units: Day
183. Indicated fraction negative demand tested[Rgn] = 1 - exp (Flu Acuity * (Prob Missing Symptom[Rgn] - 1)) Units: dmnl
184. Indicated fraction positive demand tested[Rgn] = 1 - exp (Covid Acuity[Rgn] * (Prob Missing Symptom[Rgn] - 1)) Units: dmnl
185. Indicated Fraction Post Mortem Testing[Rgn] = Fraction Covid Death In Hospitals Previously Tested[Rgn] ^ Sensitivity Post Mortem Testing to Capacity[Rgn] Units: dmnl
186. Indicated Risk of Life Loss[Rgn] = Perceived Hazard of Death[Rgn] / Discount Rate per Day Units: dmnl
187. Infected pre Detection[Rgn] = INTEG(Onset of Symptoms[Rgn] - Continue without Testing[Rgn] - Symptomatic Infected to Testing[Rgn] - Untested symptomatic Infected to Hospital[Rgn], 0) Units: Person
188. “Infected Unconfirmed Post-Detection”[Rgn] = INTEG(Continue without Testing[Rgn] - Deaths of Symptomatic Untested[Rgn] - Recovery of Untested[Rgn], 0) Units: Person
189. Infection Rate[Rgn] = Infectious Contacts[Rgn] * (Susceptible[Rgn] / Population[Rgn]) * Weather Effect on Transmission[Rgn] Units: Person/Day
190. InfectionUFractionCounted[Rgn] = if then else (DataCmltOverTime[Rgn,Infection] = :NA:, 0, ZIDZ (DataCmltOverTime[Rgn,Infection], Cumulative Cases[Rgn])) Units: dmnl
191. Infectious Confirmed Not Hospitalized[Rgn] = INTEG(Positive Testing of Infected Untreated[Rgn] - Deaths of Confirmed[Rgn] - Recovery of Confirmed[Rgn], 0) Units: Person
192. Infectious Contacts[Rgn] = (“Pre-Symptomatic Infected”[Rgn] * Transmission Multiplier Presymptomatic[Rgn] + Infected pre Detection[Rgn] * Transmission Multiplier Pre Detection[Rgn] + (Additional Asymptomatic Post Detection[Rgn] + “Poisson Not-tested Asymptomatic”[Rgn]) * Baseline Risk of Transmission by Asymptomatic[Rgn] + (Infectious not Tested or in Hospitals Poisson[Rgn] - “Poisson Not-tested Asymptomatic”[Rgn]) * Baseline Transmission Multiplier for Untested Symptomatic + Infectious Confirmed Not Hospitalized[Rgn] * Transmission Multiplier for Confirmed[Rgn] + sum (Hospitalized Infectious[Rgn,TstSts!] * Transmission Multiplier for Hospitalized[Rgn,TstSts!])) * Reference Force of Infection[Rgn] * Contacts Relative to Normal[Rgn] Units: Person/Day
193. Infectious not Tested or in Hospitals Poisson[Rgn] = “Infected Unconfirmed Post-Detection”[Rgn] - Additional Asymptomatic Post Detection[Rgn] Units: Person
194. Initial Population[Rgn] = GET VDF CONSTANTS(Constant Data File, ‘DataConstants[Rgn]’, 1) Units: Person
195. INITIAL TIME = 30 Units: Day
196. InpAveErr = INITIAL(sum (InpAveErrCmp[PriorEndoAve!] * PriorCounts[PriorEndoAve!])) Units: **undefined**
197. InpAveErrCmp[PriorEndoAve] = INITIAL((abs (CalcAve[PriorEndoAve] - InputAve[PriorEndoAve]) / Max (1e-06, CalcAve[PriorEndoAve]) * SW EndoAve)) Units: **undefined**
198. InputAve[Priors] = 1, 1.8, 0.47, 1, 0.0009, 0.81, 58, 0.017, 6.2, 0.1, 0.24, 0.4, 2.27, 0.52, 0.00055, 0.76, 5.9, 2.07, 0.55, 1e-06, 1e-06, 1e-06, 0.28, 0, 0 Units: **undefined**
199. IrD : Travel,InformalDeath
200. Known death fraction in hospitals large enough = sum (if then else (Cml Known Death Frac Hosp[Rgn!] < MinHspDTreshAdv, 1, 0) * AdvCntrs[Rgn!]) Units: dmnl
201. lastTestData[Rgn] = INITIAL(GET DATA Last TIME (DataTestRate[Rgn])) Units: Day
202. LastTestDate[Rgn] = INITIAL(GET DATA Last TIME (DataTestRate[Rgn])) Units: Day
203. Learning and Death Reduction Rate[Rgn1] = 0 Units: dmnl
204. Liver Disease Impact on Fatality[Rgn] = INITIAL((Liver Disease Rate[Rgn] / MeanLiver) ^ Sens Liver Impact Net[Rgn]) Units: dmnl
205. Liver Disease Rate[Rgn] = GET VDF CONSTANTS(Constant Data File, ‘DataConstants[Rgn]’, 18) Units: dmnl
206. lnymix[Rgn,expnt] = - ln (Max (1e-06, 1 - Ymix[Rgn,expnt])) Units: dmnl
207. lnymix 0[Rgn,expnt] = - ln (Max (1e-06, 1 - Ymix[Rgn,expnt])) Units: dmnl
208. Max Test Rate per Capita = 0.001 Units: 1/Day
209. Max Time Data Used = 550 Units: Day
210. maxAlp = 1 Units: dmnl
211. MaxData[Rgn] = INITIAL(GET DATA MAX (DataCmltOverTime[Rgn,Infection], 0, 500)) Units: Person
212. MaxRTresh = 8 Units: dmnl
213. MaxVacRate[Rgn] = INITIAL(if then else (Vaccination Period < 10, 0, Initial Population[Rgn] * (1 - AntiVaxxerFrac[Rgn]) / (Vaccination Period - frac rampup * Vaccination Period / 2))) Units: Person/Day
214. MeanChronic = GET VDF CONSTANTS(Constant Data File, ‘MeanChronic’, 1) Units: dmnl
215. MeanFlowOverTime[Rgn,Infection] = Post Mortem Test Rate[Rgn] + Positive Tests of Infected[Rgn]
216. MeanFlowOverTime[Rgn,Death] = Recorded Deaths[Rgn]
217. MeanFlowOverTime[Rgn,Test] = Total Simulated Tests[Rgn] Units: Person/Day
218. MeanLiver = GET VDF CONSTANTS(Constant Data File, ‘MeanLiver’, 1) Units: dmnl
219. MeanObesity = GET VDF CONSTANTS(Constant Data File, ‘MeanObesity’, 1) Units: dmnl
220. Min Contact Fraction[Rgn] = 0.04 Units: dmnl
221. Min Excess Death Attributable to COVID = 0.5 Units: dmnl
222. Min Fatality Multiplier = 0.1 Units: dmnl
223. Min Vaccination Time = 10 Units: Day
224. MinAdjT = 1 Units: Day
225. MinHspDTresh = 0.2 Units: dmnl
226. MinHspDTreshAdv = 0.8 Units: dmnl
227. MinSuscTresh = 0.7 Units: dmnl
228. MinTimeDwngRisk = 5 Units: Day
229. Mu[Rgn,Series] = Max (eps, MeanFlowOverTime[Rgn,Series]) Units: Person/Day
230. Multiplier Recent Infections to Test[Rgn] = 45 Units: dmnl
231. Multiplier Transmission Risk for Asymptomatic[Rgn] = 0.3 Units: dmnl
232. Multiplier Transmission Risk for Asymptomatic Net[Rgn] = INITIAL(Multiplier Transmission Risk for Asymptomatic[Rgn] * (1 - SW Gen[MTrAsym]) + SW Gen[MTrAsym] * InputAve[MTrAsym]) Units: dmnl
233. NBL1[Rgn,Series] = if then else (DataFlowOverTime[Rgn,Series] = 0, - ln (1 + alp[Rgn,Series] * Mu[Rgn,Series]) / alp[Rgn,Series], 0) Units: dmnl
234. NBL2[Rgn,Series] = if then else (DataFlowOverTime[Rgn,Series] > 0, GAMMA LN (Di[Rgn,Series] + 1 / alp[Rgn,Series]) - GAMMA LN (1 / alp[Rgn,Series]) - GAMMA LN (Di[Rgn,Series] + 1) - (Di[Rgn,Series] + 1 / alp[Rgn,Series]) * ln (1 + alp[Rgn,Series] * Mu[Rgn,Series]) + Di[Rgn,Series] * (ln alp[Rgn,Series]) + ln (Mu[Rgn,Series])), 0) Units: dmnl
235. NBL3[Rgn,Series] = if then else (Di[Rgn,Series] > 0, - GAMMA LN (Di[Rgn,Series] + 1) - (Di[Rgn,Series] + 1 / alp[Rgn,Series]) * ln (1 + alp[Rgn,Series] * Mu[Rgn,Series]) + Di[Rgn,Series] * (ln (alp[Rgn,Series]) + ln (Mu[Rgn,Series])), 0) Units: dmnl
236. NBLLFlow[Rgn,Series] = (NBL1[Rgn,Series] + NBL2[Rgn,Series]) * FlowToInclude[Series] * DataIncluded[Rgn] Units: dmnl
237. Negative Test Results[Rgn] = Testing on Living[Rgn] - Positive Tests of Infected[Rgn] Units: Person/Day
238. New Cases[Rgn] = Infection Rate[Rgn] + Patient Zero Arrival[Rgn] Units: Person/Day
239. New Testing Time = 1000 Units: Day
240. Nominal Hospital Capacity[Rgn] = INITIAL(Initial Population[Rgn] * Beds per Thousand Population[Rgn] / 1000) Units: Person
241. Normalized Hospital density[Rgn] = INITIAL(Bed per Square Kilometer[Rgn] / Reference Hospital Density) Units: dmnl
242. Not too few susceptibles = sum (if then else (SuscFrac[Rgn!] < MinSuscTresh, 1, 0)) Units: dmnl
243. NSeed = 1 Units: dmnl
244. numTrial[Rgn,UsedSeries] = INITIAL(Max (1.01, 1 / alp[Rgn,UsedSeries])) Units: dmnl
245. Obesity Impact on Fatality[Rgn] = INITIAL((Obesity Rates[Rgn] / MeanObesity) ^ Sens Obesity Impact Net[Rgn]) Units: dmnl
246. Obesity Rates[Rgn] = GET VDF CONSTANTS(Constant Data File, ‘DataConstants[Rgn]’, 16) Units: dmnl
247. Onset of Symptoms[Rgn] = DELAY N (Infection Rate[Rgn], Incubation Period, 0, Delay Order) Units: Person/Day
248. Onset to Detection Delay = 5 Units: Day
249. OtherVaccination[Rgn] = Vaccination On[Rgn] * min (Total Vaccination Rate[Rgn] * (1 - FracNotVaccinated Susceptible[Rgn]), RemainingFractionForVaccine[Rgn] * (Initial Population[Rgn] - Vaccinated[Rgn] - Susceptible[Rgn]) / Min Vaccination Time) Units: Person/Day
250. Overall Death Fraction[Rgn] = ZIDZ (Death Rate[Rgn], All Recovery[Rgn]) Units: dmnl
251. Patient Zero Arrival[Rgn] = if then else (Time < Patient Zero Arrival Time[Rgn] :AND: Time + TIME STEP >= Patient Zero Arrival Time[Rgn], PatientZero / TIME STEP, 0) Units: Person/Day
252. Patient Zero Arrival Time[Rgn] = 1 Units: Day [0,200]
253. PatientZero = 1 Units: Person
254. payoff = 0 Units: dmnl
255. pdim : tstP,dfcP,dgtP,scuP
256. Perceived Hazard of Death[Rgn] = (Weight on Reported Probability of Infection[Rgn] * Reported Hazard of Death[Rgn] + (1 - Weight on Reported Probability of Infection[Rgn]) * True Hazard of death[Rgn]) Units: 1/Day
257. Perceived Risk of Life Loss[Rgn] = INTEG((Indicated Risk of Life Loss[Rgn] - Perceived Risk of Life Loss[Rgn]) / if then else (Indicated Risk of Life Loss[Rgn] > Perceived Risk of Life Loss[Rgn], Time to Upgrade Risk[Rgn], Time to Downgrade Risk With Vaccine[Rgn]), 0) Units: dmnl
258. PG1 : PG
259. PMAS Confirmed for Hospital Demand[Rgn] = (1 - Reference COVID Hospitalization Fraction Confirmed[Rgn]) ^ (1 / Average Acuity of Positively Tested[Rgn]) Units: dmnl
260. PMAS Unconfirmed for Hospital Demand[Rgn] = PMAS Confirmed for Hospital Demand[Rgn] + (1 - PMAS Confirmed for Hospital Demand[Rgn]) * Untested PMAS Gap with Tested[Rgn] Units: dmnl
261. “Poisson Not-tested Asymptomatic”[Rgn] = Infectious not Tested or in Hospitals Poisson[Rgn] * exp (− Average Acuity Not Hospitalized Poisson[Rgn,Notest]) Units: Person
262. Poisson Subset Not Tested Passing Gate[Rgn] = Poisson Subset Reaching Test Gate[Rgn] - Positive Tests of Infected[Rgn] Units: Person/Day
263. Poisson Subset Reaching Test Gate[Rgn] = Reaching Testing Gate[Rgn] / (1 + Additional Asymptomatic Relative to Symptomatic[Rgn]) Units: Person/Day
264. Policy Test Rate[Rgn] = if then else (Time < New Testing Time, Current Test Rate per Capita[Rgn], Final Test Rate Per Capita[Rgn]) Units: 1/Day
265. Population[Rgn] = Infected pre Detection[Rgn] + “Infected Unconfirmed Post-Detection”[Rgn] + Susceptible[Rgn] + Recovered Unconfirmed[Rgn] + Confirmed Recovered[Rgn] + Infectious Confirmed Not Hospitalized[Rgn] + “Pre-Symptomatic Infected”[Rgn] + sum (Hospitalized Infectious[Rgn,TstSts!]) + sum (Cumulative Recovered at Hospitals[Rgn,TstSts!]) Units: Person
266. PopulationCheck[Rgn] = Recovered Unconfirmed[Rgn] + Confirmed Recovered[Rgn] + sum (Cumulative Recovered at Hospitals[Rgn,TstSts!]) + Different Infectious Counted[Rgn] + Susceptible[Rgn] Units: Person
267. Positive Candidates Interested in Testing Poisson Subset[Rgn] = Poisson Subset Reaching Test Gate[Rgn] * Fraction Seeking Test[Rgn] Units: Person/Day
268. Positive Candidates Interested in Testing Poisson Subset Adj[Rgn] = Max (0.001 * Potential Test Demand from Susceptible Population[Rgn], Positive Candidates Interested in Testing Poisson Subset[Rgn]) Units: Person/Day
269. Positive Testing of Infected Untreated[Rgn] = Positive Tests of Infected[Rgn] * Fraction Poisson not Hospitalized[Rgn,Tested] Units: Person/Day
270. Positive Tests of Infected[Rgn] = Positive Candidates Interested in Testing Poisson Subset[Rgn] * (1 - Fraction Interseted not Correctly Tested[Rgn]) Units: Person/Day
271. Post Mortem Test Delay = 1 Units: Day
272. Post Mortem Test Rate[Rgn] = Post Mortem Tests Total[Rgn] * Sensitivity of COVID Test Units: Person/Day
273. Post Mortem Test Untreated[Rgn] = Post Mortem Test Rate[Rgn] * Frac Post Mortem from Untreated[Rgn] Units: Person/Day
274. Post Mortem Testing Need[Rgn] = SMOOTHI ((Deaths of Symptomatic Untested[Rgn] + Hospitalized Infectious Deaths[Rgn,Notest]) * Fraction of Fatalities Screened Post Mortem[Rgn], Post Mortem Test Delay, 0) Units: Person/Day
275. Post Mortem Tests Total[Rgn] = min (Post Mortem Testing Need[Rgn], Active Test Rate[Rgn]) Units: Person/Day
276. “Post-Detection Phase Resolution Time” = 10 Units: Day
277. PostMortemCorrection[Rgn] = min (Hospitalized Infectious[Rgn,Notest] / MinAdjT, Post Mortem Test Rate[Rgn] * (1 - Frac Post Mortem from Untreated[Rgn])) Units: Person/Day
278. Potential Hospital Demand[Rgn,Notest] = Hospital Demand from Not Tested[Rgn]
279. Potential Hospital Demand[Rgn,Tested] = Hospital Demand from Tested[Rgn] Units: Person/Day
280. Potential Test Demand from Susceptible Population[Rgn] = (Susceptible[Rgn] + Recovered Unconfirmed[Rgn] + Cumulative Recovered at Hospitals[Rgn,Notest]) * (Baseline Daily Fraction Susceptible Seeking Tests[Rgn] * Fraction Seeking Test[Rgn] + Multiplier Recent Infections to Test[Rgn] / Population[Rgn] * Recent Detected Infections[Rgn]) Units: Person/Day
281. “Pre-Symptomatic Infected”[Rgn] = INTEG(Infection Rate[Rgn] + Patient Zero Arrival[Rgn] - Onset of Symptoms[Rgn], 0) Units: Person
282. PriorCounts[PriorEndoAve] = INITIAL(if then else (PriorEndoAve < 26, 1, 0)) Units: dmnl
283. PriorEndoAve : UpAdj,DwnAdj,RFI,RfSkTs,WRpPIn,MInfTs,MnCnFrc,SnCnRdUt,CfImCn,ImPDnHs,ImTrFt,DrdFac,MxHsFr,B sFtRt,SnsWth,Acty,SnFtAc,TtAsyFr,ObsImp,ChrImp,LivImp,MTrAsym,HspLrng,AdhrFtg
284. PriorErrs[Rgn,Priors] = INITIAL(ZIDZ (ActiveAve[Priors] - RegionalInputs[Priors,Rgn], (AbsStd[Priors] * StdScale)) ^ 2 / 2) Units: **undefined**
285. PriorGen : BsFtRt,SnsWth,Acty,SnFtAc,TtAsyFr,ObsImp,ChrImp,LivImp,MTrAsym
286. Priors : UpAdj,DwnAdj,RFI,PMT,RfSkTs,WRpPIn,MInfTs,MnCnFrc,SnCnRdUt,CfImCn,ImPDnHs,ImTrFt,DrdFac,MxHsFr,BsFtRt,SnsWth,Acty,SnFtAc,TtAsyFr,ObsImp,ChrImp,LivImp,MTrAsym,HspLrng,AdhrFtg
287. Prob Missing Symptom[Rgn] = Max (0, ln (Y[Rgn]) / Flu Acuity + 1) Units: dmnl
288. Probability of Missing Acuity Signal at Hospitals[Rgn,Tested] = ZIDZ (ln (Max (1e-06, 1 - ZIDZ (Hospital Admits All[Rgn,Tested], Positive Tests of Infected[Rgn]))), Average Acuity of Positively Tested[Rgn]) + 1
289. Probability of Missing Acuity Signal at Hospitals[Rgn,Notest] = ZIDZ (ln (Max (1e-06, 1 - ZIDZ (Hospital Admits All[Rgn,Notest], Poisson Subset Not Tested Passing Gate[Rgn]))), Average Acuity of Untested Poisson Subset[Rgn]) + 1 Units: dmnl
290. PseudoCFR Units: dmnl
291. R Effective Reproduction Rate[Rgn] = ZIDZ (Infection Rate[Rgn], Total Weighted Infected Population[Rgn]) * Total Disease Duration Units: dmnl
292. RandFlowTime = 1000 Units: Day
293. Reaching Testing Gate[Rgn] = Infected pre Detection[Rgn] / Onset to Detection Delay Units: Person/Day
294. Realistic R0 = sum (if then else (R Effective Reproduction Rate[Rgn!] > MaxRTresh, 1, 0)) Units: dmnl
295. Recent Detected Infections[Rgn] = SMOOTHI (Positive Tests of Infected[Rgn], Time to Respond with Tests, 0) Units: Person/Day
296. Recent Relative Contacts[Rgn] = SMOOTHI (Contacts Relative to Normal[Rgn], Adherence Fatigue Time, 1) Units: dmnl
297. Recorded Deaths[Rgn] = Post Mortem Test Rate[Rgn] + Deaths of Confirmed[Rgn] + Hospitalized Infectious Deaths[Rgn,Tested] Units: Person/Day
298. Recovered Unconfirmed[Rgn] = INTEG(Recovery of Untested[Rgn], 0) Units: Person
299. Recovery of Confirmed[Rgn] = Tested Untreated Resolution[Rgn] * (1 - Fatality Rate Untreated[Rgn,Tested]) Units: Person/Day
300. Recovery of Untested[Rgn] = (“Infected Unconfirmed Post-Detection”[Rgn] / “Post-Detection Phase Resolution Time”) - Deaths of Symptomatic Untested[Rgn] Units: Person/Day
301. Reference COVID Hospitalization Fraction Confirmed[Rgn] = 0.7 Units: dmnl
302. Reference Force of Infection[Rgn] = 0.6 Units: 1/Day [0,2]
303. Reference Hospital Density = 6.06 Units: Person/(Km*Km)
304. RegionalInputs[UpAdj,Rgn] = INITIAL(Log (Time to Upgrade Risk[Rgn], 10))
305. RegionalInputs[DwnAdj,Rgn] = Log (Time to Downgrade Risk[Rgn], 10)
306. RegionalInputs[RFI,Rgn] = Reference Force of Infection[Rgn]
307. RegionalInputs[PMT,Rgn] = Sensitivity Post Mortem Testing to Capacity[Rgn]
308. RegionalInputs[RfSkTs,Rgn] = Baseline Daily Fraction Susceptible Seeking Tests[Rgn]
309. RegionalInputs[WRpPIn,Rgn] = Weight on Reported Probability of Infection[Rgn]
310. RegionalInputs[MInfTs,Rgn] = Multiplier Recent Infections to Test[Rgn]
311. RegionalInputs[MnCnFrc,Rgn] = Min Contact Fraction[Rgn]
312. RegionalInputs[SnCnRdUt,Rgn] = Sensitivity of Contact Reduction to Utility[Rgn]
313. RegionalInputs[CfImCn,Rgn] = Confirmation Impact on Contact[Rgn]
314. RegionalInputs[ImPDnHs,Rgn] = Impact of Population Density on Hospital Availability[Rgn]
315. RegionalInputs[ImTrFt,Rgn] = Impact of Treatment on Fatality Rate[Rgn]
316. RegionalInputs[DrdFac,Rgn] = Log (Dread Factor in Risk Perception[Rgn], 10)
317. RegionalInputs[MxHsFr,Rgn] = Reference COVID Hospitalization Fraction Confirmed[Rgn]
318. RegionalInputs[BsFtRt,Rgn] = Base Fatality Rate for Unit Acuity Net[Rgn]
319. RegionalInputs[SnsWth,Rgn] = Sensitivity to Weather Net[Rgn]
320. RegionalInputs[Acty,Rgn] = Covid Acuity Relative to Flu Init Net[Rgn]
321. RegionalInputs[SnFtAc,Rgn] = Sensitivity of Fatality Rate to Acuity Net[Rgn]
322. RegionalInputs[TtAsyFr,Rgn] = Total Asymptomatic Fraction Init Net[Rgn]
323. RegionalInputs[ObsImp,Rgn] = Sens Obesity Impact Net[Rgn]
324. RegionalInputs[ChrImp,Rgn] = Sens Chronic Impact Net[Rgn]
325. RegionalInputs[LivImp,Rgn] = Sens Liver Impact Net[Rgn]
326. RegionalInputs[MTrAsym,Rgn] = Multiplier Transmission Risk for Asymptomatic Net[Rgn]
327. RegionalInputs[HspLrng,Rgn] = Learning and Death Reduction Rate[Rgn]
328. RegionalInputs[AdhrFtg,Rgn] = Strength of Adherence Fatigue[Rgn] Units: **undefined**
329. Relative Risk of Transmission by Hospitalized = 1 Units: dmnl
330. Relative Risk of Transmission by Presymptomatic = 1 Units: dmnl
331. RemainingFractionForVaccine[Rgn] = (1 - AntiVaxxerFrac[Rgn]) - Vaccinated Fraction[Rgn] Units: dmnl
332. Reported Hazard of Death[Rgn] = SimFlowOverTime[Rgn,Death] / Population[Rgn] Units: 1/Day
333. Response Policy Time On = 1000 Units: Day
334. Response Policy Weight = 0 Units: dmnl
335. Rgn : Argentina,Australia,Austria,Bahrain,Bangladesh,Belarus,Belgium,Bolivia,Bulgaria,Canada,Chile,Colombia,CostaRica, Croatia,Cuba,Cyprus,CzechRepublic,Denmark,DominicanRepublic,Ecuador,ElSalvador,Estonia,Ethiopia,Finland,France,Germany,Ghana,Greece,Hungary,Iceland,India,Indonesia,Iran,Iraq,Ireland,Israel,Italy,Jamaica,Japan,Kazakhstan,Kenya,Kuwait,Latvia,Lithuania,Luxembourg,Madagascar,Malawi,Malaysia,Maldives,Malta,Mexico,Morocco,Mozambique,Nepal,Netherlands,NewZealand,Nigeria,NorthMacedonia,Norway,Pakistan,Panama,Paraguay,Peru,Philippi nes,Poland,Portugal,Qatar,Romania,Russia,Rwanda,SaudiArabia,Senegal,Serbia,Singapore,Slovakia,Slovenia,SouthAfrica,SouthKorea,Spain,SriLanka,Sweden,Switzerland,Thailand,Togo,Tunisia,Turkey,UAE,UK,Ukraine,Uruguay,USA, Zambia
336. Rgn1 : Rgn
337. Risk threshold for response[Rgn] = if then else (Response Policy Time On < Time, (1 - Response Policy Weight) * Sensitivity of Contact Reduction to Utility[Rgn], Sensitivity of Contact Reduction to Utility[Rgn]) / Impact of Adherence Fatigue[Rgn] Units: dmnl
338. SAVEPER = 1 Units: Day [0,?]
339. Sens Chronic Impact = 1e-06 Units: dmnl
340. Sens Chronic Impact Net[Rgn] = INITIAL(Sens Chronic Impact * (1 - SW Gen[ChrImp]) + SW Gen[ChrImp] * InputAve[ChrImp]) Units: dmnl
341. Sens Liver Impact = 1e-06 Units: dmnl
342. Sens Liver Impact Net[Rgn] = INITIAL(Sens Liver Impact * (1 - SW Gen[LivImp]) + SW Gen[LivImp] * InputAve[LivImp]) Units: dmnl
343. Sens Obesity Impact = 1e-06 Units: dmnl
344. Sens Obesity Impact Net[Rgn] = INITIAL(Sens Obesity Impact * (1 - SW Gen[ObsImp]) + SW Gen[ObsImp] * InputAve[ObsImp]) Units: dmnl
345. SensCovidUntestedAdmission = 1 Units: dmnl
346. Sensitivity of Contact Reduction to Utility[Rgn] = 15 Units: dmnl
347. Sensitivity of Contact Reduction to Utility Policy = 10 Units: dmnl
348. Sensitivity of COVID Test = 0.7 Units: dmnl
349. Sensitivity of Fatality Rate to Acuity[Rgn] = 2 Units: dmnl
350. Sensitivity of Fatality Rate to Acuity Net[Rgn] = INITIAL(Sensitivity of Fatality Rate to Acuity[Rgn] * (1 - SW Gen[SnFtAc]) + SW Gen[SnFtAc] * InputAve[SnFtAc]) Units: dmnl [0,3]
351. Sensitivity Post Mortem Testing to Capacity[Rgn] = 1 Units: dmnl
352. Sensitivity to Weather = 0.76 Units: dmnl
353. Sensitivity to Weather Net[Rgn] = INITIAL(Sensitivity to Weather * (1 - SW Gen[SnsWth]) + SW Gen[SnsWth] * InputAve[SnsWth]) Units: dmnl
354. Series : Infection,Death,Test
355. SeriesErrorTerm[Rgn,Series] = if then else (DataCmltOverTime[Rgn,Series] = :NA:, 0, (abs (DataCmltOverTime[Rgn,Series] - SimCmltOverTime[Rgn,Series])) ^ CmltErrPW) / (BaseError + DataCmltOverTime[Rgn,Series]) * CmltPenaltyScl * CmltToInclude[Series] * DataIncluded[Rgn] Units: dmnl
356. Sim Pseudo Case Fatality[Rgn] = ZIDZ (Cumulative Deaths of Confirmed[Rgn], Cumulative Confirmed Cases[Rgn]) Units: dmnl
357. SimCmltOverTime[Rgn,Infection] = Cumulative Confirmed Cases[Rgn]
358. SimCmltOverTime[Rgn,Death] = Cumulative Deaths of Confirmed[Rgn]
359. SimCmltOverTime[Rgn,Test] = Cumulative Tests Conducted[Rgn] Units: Person
360. SimFlowOverTime[Rgn,UsedSeries] = if then else (SwitchRandFlowTime < Time, RANDOM NEGATIVE BINOMIAL (−1, 1e+06, successP[Rgn,UsedSeries], numTrial[Rgn,UsedSeries], 0, 1, NSeed), Mu[Rgn,UsedSeries]) Units: Person/Day
361. SimTestRate[Rgn] = Total Simulated Tests[Rgn] Units: Person/Day
362. SqrdErr[Rgn,Series] = if then else (FlowResiduals[Rgn,Series] = :NA:, :NA:, FlowResiduals[Rgn,Series] ^ 2) Units: Person*Person/(Day*Day)
363. StdScale = 1 Units: dmnl
364. StopDataUseTime[Rgn] = INITIAL(min (lastTestData[Rgn], Max Time Data Used))Units: Day
365. Strength of Adherence Fatigue[Rgn] = 0 Units: dmnl
366. successP[Rgn,UsedSeries] = 1 / (1 + alp[Rgn,UsedSeries] * Mu[Rgn,UsedSeries]) Units: dmnl
367. Susceptible[Rgn] = INTEG(− Infection Rate[Rgn] - Susceptible Vaccination[Rgn], Initial Population[Rgn]) Units: Person
368. Susceptible Vaccination[Rgn] = Vaccination On[Rgn] * min (Total Vaccination Rate[Rgn] * FracNotVaccinated Susceptible[Rgn], RemainingFractionForVaccine[Rgn] * Susceptible[Rgn] / Min Vaccination Time) Units: Person/Day
369. SuscFrac[Rgn] = Susceptible[Rgn] / Population[Rgn] Units: dmnl
370. SW EndoAve = INITIAL(if then else (ELMCOUNT(Rgn) > 1, 1, 0)) Units: dmnl
371. SW Gen[PriorGen] = 0 Units: dmnl
372. SWadhFtg = 0 Units: dmnl
373. Switch for Government Response[Rgn] = if then else (Time > Government Response Start Time[Rgn], 1, 0) Units: dmnl
374. SwitchRandFlow = 0 Units: dmnl
375. SwitchRandFlowTime = if then else (SwitchRandFlow = 1, min (Max Time Data Used, RandFlowTime), 1000) Units: Day
376. Sympthom Reduction by Infectioun[Rgn] = Average Acuity in Susceptible[Rgn] * Infection Rate[Rgn] Units: Person/Day
377. Sympthom Reduction by Vaccination[Rgn] = Susceptible Vaccination[Rgn] * Average Acuity in Susceptible[Rgn] * Vaccination Priority Multiplier Units: Person/Day
378. Sympthoms in Susceptible[Rgn] = INTEG(− Sympthom Reduction by Infectioun[Rgn] - Sympthom Reduction by Vaccination[Rgn], Susceptible[Rgn] * (1 - Additional Asymptomatic Fraction Init[Rgn]) * Covid Acuity Relative to Flu Init Net[Rgn]) Units: Person
379. Symptomatic Fraction in Poisson[Rgn] = INITIAL(1 - exp (− Covid Acuity[Rgn])) Units: dmnl
380. Symptomatic Fraction Negative[Rgn] = INITIAL(1 - exp (− Flu Acuity)) Units: dmnl
381. Symptomatic Infected to Testing[Rgn] = Positive Testing of Infected Untreated[Rgn] + Hospital Admission Infectious[Rgn,Tested] Units: Person/Day
382. t3[Rgn] = (−9 * b[Rgn] + 1.7321 * Sqrt (4 * a[Rgn] ^ 3 + 27 * b[Rgn] ^ 2)) ^ (1 / 3) Units: dmnl
383. talp = 5 Units: dmnl
384. Tested Untreated Resolution[Rgn] = Infectious Confirmed Not Hospitalized[Rgn] / “Post-Detection Phase Resolution Time” Units: Person/Day
385. TestErrorFrac = 0.0001 Units: dmnl
386. TestFlowErr[Rgn] = ((DataFlowOverTime[Rgn,Test] - MeanFlowOverTime[Rgn,Test]) * WTestFlowErr[Rgn]) ^ 2 Units: dmnl
387. Testing Capacity Net of Post Mortem Tests[Rgn] = Active Test Rate[Rgn] - Post Mortem Tests Total[Rgn] Units: Person/Day
388. Testing Demand[Rgn] = Positive Candidates Interested in Testing Poisson Subset[Rgn] * Symptomatic Fraction in Poisson[Rgn] + Potential Test Demand from Susceptible Population[Rgn] * Symptomatic Fraction Negative[Rgn] Units: Person/Day
389. Testing on Living[Rgn] = min (Testing Capacity Net of Post Mortem Tests[Rgn], Testing Demand[Rgn]) Units: Person/Day
390. Tests on Negative Patients[Rgn] = Testing on Living[Rgn] * ZIDZ (Indicated fraction negative demand tested[Rgn] * Potential Test Demand from Susceptible Population[Rgn], Indicated fraction negative demand tested[Rgn] * Potential Test Demand from Susceptible Population[Rgn] + Indicated fraction positive demand tested[Rgn] * Positive Candidates Interested in Testing Poisson Subset[Rgn]) Units: Person/Day
391. Tests Per Million[Rgn] = Cumulative Tests Data[Rgn] / Initial Population[Rgn] * 1e+06Units: dmnl
392. ThrsInc[Rgn] = Max (FracThresh * MaxData[Rgn], 50) Units: Person
393. TIME STEP = 0.25 Units: Day [0,?]
394. Time to Adjust Testing = 30 Units: Day
395. Time to Downgrade Risk[Rgn] = 60 Units: Day
396. Time to Downgrade Risk Net[Rgn] = if then else (Response Policy Time On < Time, (1 + Response Policy Weight) * Time to Downgrade Risk[Rgn], Time to Downgrade Risk[Rgn]) Units: Day
397. Time to Downgrade Risk Policy = 300 Units: Day
398. Time to Downgrade Risk With Vaccine[Rgn] = Time to Downgrade Risk Net[Rgn] * (1 - (1 - SuscFrac[Rgn]) * Vaccination On[Rgn]) + (1 - SuscFrac[Rgn]) * Vaccination On[Rgn] * MinTimeDwngRisk Units: Day
399. Time to Herd Immunity[Rgn] = XIDZ (Herd Immunity Fraction * Susceptible[Rgn], Total Weighted Infected Population[Rgn] / Total Disease Duration, 0) Units: Day
400. Time to Respond with Tests = 5 Units: Day
401. Time to Stop Adherence Fatigue = 1000 Units: Day
402. Time to Upgrade Risk[Rgn] = 10 Units: Day
403. Time variant change in fatality[Rgn] = Max (Min Fatality Multiplier, (Max (Baseline Cumulative Cases for Learning, Cumulative Cases[Rgn] / Initial Population[Rgn]) / Baseline Cumulative Cases for Learning) ^ (− Learning and Death Reduction Rate[Rgn])) Units: dmnl
404. TimeVar Impact of Treatment on Fatality[Rgn] = Impact of Treatment on Fatality Rate[Rgn] * Time variant change in fatality[Rgn] Units: dmnl
405. Total Asymptomatic Fraction[Rgn] = 0.5 Units: dmnl
406. Total Asymptomatic Fraction Init Net[Rgn] = INITIAL(Total Asymptomatic Fraction[Rgn] * (1 - SW Gen[TtAsyFr]) + SW Gen[TtAsyFr] * InputAve[TtAsyFr]) Units: dmnl
407. Total Asymptomatic Fraction Net[Rgn] = Additional Asymptomatic Fraction Init[Rgn] + exp (− Covid Acuity[Rgn]) * (1 - Additional Asymptomatic Fraction Init[Rgn]) Units: dmnl
408. Total Covid Hospitalized[Rgn] = sum (Hospitalized Infectious[Rgn,TstSts!]) Units: Person
409. Total Disease Duration = Onset to Detection Delay + “Post-Detection Phase Resolution Time” + Incubation Period Units: Day
410. Total Simulated Tests[Rgn] = Post Mortem Tests Total[Rgn] + Testing on Living[Rgn] Units: Person/Day
411. Total Test on Covid Patients[Rgn] = Max (0, min (Positive Candidates Interested in Testing Poisson Subset[Rgn], Testing on Living[Rgn] - Tests on Negative Patients[Rgn])) Units: Person/Day
412. Total to Official Cases Simulated[Rgn] = ZIDZ (Cumulative Cases[Rgn], SimCmltOverTime[Rgn,Infection]) Units: dmnl
413. Total Vaccination Rate[Rgn] = if then else (Vaccination Period < 10, 0, MaxVacRate[Rgn] * (1 - min (1, Max (0, (frac rampup * Vaccination Period - (Time - Vaccine Start Time)) / (Vaccination Period * frac rampup))))) Units: Person/Day
414. Total Weighted Infected Population[Rgn] = Infected pre Detection[Rgn] + “Pre-Symptomatic Infected”[Rgn] + Weighted Infected Post Detection Gate[Rgn] Units: Person
415. Transmission Multiplier for Confirmed[Rgn] = INITIAL(Baseline Transmission Multiplier for Untested Symptomatic * Confirmation Impact on Contact[Rgn]) Units: dmnl
416. Transmission Multiplier for Hospitalized[Rgn,TstSts] = INITIAL(Baseline Transmission Multiplier for Untested Symptomatic * Relative Risk of Transmission by Hospitalized * if then else (TstSts = 1, Confirmation Impact on Contact[Rgn], 1)) Units: dmnl
417. Transmission Multiplier Pre Detection[Rgn] = INITIAL(Baseline Transmission Multiplier for Untested Symptomatic * (1 - Total Asymptomatic Fraction Net[Rgn]) + Total Asymptomatic Fraction Net[Rgn] * Baseline Risk of Transmission by Asymptomatic[Rgn]) Units: dmnl
418. Transmission Multiplier Presymptomatic[Rgn] = INITIAL((Baseline Transmission Multiplier for Untested Symptomatic * Relative Risk of Transmission by Presymptomatic) * (1 - Total Asymptomatic Fraction Net[Rgn]) + Total Asymptomatic Fraction Net[Rgn] * Baseline Risk of Transmission by Asymptomatic[Rgn] * Relative Risk of Transmission by Presymptomatic) Units: dmnl
419. True Hazard of death[Rgn] = Death Rate[Rgn] / Population[Rgn] Units: 1/Day
420. TstInc[Rgn] = Active Test Rate[Rgn] Units: Person/Day
421. TstSts : Tested,Notest
422. Untested PMAS Gap with Tested[Rgn] = (1 - Allocated Fration NonCOVID Hospitalized[Rgn]) ^ SensCovidUntestedAdmission Units: dmnl
423. Untested symptomatic Infected to Hospital[Rgn] = Hospital Admission Infectious[Rgn,Notest] Units: Person/Day
424. UsedSeries : Infection,Death
425. Vaccinated[Rgn] = INTEG(OtherVaccination[Rgn] + Susceptible Vaccination[Rgn], 0) Units: Person
426. Vaccinated Fraction[Rgn] = Vaccinated[Rgn] / Initial Population[Rgn] Units: dmnl
427. Vaccination On[Rgn] = if then else (Time < Vaccine Start Time, 0, 1) Units: dmnl
428. Vaccination Period = 150 Units: Day
429. Vaccination Priority Multiplier = 1.5 Units: dmnl
430. Vaccine Start Time = 800 Units: Day
431. VacWinStart[Rgn] = if then else (Vaccination Period > 1, if then else (Time > Vaccine Start Time, 2, -1), -1) Units: dmnl
432. Voluntary Reduction in Contacts[Rgn] = exp (− Max (0, Dread Factor in Risk Perception Net[Rgn] * Perceived Risk of Life Loss[Rgn] - Risk threshold for response[Rgn])) * (1 - Min Contact Fraction[Rgn]) + Min Contact Fraction[Rgn] Units: dmnl
433. W Ave Acuity Hospitalized[Rgn] = ZIDZ (sum (Average Acuity Hospitalized[Rgn,TstSts!] * Hospitalized Infectious[Rgn,TstSts!]), sum (Hospitalized Infectious[Rgn,TstSts!])) Units: dmnl
434. Weather Effect on Transmission[Rgn] = CRW[Rgn] ^ Sensitivity to Weather Net[Rgn] Units: dmnl
435. Weight Max in Test Goal = 0 Units: dmnl
436. Weight on Reported Probability of Infection[Rgn] = 0.78 Units: dmnl [0,1,0.01]
437. Weighted Infected Post Detection Gate[Rgn] = “Infected Unconfirmed Post-Detection”[Rgn] + Infectious Confirmed Not Hospitalized[Rgn] + sum (Hospitalized Infectious[Rgn,TstSts!]) * “Post-Detection Phase Resolution Time” / Hospitalized Resolution Time Units: Person
438. WTestFlowErr[Rgn] = if then else (DataFlowOverTime[Rgn,Test] = :NA:, 0, 1 / Max (10, DataFlowOverTime[Rgn,Test] * TestErrorFrac)) Units: Day/Person
439. Y[Rgn] = min (1, Max (1e-06, 1 - exp (− Extrapolated Estimator[Rgn]))) Units: dmnl
440. Ymix[Rgn,p2] = - b[Rgn] / (1 + a[Rgn])
441. Ymix[Rgn,p3] = (Sqrt (a[Rgn] ^ 2 - 4 * b[Rgn]) - a[Rgn]) / 2
442. Ymix[Rgn,p4] = (−0.87358 * a[Rgn]) / t3[Rgn] + 0.38157 * t3[Rgn] Units: dmnl

## Footnotes

1 In the equations below we use short-hand to simplify mathematical notations. The full model documentation uses full variable names. Table S1 provides the mapping between the short-hand and the full names, as well as the sources and equations for the variables and parameters discussed below.

2 https://blog.ons.gov.uk/2020/03/31/counting-deaths-involving-the-coronavirus-covid-19/

3 https://www.cdc.gov/coronavirus/2019-ncov/cases-updates/previous-testing-in-us.html

4 See e.g. https://www.wcvb.com/article/massachusetts-coronavirus-reporting-delay-due-to-quest-lab-it-glitch/32288903#

5 It may be argued that there are weekly cycles in large-scale human behaviour that may drive some true weekly cyclicality in the true rates of infection and death, and as such it may be wrong to consider such cycles to be artefacts of the data- generation process. However, we find this unlikely for a few reasons. First, weekly cycles in human interactions, largely driven by the work and school week and weekend, will have been significantly attenuated by widespread adoption of social distancing measures around the world. Second and more importantly, variation in incubation period and time before development of symptoms means that any true cyclicality in the timing of initial infection will be further attenuated in the timing of symptom development. By the same logic, wide variability in the delay from symptom development to death means there should be minimal cyclicality, if any, in the timing of deaths, meaning any such cycles visible in the data are due to measurement and reporting lags.

6 https://docs.scipy.org/doc/scipy/reference/generated/scipy.interpolate.PchipInterpolator.html

7 Cleveland R.B., Cleveland W.S., McRae J.E., Terpenning I. (1990) STL: A seasonal-trend decomposition procedure based on Loess. *J Off Stat* 6: 3-73

8 https://www.statsmodels.org/stable/generated/statsmodels.tsa.seasonal.STL.html

